# The impact of the COVID-19 pandemic on short-term cancer survival in the United Kingdom: a cohort analysis

**DOI:** 10.1101/2023.09.14.23295563

**Authors:** Nicola L Barclay, Theresa Burkard, Edward Burn, Antonella Delmestri, Andrea Miquel Dominguez, Asieh Golozar, Carlos Guarner-Argente, Francesc Xavier Avilés-Jurado, Wai Yi Man, Àlvar Roselló Serrano, Andreas Weinberger Rosen, Eng Hooi Tan, Ilona Tietzova, OPTIMA Consortium, Daniel Prieto Alhambra, Danielle Newby

**Author notes:** **Correspondence to:** Professor Daniel Prieto Alhambra, Centre for Statistics in Medicine, Nuffield Department of Orthopaedics, Rheumatology and Musculoskeletal Sciences, University of Oxford, Oxford, OX3 7LD, United Kingdom. Joint senior authors (Daniel Prieto Alhambra, Danielle Newby).

## Abstract

**Objectives:** The COVID-19 pandemic profoundly affected healthcare systems and patients. There is a pressing need to comprehend the collateral effects of the pandemic on noncommunicable diseases. Here we examined the impact of the COVID-19 pandemic on shortterm cancer survival in the United Kingdom (UK). We hypothesised that short-term survival from nine cancers would be reduced during the pandemic, particularly cancers that benefit from screening and early detection (e.g., breast and colorectal cancer).

**Design:** Population-based cohort study.

**Setting:** Electronic health records from UK primary care Clinical Practice Research Datalink (CPRD) GOLD database.

**Participants:** There were 12,259,744 eligible patients aged ≥18 years with ≥one year of prior history identified from January 2000 to December 2021.

**Main outcome measures:** We estimated age-standardised incidence rates (IR) and short-term (one- and two-year) survival of several common cancers (breast, colorectal, head and neck, liver, lung, oesophagus, pancreatic, prostate, and stomach cancer) from 2000 to 2019 (in five-year strata) compared to 2020 to 2021 using the Kaplan-Meier method.

**Results:** Apart from pancreatic cancer, IRs decreased for all cancers in 2020 and recovered to different extents in 2021. Short-term survival improved for most cancers between 2000 to 2019, but then declined for those diagnosed in 2020 to 2021.This was most pronounced for colorectal cancer, with one-year survival falling from 79.3% [95% confidence interval: 78.5%-80.1%] in 2015 to 2019 to 76.3% [74.6%-78.1%] for those diagnosed in 2020 to 2021.

**Conclusion:** Short-term survival for many cancers was impacted by the management of the COVID-19 pandemic in the UK. This decline was most prominent for colorectal cancer, with reductions in survivorship equivalent to returning to mortality seen in the first decade of the 2000s. These results illustrate the need for an immediate and well-funded investment in resolving the current backlog in cancer screening and diagnostic procedures in the UK National Health Service to improve patient outcomes.

## Introduction

The COVID-19 pandemic in late 2019 has left a permanent mark on global public health, with profound implications for healthcare systems and patients. Whilst healthcare systems grappled with the overwhelming demands of treating COVID-19 patients, there has emerged a pressing need to comprehend the collateral effects of the pandemic on non-communicable diseases, including cancer. Social restrictions and the prioritization of urgent over elective healthcare provision imposed by the pandemic impacted screening and diagnostic pathways for many cancers worldwide. Previous work has revealed that the number of routinely performed screening and diagnostic tests for various cancers were reduced during the first lockdown, particularly mammograms and colonoscopies [1-3]. Worryingly, rates of testing and screening in the United Kingdom (UK) have remained below those observed before the pandemic from the first lockdown until December 2021 [2]. Reduced screening and diagnostic tests inevitably cause delays leading to more advanced stage at diagnosis and worsened prognosis.

Short-term survival from cancer has generally improved over the past two decades [4]. The improvement in cancer survival can be attributed to several interconnected factors that have contributed to advancements in cancer prevention: screening, early diagnosis, and treatment [5]. These factors have collectively led to better outcomes and increased survival rates for cancer patients. However, the pandemic’s disruptive impact on healthcare systems and cancer care delivery has raised concerns about delayed diagnosis, altered treatment regimens, delays in surgery, and reduced access to essential services all of which may have adverse consequences for cancer patients’ survival.

The UK’s National Health Service (NHS) was already under pressure prior to the pandemic, suffering from years of reduced funding, and substantial challenges including staff shortages and reduced hospital beds per capita [6]. Staffing issues were further compounded because many doctors and nurses were on sick leave or isolating due to COVID-19 exposure. Surgical staff were redeployed to care for COVID-19 patients, and operating theatres and outpatient clinics were closed to free up resources for patients with COVID-19 [7]. This led to the halting of routine and elective care in the months following the outbreak [8]. Despite these efforts, the rapid spread of COVID-19 in the UK impacted health and social care nationally, and the government implemented three national lockdowns.

Given the prioritization of urgent care for COVID-19 during the pandemic on top of a stretched NHS, we hypothesised that delays in the diagnosis and/or lack of cancer screening would lead to increased short-term mortality due to more advanced stages at diagnosis. We speculated that the expected delay in diagnosis and hence potentially reduced short-term survival would be more noticeable for cancers that benefit from early detection, and for which we have successful screening programmes (such as breast and colorectal cancer).

The overall aim of this study was to investigate the impact of the COVID-19 pandemic and its management on short-term cancer survival in the UK. Using pseudonymized UK NHS records we examined: 1) secular trends in the incidence of several common cancers from 2000 to 2019 compared to 2020 to 2021; and 2) the short-term survival following diagnosis of the same cancers from 2000 to 2019 and compared to those diagnosed in 2020 to 2021.

## Methods

### Study Participants

All patients were required to be aged 18 years or older and have at least one year of prior history within the database and information on age and sex available, excluding individuals diagnosed with the same cancer any time in clinical history. For the incidence rate estimations, the study cohort consisted of individuals present in the database from 1st January 2000 or their first day of eligibility (whichever occurred first). These individuals were followed up to whichever came first: the cancer outcome of interest, date if death, exit from the database, or the 31st of December 2021 (the end of study period). For the survival analysis, only individuals with a newly diagnosed cancer were included. These individuals were followed up from the date of their diagnosis to either date of death, exit from the database, or end of the study period. For the breast cancer outcome, only females were included and for the prostate cancer outcome, only males were included in the study whereas for all other outcomes both sexes were included.

### Study design

This was a population-based cohort study using routinely collected primary care data from the Clinical Practice Research Datalink (CPRD) GOLD database (July 2022 Version 2022.07.001) in the United Kingdom (UK). This study uses data provided by patients and collected by the NHS as part of their care and support.

### Procedures

People with a diagnosis of a cancer of interest and a denominator cohort were identified from CPRD GOLD to estimate cancer incidence and overall survival. CPRD GOLD contains pseudonymised patient-level information on demographics, lifestyle data, clinical diagnoses, prescriptions and preventive care contributed by general practitioners (GP) from the UK. It is an established primary care database broadly representative of the UK population [9]. Patient-level data used in this study was obtained through an approved application to the CPRD (application number 22_001843). This database was mapped to the Observational Medical Outcomes Partnership (OMOP) Common Data Model (CDM) [10].

### Outcomes

We used SNOMED CT diagnostic codes to identify incident cancer events for the nine cancers. Diagnostic codes indicative of either non-malignant cancer or metastasis were excluded as well as diagnosis code indicative of melanoma and lymphoma occurring in the organs/sites of interest. The cancer outcome definitions were reviewed with the aid of the CohortDiagnostics R package [11]. This package was used to identify additional codes of interest and to remove those highlighted as irrelevant based on feedback from clinicians with oncology expertise through an iterative process. The clinical codelists used to define all cancer outcomes can be found in Supplementary Table S1. Additionally, a detailed description of all cancer outcomes is also provided at https://dpa-pde-oxford.shinyapps.io/EHDENCancerIncPrevCohortDiagShiny/. For survival analysis, mortality was defined as all-cause mortality based on date of death records. Mortality data in CPRD GOLD has been previously validated and shown to be over 98% accurate [12].

### Statistical Analysis

A population-based cohort study was conducted to examine the incidence and survival of nine cancers in patients aged 18 years and over registered with CPRD GOLD between January 2000 and December 2021.

The population characteristics of patients with a diagnosis of each cancer were summarised overall and separately for each calendar year strata, with median and interquartile range (IQR) used for continuous variables and counts and percentages used for categorical variables.

Annual crude incidence rates (IR) were calculated for all cancer outcomes from 2000 to 2021. For incidence, the number of events, the observed time at risk, and the IR per 100,000 person years were summarised along with 95% confidence intervals. Annual IRs were calculated as the number of incident cancer cases as the numerator and the exact recorded number of person-years in the general population within that year as the denominator.

Using the crude IRs, age-standardized IRs were calculated using the 2013 European Standard Population ESP2013 [13]. The ESP2013 serves as a standard population with a predefined age distribution where results are adjusted to match this distribution and account for differences in age structures between different populations to ensure fair comparisons. The ESP2013 provides predefined age distribution in five-year age bands; therefore, we collapsed these to obtain distributions for 10-year age bands used in this study. We used the age distribution of 20 to 29 years from ESP2013 for age-standardization as age distributions were not available for 18 to 29 years age band used in this study.

For survival analysis, we stratified by calendar time of cancer diagnosis (2000 to 2004, 2005 to 2009, 2010 to 2014, 2015 to 2019, and 2020 to 2021). To estimate short-term survival at one and two years, we used the Kaplan-Meier (KM) method. Any patients whose death date and cancer diagnosis date occurred on the same date were removed from the survival analysis (0.48-3.64% of patients depending on cancer). To avoid re-identification, we do not report results with less than five cases.

### Code availability

All code used for these analyses is publicly available online (https://github.com/oxford-pharmacoepi/EHDENCancerIncidencePrevalence). Analyses were carried out using R (version 4.2.3).

### Patient and public involvement

No patients or members of the public were involved in the design, analysis or interpretation of this study or the reported data because the study aims to examine population-level trends and patterns rather than individual experiences or perspectives.

## Results

There were 12,254,874 eligible patients 18 years and older, with at least one year of prior history identified from January 2000 to December 2021 from CPRD GOLD. The attrition table for this study for each cancer can be found in Supplementary Table S2. A summary of patient characteristics of those with a diagnosis of the different cancers is shown in Table 1.

**Table 1:** Baseline characteristics of patients at the time of cancer diagnosis for this study from 2000 to 2021.

|  | **Breast** | **Colorectal** | **Liver** | **Stomach** | **Prostate** | **Pancreatic** | **Head and Neck** | **Lung** | **Oesophageal** |
| --- | --- | --- | --- | --- | --- | --- | --- | --- | --- |
| **N** | 85400 | 53797 | 3999 | 7333 | 64925 | 10116 | 12455 | 45563 | 15170 |
| **Sex: Male (N[%])** | 0 (0%) | 29800 (55.4%) | 2848 (71.2%) | 4601 (62.7%) | 64925 (100.0%) | 5035 (49.8%) | 8614 (69.2%) | 24569 (53.9%) | 10348 (68.2%) |
| **Age (Median [IQR])** | 63 (52 to 73) | 72 (63 to 80) | 71 (63 to 78) | 75 (67 to 82) | 72 (65 to 78) | 73 (64 to 80) | 64 (56 to 73) | 72 (65 to 79) | 72 (63 to 79) |
| **Age Groups N (%)** |  |  |  |  |  |  |  |  |  |
| **18 to 29** | 278 (0.3%) | 123 (0.2%) | 14 (0.4%) | 20 (0.3%) | 6 (0.0%) | 6 (0.1%) | 95 (0.8%) | 24 (0.1%) | 12 (0.1%) |
| **30 to 39** | 2860 (3.3%) | 524 (1.0%) | 27 (0.7%) | 69 (0.9%) | 11 (0.0%) | 35 (0.3%) | 258 (2.1%) | 129 (0.3%) | 62 (0.4%) |
| **40 to 49** | 11298 (13.2%) | 2001 (3.7%) | 138 (3.5%) | 232 (3.2%) | 448 (0.7%) | 314 (3.1%) | 1071 (8.6%) | 1030 (2.3%) | 471 (3.1%) |
| **50 to 59** | 21496 (25.2%) | 6367 (11.8%) | 560 (14.0%) | 612 (8.3%) | 5841 (9.0%) | 1136 (11.2%) | 3001 (24.1%) | 4660 (10.2%) | 1961 (12.9%) |
| **60 to 69** | 22273 (26.1%) | 13466 (25.0%) | 1071 (26.8%) | 1363 (18.6%) | 20168 (31.1%) | 2546 (25.2%) | 3730 (29.9%) | 12249 (26.9%) | 4019 (26.5%) |
| **70 to 79** | 15090 (17.7%) | 17286 (32.1%) | 1337 (33.4%) | 2597 (35.4%) | 25025 (38.5%) | 3316 (32.8%) | 2790 (22.4%) | 16745 (36.8%) | 4856 (32.0%) |
| **80 to 89** | 9665 (11.3%) | 12118 (22.5%) | 783 (19.6%) | 2080 (28.4%) | 11916 (18.4%) | 2337 (23.1%) | 1293 (10.4%) | 9546 (21.0%) | 3206 (21.1%) |
| **90+** | 2440 (2.9%) | 1912 (3.6%) | 69 (1.7%) | 360 (4.9%) | 1510 (2.3%) | 426 (4.2%) | 217 (1.7%) | 1180 (2.6%) | 583 (3.8%) |
| **Median days of prior history (IQR)** | 3189 (1,637 to 4,992) | 3493 (1,892 to 5,250) | 3956 (2,141 to 5,642) | 3352 (1,813 to 5,044) | 3632 (1,943 to 5,369) | 3776 (2,103 to 5,489) | 3368 (1,818 to 5,119) | 3660 (2,009 to 5,352) | 3536 (1,932 to 5,310) |
| **Atrial fibrillation** | 2797 (3.3%) | 3747 (7.0%) | 298 (7.5%) | 560 (7.6%) | 4448 (6.9%) | 645 (6.4%) | 573 (4.6%) | 3207 (7.0%) | 1009 (6.7%) |
| **Heart failure** | 1380 (1.6%) | 1796 (3.3%) | 166 (4.2%) | 337 (4.6%) | 1964 (3.0%) | 321 (3.2%) | 255 (2.0%) | 2006 (4.4%) | 568 (3.7%) |
| **Ischemic heart disease** | 3878 (4.5%) | 5495 (10.2%) | 525 (13.1%) | 1002 (13.7%) | 7710 (11.9%) | 1106 (10.9%) | 1029 (8.3%) | 6237 (13.7%) | 1706 (11.2%) |
| **Cerebrovascular disease** | 2676 (3.1%) | 3288 (6.1%) | 265 (6.6%) | 562 (7.7%) | 3867 (6.0%) | 649 (6.4%) | 727 (5.8%) | 3840 (8.4%) | 968 (6.4%) |
| **Hyperlipidaemia** | 5865 (6.9%) | 5009 (9.3%) | 308 (7.7%) | 635 (8.7%) | 6720 (10.4%) | 1005 (9.9%) | 996 (8.0%) | 4586 (10.1%) | 1375 (9.1%) |
| **Hypertensive disorder** | 17772 (20.8%) | 14932 (27.8%) | 1199 (30.0%) | 2016 (27.5%) | 19094 (29.4%) | 2867 (28.3%) | 2966 (23.8%) | 12404 (27.2%) | 4201 (27.7%) |
| **Pulmonary embolism** | 582 (0.7%) | 724 (1.3%) | 52 (1.3%) | 112 (1.5%) | 641 (1.0%) | 174 (1.7%) | 86 (0.7%) | 804 (1.8%) | 231 (1.5%) |
| **Venous Thrombosis** | 3485 (4.1%) | 2848 (5.3%) | 215 (5.4%) | 396 (5.4%) | 3017 (4.6%) | 670 (6.6%) | 451 (3.6%) | 2331 (5.1%) | 789 (5.2%) |
| **Type 2 diabetes** | 5083 (6.0%) | 5915 (11.0%) | 1066 (26.7%) | 879 (12.0%) | 6154 (9.5%) | 2065 (20.4%) | 993 (8.0%) | 4819 (10.6%) | 1632 (10.8%) |
| **Chronic liver disease** | 216 (0.3%) | 201 (0.4%) | 839 (21.0%) | 22 (0.3%) | 140 (0.2%) | 43 (0.4%) | 178 (1.4%) | 247 (0.5%) | 92 (0.6%) |
| **Renal impairment** | 6576 (7.7%) | 6891 (12.8%) | 642 (16.1%) | 1048 (14.3%) | 7712 (11.9%) | 1399 (13.8%) | 1067 (8.6%) | 6156 (13.5%) | 1932 (12.7%) |
| **COPD** | 2751 (3.2%) | 3365 (6.3%) | 361 (9.0%) | 662 (9.0%) | 4103 (6.3%) | 743 (7.3%) | 1217 (9.8%) | 11163 (24.5%) | 1334 (8.8%) |
| **Gastrointestinal haemorrhage** | 4403 (5.2%) | 10245 (19.0%) | 454 (11.4%) | 732 (10.0%) | 4871 (7.5%) | 775 (7.7%) | 797 (6.4%) | 3113 (6.8%) | 1156 (7.6%) |
| **Osteoarthritis** | 14817 (17.4%) | 10583 (19.7%) | 901 (22.5%) | 1639 (22.4%) | 13116 (20.2%) | 2346 (23.2%) | 1938 (15.6%) | 9841 (21.6%) | 3019 (19.9%) |
| **Depressive disorder** | 12862 (15.1%) | 5352 (9.9%) | 527 (13.2%) | 775 (10.6%) | 4915 (7.6%) | 1229 (12.1%) | 1655 (13.3%) | 6397 (14.0%) | 1567 (10.3%) |
| **Dementia** | 1082 (1.3%) | 774 (1.4%) | 56 (1.4%) | 137 (1.9%) | 647 (1.0%) | 167 (1.7%) | 163 (1.3%) | 772 (1.7%) | 220 (1.5%) |

From Table 1, breast cancer was the most common cancer followed by prostate, colorectal and lung. Males were more likely to have a cancer diagnosis apart from breast cancer and for pancreatic cancer where there were no sex differences in numbers diagnosed. Cancer diagnoses were more common with increasing age peaking in those aged 70 to 79 years of age for all cancers apart from those diagnosed with breast or head and neck where more diagnoses were in those aged 60 to 69 years of age. For breast, prostate and head and neck cancers, these patients had the lowest percentages of comorbidities whereas those with a diagnosis of liver, stomach and lung cancers had the highest percentages of comorbidities.

Comparing patient characteristics of those diagnosed in 2020 to 2021 with the other calendar year groups (2000 to 2004, 2005 to 2009, 2010 to 2014, 2015 to 2019) showed similar socio-demographics with very similar age and sex distributions regardless of the year of diagnosis. Regarding comorbidities, atrial fibrillation was more common in those diagnosed in 2020 to 2021 compared to people diagnosed with similar cancers in previous years. Detailed patient characteristics stratified by calendar year of diagnosis in five-year strata can be found in Supplementary Tables S3.1-3.9.

Regarding secular trends in the incidence of cancer, annual age-standardised IRs increased from 2000 up to 2019 for all cancers except breast, colorectal, oesophageal, and stomach cancer (Figure 1). Apart from pancreatic cancer, IRs decreased for all cancers in 2020 and recovered to different extents in 2021. For breast, colorectal, oesophageal and pancreatic cancers, IRs in 2021 were higher compared to 2019. Whereas for head and neck, liver, lung, prostate and stomach cancers IRs were still lower in 2021 compared to before the pandemic in 2019. Results showing the crude annualised IRs can be found in Supplementary Figure S4.

**Figure 1:**
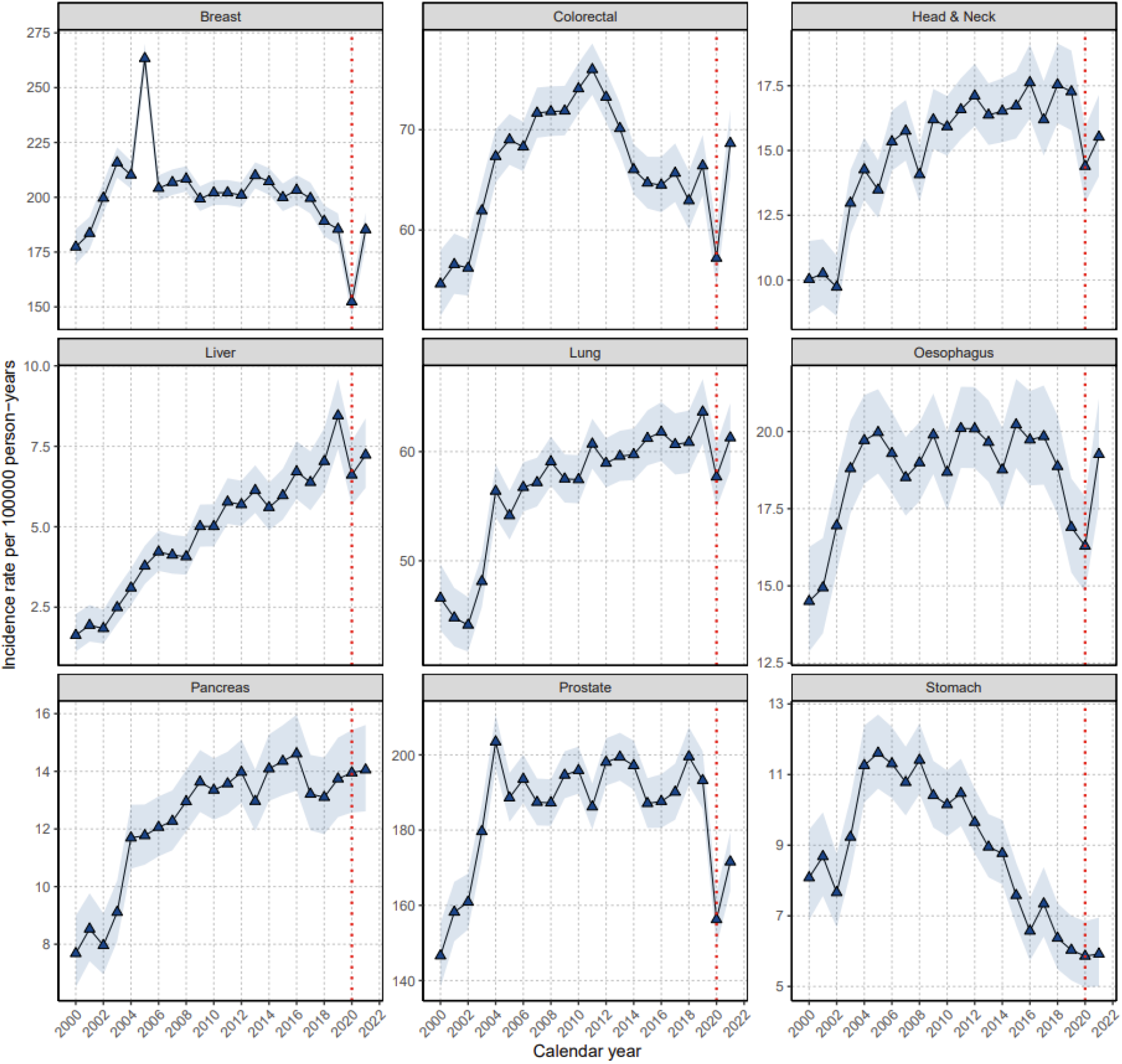
Age-standardised annual incidence rates (per 100000 person years) for nine cancers from 2000 to 2021 (red line indicating start of covid pandemic in 2020).

Positive trends in the survival of most cancers were observed in the years 2000 to 2019. Improvements in one- and two-year survival were more obvious for liver, lung, and prostate cancer (one-year survival increased from 34.6% to 47% for liver cancer; from 33.4% to 45.2% for lung cancer; and from 90.7% to 94.9% for prostate cancer from 2000 to 2019) and minimal or non-significant for head and neck and pancreatic cancers (one-year survival increased from 81.1% to 81.2% for head and neck cancer and from 23.3% to 28.1% for pancreatic cancer from 2000 to 2019) (Table 2).

**Table 2.** Survival probabilities for all cancer outcomes at one and two years after diagnosis stratified by calendar year group.

| Cancer | Calendar years | % One year survival<br>(95% CI) | % Two-year survival<br>(95% CI) |
| --- | --- | --- | --- |
| Breast | 2000 to 2004 | 93.7 (93.3 to 94.1) | 88.4 (87.8 to 89.0) |
|  | 2005 to 2009 | 95.2 (94.9 to 95.5) | 90.8 (90.4 to 91.2) |
|  | 2010 to 2014 | 95.6 (95.3 to 95.9) | 91.1 (90.7 to 91.5) |
|  | 2015 to 2019 | 95.8 (95.5 to 96.2) | 92.1 (91.6 to 92.6) |
|  | 2020 to 2021 | 95.4 (94.6 to 96.2) | 91.8 (90.4 to 93.2) |
| Colorectal | 2000 to 2004 | 76.8 (75.8 to 77.8) | 66.0 (64.8 to 67.3) |
|  | 2005 to 2009 | 77.7 (77.0 to 78.4) | 66.2 (65.4 to 67.1) |
|  | 2010 to 2014 | 79.5 (78.9 to 80.2) | 69.1 (68.3 to 69.9) |
|  | 2015 to 2019 | 79.3 (78.5 to 80.1) | 69.1 (68.1 to 70.1) |
|  | 2020 to 2021 | 76.3 (74.6 to 78.1) | 65.8 (62.5 to 69.2) |
| Head & Neck | 2000 to 2004 | 81.1 (79.1 to 83.2) | 71.7 (69.1 to 74.3) |
|  | 2005 to 2009 | 79.6 (78.1 to 81.1) | 69.0 (67.3 to 70.8) |
|  | 2010 to 2014 | 81.9 (80.6 to 83.3) | 71.6 (70.0 to 73.3) |
|  | 2015 to 2019 | 81.2 (79.6 to 82.8) | 71.7 (69.7 to 73.6) |
|  | 2020 to 2021 | 78.8 (75.3 to 82.4) | 65.1 (54.8 to 77.4) |
| Liver | 2000 to 2004 | 34.6 (29.0 to 41.2) | 22.5 (17.1 to 29.7) |
|  | 2005 to 2009 | 34.0 (30.8 to 37.5) | 19.1 (16.2 to 22.5) |
|  | 2010 to 2014 | 43.4 (40.6 to 46.5) | 28.8 (26.0 to 31.9) |
|  | 2015 to 2019 | 47.0 (43.8 to 50.3) | 31.4 (28.2 to 35.0) |
|  | 2020 to 2021 | 42.7 (36.8 to 49.4) | 31.6 (25.4 to 39.4) |
| Lung | 2000 to 2004 | 33.4 (32.1 to 34.8) | 19.2 (18.0 to 20.5) |
|  | 2005 to 2009 | 35.3 (34.4 to 36.2) | 19.8 (18.9 to 20.6) |
|  | 2010 to 2014 | 39.6 (38.7 to 40.5) | 24.0 (23.2 to 24.9) |
|  | 2015 to 2019 | 45.2 (44.1 to 46.3) | 30.4 (29.3 to 31.5) |
|  | 2020 to 2021 | 44.1 (42.1 to 46.3) | 31.2 (28.5 to 34.1) |
| Oesophagus | 2000 to 2004 | 41.2 (39.0 to 43.5) | 23.2 (21.2 to 25.5) |
|  | 2005 to 2009 | 42.0 (40.4 to 43.6) | 23.5 (22.0 to 25.1) |
|  | 2010 to 2014 | 46.1 (44.5 to 47.7) | 27.9 (26.3 to 29.5) |
|  | 2015 to 2019 | 45.2 (43.3 to 47.2) | 26.2 (24.4 to 28.1) |
|  | 2020 to 2021 | 40.9 (37.1 to 45.2) | 25.6 (20.6 to 31.9) |
| Pancreas | 2000 to 2004 | 23.3 (20.7 to 26.2) | 11.7 (9.5 to 14.4) |
|  | 2005 to 2009 | 23.0 (21.4 to 24.9) | 11.6 (10.2 to 13.2) |
|  | 2010 to 2014 | 25.2 (23.5 to 26.9) | 12.5 (11.1 to 14.1) |
|  | 2015 to 2019 | 28.1 (26.1 to 30.3) | 14.3 (12.6 to 16.3) |
|  | 2020 to 2021 | 25.1 (21.4 to 29.3) | 14.3 (9.9 to 20.6) |
| Prostate | 2000 to 2004 | 90.7 (90.1 to 91.3) | 81.6 (80.6 to 82.5) |
|  | 2005 to 2009 | 92.4 (92.0 to 92.9) | 85.3 (84.7 to 85.9) |
|  | 2010 to 2014 | 94.4 (94.1 to 94.8) | 88.7 (88.1 to 89.2) |
|  | 2015 to 2019 | 94.9 (94.5 to 95.3) | 89.1 (88.5 to 89.7) |
|  | 2020 to 2021 | 94.5 (93.6 to 95.4) | 90.5 (89.0 to 92.0) |
| Stomach | 2000 to 2004 | 41.8 (38.8 to 45.0) | 27.7 (24.8 to 31.0) |
|  | 2005 to 2009 | 40.1 (38.0 to 42.2) | 24.6 (22.7 to 26.7) |
|  | 2010 to 2014 | 44.5 (42.2 to 46.8) | 28.4 (26.3 to 30.7) |
|  | 2015 to 2019 | 47.9 (44.8 to 51.2) | 32.9 (29.8 to 36.4) |
|  | 2020 to 2021 | 45.5 (39.1 to 52.9) | 31.2 (22.0 to 44.2) |

In contrast, the first two years of the pandemic (2020 to 2021) saw a decline in short-term (one-year) survival for most cancers (Table 2). More specifically, one-year survival dropped significantly after a colorectal cancer diagnosis, from 79.3% (95%CI 78.5% to 80.1%) in 2015 to 2019 to 76.3% (74.6% to 78.1%) in 2020 to 2021. A decrease in one-year survival was also observed for many other cancers: from 81.2% (79.6% to 82.8%) to 78.8% (75.3% to 82.4%) for head and neck cancer; from 47% (43.8% to 50.3%) to 42.7% (36.8% to 49.4%) for liver cancer; from 45.2% (43.3% to 47.2%) to 40.9% (37.1% to 45.2%) for oesophageal cancer; from 28.1% (26.1% to 30.3%) to 25.1% (21.4% to 29.3%) for pancreatic cancer; and from 47.9% (44.8% to 51.2%) to 45.5% (39.1% to 52.9%) for stomach cancer. Lung, breast, and prostate cancers were an exception, with no observable decline in one-year survival in 2020 to 2021.

Survival after two years since diagnosis also indicated that those diagnosed during 2020 to 2021 had lower survival for some cancers (Table 2), however sample sizes were small. We observed slight decreases in two-year survival for those diagnosed in 2015 to 2019 compared to those diagnosed in 2020 to 2021 for many cancers: from 92.1% (91.6 to 92.6) to 91.8% (90.4 to 93.2) for breast cancer; from 69.1% (68.1% to 70.1%) to 65.8% (62.5% to 69.2%) for colorectal cancer; from 71.7% (69.7% to 73.6%) to 65.1% (54.8% to 77.4%) for head and neck cancer; from 26.2% (24.4% to 28.1%) to 25.6% (20.6% to 31.9%) for oesophageal cancer; from 14.3% (12.6% to 16.3%) to 14.3% (9.9% to 20.6%) for pancreatic cancer; and from 32.9% (29.8% to 36.4%) to 31.2% (22.0% to 44.2%) for stomach cancer. Two-year survivals of lung, liver, and prostate cancers remained unchanged when comparing those diagnosed in 2015 to 2019 and those diagnosed in 2020 to 2021.

The KM survival curves for all 9 cancers from the date of diagnosis to two years of follow-up are shown in Figure 2. Individual plots for each cancer can be found in Supplementary Figures S5.1-5.9. The numbers at risk, events and censoring at each of the time points in the KM plots for each cancer shown in Figure 2 is included in Supplementary Table S6.

**Figure 2:**
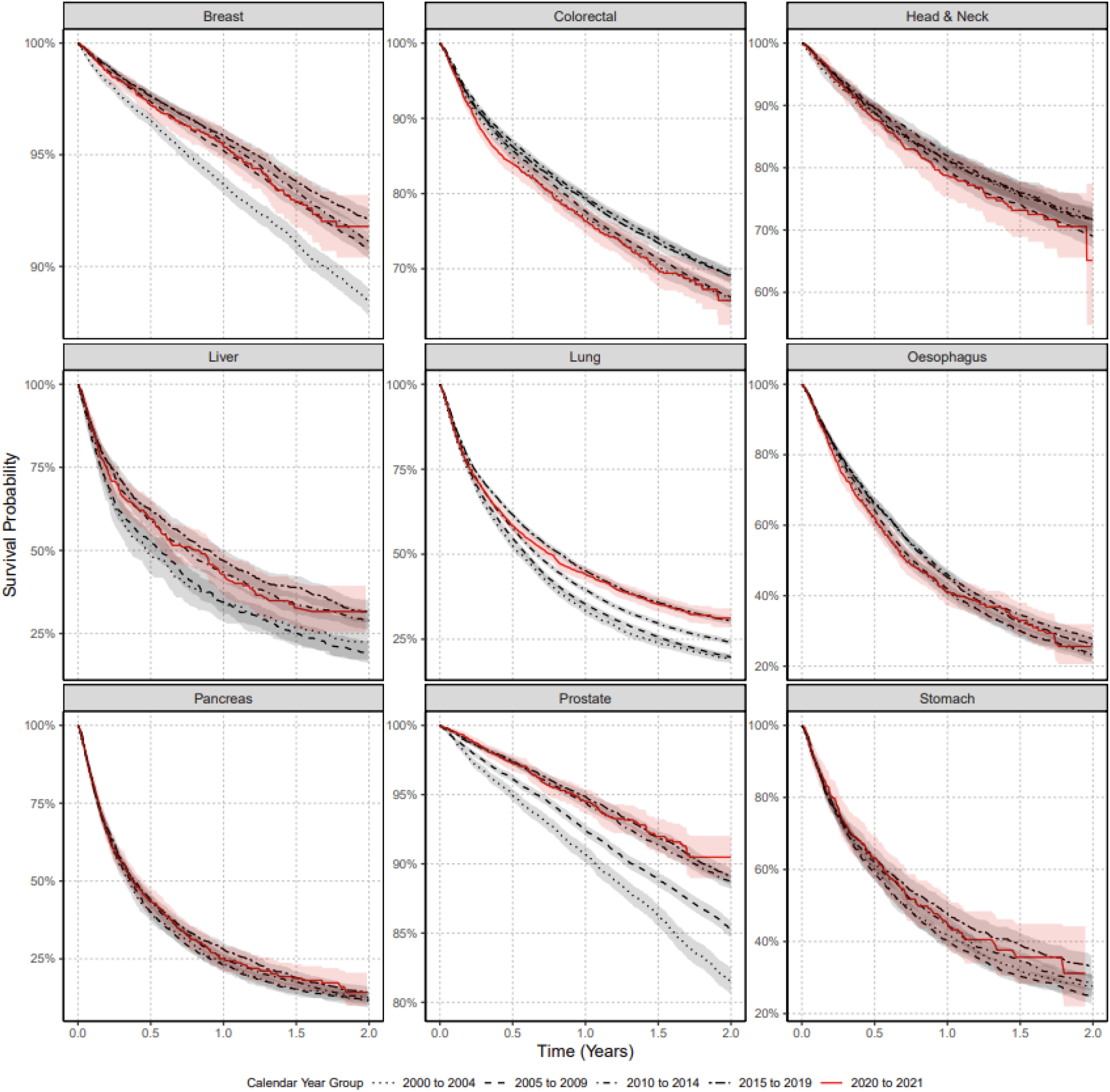
Kaplan Meier survival curves for all nine cancers stratified by calendar time of cancer diagnosis.

Cancer survival curves depicted in Figure 2 are consistent with the one- and two-year survival probabilities shown in Table 2. Survival trends tended to improve in the years 2000 to 2019 but then declined for colorectal, head and neck, oesophageal, pancreatic, and stomach cancers after their diagnosis in 2020 to 2021.

## Discussion

In this large cohort of over 12 million people, the incidence of cancer diagnoses in the UK decreased in the first year of the pandemic (2020) to then recover to different extents in 2021 for most cancers. This was more obvious for colorectal and oesophageal cancer, with rates of new diagnosis in 2021 higher than those recorded immediately before the pandemic in 2019.

One- and two-year survival after a diagnosis of cancer increased in the years 2000-2019 for most cancers, and more clearly for cancers that benefit from novel treatments including antivirals (e.g., liver cancer) and immune therapies (e.g., lung cancer). Cancers included in national screening programmes (e.g., breast, colorectal) have also improved prognosis in these same years, likely due to earlier diagnosis.

In contrast with observable improvements over the previous 20 years, our results demonstrate how the first two years of the pandemic led to a decline in short-term survival for most cancers in the UK. This decline was most pronounced for colorectal cancers, with reductions in survivorship equivalent to returning to mortality seen in the first decade of the 2000s. We found no evidence that this decrease in survivorship could be due to differences in socio-demographics or comorbidity: subjects diagnosed with cancer in 2020-2021 were of similar age and sex and had fewer comorbidities compared to those diagnosed in previous years.

In line with our results, UK cancer statistics demonstrate the increasing trend in cancer incidence over the past decades [14, 15]. Reasons for increases are multifaceted and vary per cancer type but generally include diagnostic testing (e.g., PSA for prostate cancer), public awareness of symptomology (e.g., for breast cancer), and an ageing population [14]. Unsurprisingly, the COVID-19 pandemic halted screening programmes, diagnostic appointments, and may have altered patients’ health-seeking behaviours in an attempt to contain the virus. These alterations during the pandemic led to reduced cancer diagnoses, as evidenced by our results as well as other European data [16-18]. This contrasts with data from the United States showing an increase in diagnoses of lung, colorectal, pancreatic, breast and prostate cancers in 2020 compared to 2019 [19], and illustrates the selective channelling of healthcare resources in the UK NHS during the first 2 years of the pandemic. Our data for the first time show that cancer incidence rates have largely recovered since 2020, yet diagnoses should likely exceed observed rates in order to account for the backlog in diagnoses accumulated in 2020.

One major implication of reductions in cancer incidence is that diagnoses and treatments have been delayed, affecting patient survival. This may be particularly prominent for cancers included in screening programmes, like colorectal cancers. Whilst the literature on survival estimates post-pandemic are sparse, some global estimates align with our UK-based estimates demonstrating the impact of diagnostic delays, reduction in care/availability of treatments on cancer survival, with an increase in total deaths from most cancers from 2019 to 2021 [19]. For example, data from Canada indicates that cancers for which there are organised screening programmes were most impacted by the pandemic in terms of one-year survival, including colorectal, non-Hodgkin lymphoma and uterine cancers, but not breast cancer [20]. This is in line with our findings of a more obvious and significant decrease in short-term survival seen among those newly diagnosed with colorectal cancer in 2020-2021 compared to previous years. The relatively small effect of the pandemic on breast cancer survival may be related to increased patient awareness of symptomology and surveillance of potential breast lumps at home (allowing greater detection); coupled with the fact that short-term survival is relatively much higher for breast cancer compared to other cancers.

Although cancer-related complications are often considered the primary cause of death in cancer patients, it is essential to acknowledge that individuals with cancer face an elevated risk of mortality due to direct COVID-19 infection [21]. Attributable causes of death were investigated in a small cohort study from France, demonstrating that whilst 1-year overall survival of colorectal cancer patients substantially decreased in 2020 compared to 2018-2019, these additional deaths were attributable to COVID-19 infection, not the impact on healthcare delivery or cancer severity at diagnosis [22]. Though it should be noted that some reports demonstrate no effect of the pandemic on some cancer mortality rates during 2020 compared to 2019 [23-25]. Regional differences in cancer care disruptions, particularly changes in treatment regimens and surgical delays following the pandemic, may explain these discrepancies. Indeed, at least in the Netherlands, the Dutch healthcare system successfully upheld essential care for individuals diagnosed with pancreatic cancer, with no effect of the pandemic on survival [24]. Despite these discrepancies, our results align with projections. A simulation study using cancer survival data from Public Health England and UK NHS digital estimated nearly 10,000 life-years lost over 10 years due to reductions in 2-week wait referrals [26]. Another UK study estimated a 7.9%-9.6% increase in breast cancer deaths up to 5 years post-diagnosis, with corresponding figures of 15.3%-16.6% for colorectal cancer; 4.8%-5.3% for lung cancer; and 5.8%-6% for oesophageal cancer [27]. Collectively, these estimates correspond to around 3,500 additional deaths and around 60,000 additional years of life lost. Whether the pandemic has longer-term impact on cancer survival is yet to be determined.

Surgical delays during the pandemic may account for the decreased overall survival observed for nearly all cancers. Indeed, a meta-analysis of 25 studies demonstrated that surgical delays up to 12 weeks during the pandemic decreased overall survival from breast, lung, and colon cancers, with hazard ratios of 1.46 (95%CI 1.28-1.65), 1.04 (95%CI 1.02-1.06), and 1.24 (95%CI 1.12-1.38), respectively [28].

Our study had limitations. First, we used primary care data without linkage to cancer registry, potentially leading to misclassification and delayed recording. However, the longitudinal nature and gatekeeper role of primary care in the UK NHS maximises the likelihood of complete recording, and previous validation studies have shown high accuracy and completeness of cancer diagnoses in primary care records [29]. Second, our use of primary care records precluded us from studying tumour histology, staging or cancer therapies which can all impact survival. In this study, we calculated overall survival for each cancer which does not differentiate between deaths caused by cancer vs. other causes. Therefore, it is a broad, more realistic measure of overall survival but does not provide information about the specific impact of cancer on mortality.

Our study also has strengths. The large sample size and over 20 years of follow up provided by CPRD GOLD make this a unique dataset for longitudinal analyses like the ones reported here. Cancer incidence rates before the pandemic reported here are in line with national data from Cancer Research UK statistics for all cancers, providing confidence in the validity of our estimates [15]. The high validity and completeness of mortality data, with over 98% accuracy when compared to national mortality records [12] allowed us to demonstrate the impact of the COVID-19 pandemic on short-term survival, one of the key outcomes in cancer care. Finally, the use of a representative sample of routinely collected health data increases generalisability of our results and minimises Hawthorne effects (i.e., a change in behaviour because of a patient’s awareness of being observed).

Taken together with previous research, our results show compelling evidence that short-term survival from cancer was impacted by the management of the COVID-19 pandemic in the UK. The combination of a decline in diagnoses followed by an increase in short-term mortality for many cancers and more clearly for colorectal malignancies provide compelling evidence of delays in diagnosis during the first two years of the pandemic. Yet, other factors such as reduced care / unavailability of certain cancer treatments may have contributed to reduced survival of those diagnosed with cancer during the pandemic. While more and longer-term data are needed to fully comprehend the impact of COVID-19 on cancer care, our findings illustrate the need for an immediate and intense investment from the UK NHS to resolve the current backlog in cancer screening and diagnostic procedures to improve cancer survival.

## Data Availability

This study is based in part on data from the Clinical Practice Research Datalink (CPRD) obtained under licence from the UK Medicines and Healthcare products Regulatory Agency. The data is provided by patients and collected by the NHS as part of their care and support. The interpretation and conclusions contained in this study are those of the author/s alone. Patient- level data used in this study was obtained through an approved application to the CPRD (application number 22_001843) and is only available following an approval process to safeguard the confidentiality of patient data. Details on how to apply for data access can be found at https://cprd.com/data-access.

## Contributor Statement

DPA, NLB and DN conceived and designed the study with statistical input from EHT. NLB performed the literature search. AB, AMD, AG, CG-A, FXA-J, AR, AWR and IT provided, developed and/or reviewed the clinical code lists and provided clinical expertise in this study. DN wrote the code and carried out all data analysis and visualizations for the manuscript with supervision from EB and DPA. TB contributed code for the study analysis of age-standardization. DPA, NLB and DN wrote the initial draft of the manuscript. AD and WYM mapped the CPRD GOLD data to the OMOP CDM. DN, EB, and DPA had access to the CPRD data and had final responsibility for the decision to submit for publication. DPA is the guarantor. All authors were involved in the interpretation of the results, critically reviewed the final manuscript and gave consent for publication.

## Data sharing statement

This study is based in part on data from the Clinical Practice Research Datalink (CPRD) obtained under licence from the UK Medicines and Healthcare products Regulatory Agency. The data is provided by patients and collected by the NHS as part of their care and support. The interpretation and conclusions contained in this study are those of the author/s alone. Patient-level data used in this study was obtained through an approved application to the CPRD (application number 22_001843) and is only available following an approval process to safeguard the confidentiality of patient data. Details on how to apply for data access can be found at https://cprd.com/data-access.

## Ethics Approval

The protocol for this research was approved by the independent scientific advisory committee for Medicine and Healthcare products Regulatory Agency database research (protocol number 22_001843).

## Transparency statement

The lead author (and the manuscript’s guarantor) affirms that the manuscript is an honest, accurate, and transparent account of the study being reported; that no important aspects of the study have been omitted; and that any discrepancies from the study as originally planned (and, if relevant, registered) have been explained.

