## Supplementary Material for "The impact of the COVID-19 pandemic on short-term cancer survival in the United Kingdom: a cohort analysis"

**S1: Clinical codelists for cancers**

The clinical codelists used for each cancer are listed in the table below with the corresponding SNOMED concept ID, OMOP concept ID and concept description. Only diagnosis records alone were used to identify cancer outcome for this study. We developed concept definitions using ATLAS, the OHDSI open-source platform (https://github.com/OHDSI/atlas). Clinical adjudicators reviewed the cohort definitions and associated concept sets.

| Concept Id | Concept SNOMED Code | Concept Description | Cancer |
|---|---|---|---|
| 4157332 | 372064008 | Malignant neoplasm of female breast | Breast |
| 4112853 | 254837009 | Malignant tumor of breast | Breast |
| 4091465 | 188154003 | Malignant neoplasm of upper-outer quadrant of female breast | Breast |
| 4157447 | 372095001 | Malignant neoplasm of male breast | Breast |
| 4091464 | 188147009 | Malignant neoplasm of nipple and areola of female breast | Breast |
| 4092511 | 188151006 | Malignant neoplasm of central part of female breast | Breast |
| 4092512 | 188152004 | Malignant neoplasm of upper-inner quadrant of female breast | Breast |
| 442127 | 93924008 | Primary malignant neoplasm of nipple of female breast | Breast |
| 4091466 | 188155002 | Malignant neoplasm of lower-outer quadrant of female breast | Breast |
| 4095740 | 188153009 | Malignant neoplasm of lower-inner quadrant of female breast | Breast |
| 4091467 | 188156001 | Malignant neoplasm of axillary tail of female breast | Breast |
| 433148 | 93680004 | Primary malignant neoplasm of areola of female breast | Breast |
| 4092513 | 188157005 | Malignant neoplasm, overlapping lesion of breast | Breast |
| 442126 | 93925009 | Primary malignant neoplasm of nipple of male breast | Breast |
| 436050 | 93681000 | Primary malignant neoplasm of areola of male breast | Breast |
| 4091469 | 188163001 | Malignant neoplasm of nipple and areola of male breast | Breast |
| 4091471 | 188168005 | Malignant neoplasm of ectopic site of male breast | Breast |
| 4095741 | 188159008 | Malignant neoplasm of ectopic site of female breast | Breast |
| 133711 | 109886000 | Overlapping malignant neoplasm of female breast | Breast |
| 135489 | 93884005 | Primary malignant neoplasm of male breast | Breast |
| 137809 | 93796005 | Primary malignant neoplasm of female breast | Breast |
| 432263 | 93874009 | Primary malignant neoplasm of lower inner quadrant of female breast | Breast |
| 432845 | 93745008 | Primary malignant neoplasm of central portion of female breast | Breast |
| 436353 | 94117003 | Primary malignant neoplasm of upper outer quadrant of female breast | Breast |
| 440956 | 94115006 | Primary malignant neoplasm of upper inner quadrant of female breast | Breast |
| 441513 | 372092003 | Primary malignant neoplasm of axillary tail of breast | Breast |
| 441515 | 93876006 | Primary malignant neoplasm of lower outer quadrant of female breast | Breast |
| 759932 | 1080101000119100 | Infiltrating duct carcinoma of left female breast | Breast |
| 759933 | 1080181000119100 | Infiltrating duct carcinoma of right female breast | Breast |
| 761170 | 15635761000119100 | Infiltrating duct carcinoma of bilateral female breasts | Breast |
| 765123 | 353511000119101 | Primary malignant neoplasm of female right breast | Breast |

| | | | |
|---|---|---|---|
| 4003684 | 109887009 | Overlapping malignant neoplasm of male breast | Breast |
| 4080865 | 278054005 | Infiltrating lobular carcinoma of breast | Breast |
| 4110861 | 254843006 | Familial cancer of breast | Breast |
| 4112073 | 254839007 | Scirrhous carcinoma of breast | Breast |
| 4112074 | 254840009 | Inflammatory carcinoma of breast | Breast |
| 4112854 | 254844000 | Malignant phyllodes tumor of breast | Breast |
| 4113637 | 286894008 | Carcinoma of breast - lower, inner quadrant | Breast |
| 4113638 | 286897001 | Carcinoma of breast - axillary tail | Breast |
| 4116071 | 254838004 | Carcinoma of breast | Breast |
| 4117850 | 286893002 | Carcinoma of breast - upper, inner quadrant | Breast |
| 4117851 | 286895009 | Carcinoma of breast - upper, outer quadrant | Breast |
| 4117852 | 286896005 | Carcinoma breast - lower, outer quadrant | Breast |
| 4142116 | 427685000 | HER2-positive carcinoma of breast | Breast |
| 4155292 | 372094002 | Malignant neoplasm of axillary tail of breast | Breast |
| 4157448 | 372096000 | Carcinoma of male breast | Breast |
| 4158563 | 373089009 | Primary malignant neoplasm of breast upper inner quadrant | Breast |
| 4160780 | 373083005 | Malignant neoplasm of breast upper outer quadrant | Breast |
| 4162253 | 372137005 | Primary malignant neoplasm of breast | Breast |
| 4187848 | 373081007 | Malignant neoplasm of breast lower outer quadrant | Breast |
| 4187849 | 373082000 | Malignant neoplasm of breast upper inner quadrant | Breast |
| 4187850 | 373088001 | Primary malignant neoplasm of breast upper outer quadrant | Breast |
| 4187851 | 373090000 | Primary malignant neoplasm of breast lower inner quadrant | Breast |
| 4188544 | 373080008 | Malignant neoplasm of breast lower inner quadrant | Breast |
| 4188545 | 373091001 | Primary malignant neoplasm of breast lower outer quadrant | Breast |
| 4216891 | 417181009 | Hormone receptor positive malignant neoplasm of breast | Breast |
| 4237178 | 408643008 | Infiltrating duct carcinoma of breast | Breast |
| 4246036 | 93776002 | Primary malignant neoplasm of ectopic female breast tissue | Breast |
| 4246810 | 93777006 | Primary malignant neoplasm of ectopic male breast tissue | Breast |
| 4330242 | 431396003 | Human epidermal growth factor 2 negative carcinoma of breast | Breast |
| 35622134 | 763479005 | Metaplastic carcinoma of breast | Breast |
| 35624616 | 767444009 | Germline BRCA-mutated, HER2-negative metastatic breast cancer | Breast |
| 36684817 | 353421000119109 | Primary malignant neoplasm of axillary tail of left female breast | Breast |
| 36684818 | 353431000119107 | Primary malignant neoplasm of female left breast | Breast |
| 36684819 | 353441000119103 | Primary malignant neoplasm of central portion of female left breast | Breast |
| 36684820 | 353501000119104 | Primary malignant neoplasm of axillary tail of right female breast | Breast |
| 36684821 | 353521000119108 | Primary malignant neoplasm of central portion of female right breast | Breast |
| 36684948 | 459391000124109 | Metastatic human epidermal growth factor 2 positive carcinoma of breast | Breast |
| 36684950 | 459411000124109 | Metastatic collecting duct carcinoma | Breast |
| 36712719 | 1080151000119100 | Infiltrating ductal carcinoma of upper inner quadrant of left female breast | Breast |
| 36712720 | 1080161000119100 | Infiltrating ductal carcinoma of upper outer quadrant of left female breast | Breast |

| | | | |
|---|---|---|---|
| 36712721 | 1080191000119100 | Infiltrating ductal carcinoma of central portion of right female breast | Breast |
| 36712722 | 1080231000119100 | Infiltrating ductal carcinoma of upper inner quadrant of right female breast | Breast |
| 36712723 | 1080241000119100 | Infiltrating ductal carcinoma of upper outer quadrant of right female breast | Breast |
| 36712724 | 1080261000119100 | Infiltrating lobular carcinoma of left female breast | Breast |
| 36712725 | 1080341000119100 | Infiltrating lobular carcinoma of right female breast | Breast |
| 36712934 | 15635801000119100 | Primary malignant neoplasm of bilateral female breasts | Breast |
| 36716497 | 722524005 | Primary invasive pleomorphic lobular carcinoma of breast | Breast |
| 36717260 | 1080111000119100 | Infiltrating ductal carcinoma of central portion of left female breast | Breast |
| 36717587 | 722832009 | Primary solid papillary carcinoma with invasion of breast | Breast |
| 37016439 | 45221000119105 | Primary invasive malignant neoplasm of female breast | Breast |
| 37017351 | 713609000 | Invasive carcinoma of breast | Breast |
| 37018660 | 96291000119105 | Primary malignant inflammatory neoplasm of female breast | Breast |
| 37208047 | 354491000119109 | Primary malignant neoplasm of left male breast | Breast |
| 37208048 | 354591000119108 | Primary malignant neoplasm of right male breast | Breast |
| 37208322 | 1080091000119100 | Infiltrating ductal carcinoma of axillary tail of left female breast | Breast |
| 37208324 | 1080121000119100 | Infiltrating ductal carcinoma of lower inner quadrant of left female breast | Breast |
| 37208325 | 1080131000119100 | Infiltrating ductal carcinoma of lower outer quadrant of left female breast | Breast |
| 37208326 | 1080171000119100 | Infiltrating ductal carcinoma of axillary tail of right female breast | Breast |
| 37208328 | 1080201000119100 | Infiltrating ductal carcinoma of lower inner quadrant of right female breast | Breast |
| 37208329 | 1080211000119100 | Infiltrating ductal carcinoma of lower outer quadrant of right female breast | Breast |
| 37310457 | 1082701000112100 | Locally advanced breast cancer | Breast |
| 40480215 | 444604002 | Mixed ductal and lobular carcinoma of breast | Breast |
| 40480651 | 444712000 | Mucinous carcinoma of breast | Breast |
| 40486563 | 447782002 | Carcinoma of female breast | Breast |
| 40492507 | 448952004 | Infiltrating duct carcinoma of female breast | Breast |
| 45768522 | 706970001 | Triple-negative breast cancer | Breast |
| 46270923 | 708921005 | Carcinoma of central portion of breast | Breast |
| 4180790 | 363406005 | Malignant tumor of colon | Colorectal |
| 443390 | 363351006 | Malignant tumor of rectum | Colorectal |
| 4110575 | 254582000 | Adenocarcinoma of rectum | Colorectal |
| 443381 | 363410008 | Malignant tumor of sigmoid colon | Colorectal |
| 443391 | 363350007 | Malignant tumor of cecum | Colorectal |
| 435754 | 363412000 | Malignant tumor of ascending colon | Colorectal |
| 4180792 | 363414004 | Malignant tumor of rectosigmoid junction | Colorectal |
| 443384 | 363408006 | Malignant tumor of transverse colon | Colorectal |
| 4116240 | 255081007 | Carcinoma of cecum | Colorectal |
| 4180791 | 363407001 | Malignant tumor of hepatic flexure | Colorectal |
| 443382 | 363409003 | Malignant tumor of descending colon | Colorectal |
| 4181344 | 363413005 | Malignant tumor of splenic flexure | Colorectal |
| 4198567 | 315058005 | HNPCC - hereditary nonpolyposis colon cancer | Colorectal |
| 4089661 | 187757001 | Malignant neoplasm, overlapping lesion of colon | Colorectal |

| | | | |
|---|---|---|---|
| 74582 | 93984006 | Primary malignant neoplasm of rectum | Colorectal |
| 79740 | 109838007 | Overlapping malignant neoplasm of colon | Colorectal |
| 197500 | 93761005 | Primary malignant neoplasm of colon | Colorectal |
| 432257 | 94105000 | Primary malignant neoplasm of transverse colon | Colorectal |
| 432837 | 371977004 | Primary malignant neoplasm of cecum | Colorectal |
| 436635 | 94006002 | Primary malignant neoplasm of sigmoid colon | Colorectal |
| 437798 | 94072004 | Primary malignant neoplasm of splenic flexure of colon | Colorectal |
| 438090 | 109839004 | Overlapping malignant neoplasm of rectum, anus and anal canal | Colorectal |
| 438699 | 93980002 | Primary malignant neoplasm of rectosigmoid junction | Colorectal |
| 438979 | 93826009 | Primary malignant neoplasm of hepatic flexure of colon | Colorectal |
| 441800 | 93771007 | Primary malignant neoplasm of descending colon | Colorectal |
| 443396 | 363510005 | Malignant tumor of large intestine | Colorectal |
| 764981 | 98981000119103 | Primary malignant neoplasm of ileocecal valve | Colorectal |
| 4115028 | 285312008 | Carcinoma of sigmoid colon | Colorectal |
| 4149847 | 269533000 | Carcinoma of colon | Colorectal |
| 4151260 | 269544008 | Carcinoma of the rectosigmoid junction | Colorectal |
| 4184850 | 413446001 | Adenocarcinoma of cecum | Colorectal |
| 4193165 | 312113007 | Carcinoma of descending colon | Colorectal |
| 4193871 | 312112002 | Carcinoma of transverse colon | Colorectal |
| 4193872 | 312115000 | Carcinoma of splenic flexure | Colorectal |
| 4200514 | 301756000 | Adenocarcinoma of sigmoid colon | Colorectal |
| 4207182 | 312111009 | Carcinoma of ascending colon | Colorectal |
| 4207183 | 312114001 | Carcinoma of hepatic flexure | Colorectal |
| 4246125 | 93854002 | Primary malignant neoplasm of large intestine | Colorectal |
| 4247719 | 93683002 | Primary malignant neoplasm of ascending colon | Colorectal |
| 4256776 | 408645001 | Adenocarcinoma of large intestine | Colorectal |
| 4307687 | 422581008 | Carcinoma of colon, stage II | Colorectal |
| 4310858 | 422375001 | Carcinoma of colon, stage III | Colorectal |
| 4312001 | 422985007 | Carcinoma of colon, stage IV | Colorectal |
| 4312240 | 425213009 | Carcinoma of colon, stage I | Colorectal |
| 4322376 | 425178004 | Adenocarcinoma of rectosigmoid junction | Colorectal |
| 36683531 | 781382000 | Malignant neoplasm of colon and/or rectum | Colorectal |
| 36713361 | 681601000119101 | Primary adenocarcinoma of ascending colon | Colorectal |
| 36715911 | 721695008 | Primary adenocarcinoma of ascending colon and right flexure | Colorectal |
| 36715912 | 721696009 | Primary adenocarcinoma of transverse colon | Colorectal |
| 36717495 | 721699002 | Primary adenocarcinoma of descending colon and splenic flexure | Colorectal |
| 37016239 | 184881000119106 | Primary adenocarcinoma of rectosigmoid junction | Colorectal |
| 37018659 | 96281000119107 | Overlapping malignant neoplasm of colon and rectum | Colorectal |
| 37208245 | 681651000119102 | Primary adenocarcinoma of descending colon | Colorectal |
| 40492939 | 448994001 | Carcinoma of upper rectum | Colorectal |
| 42537577 | 737058005 | Microsatellite instability-high colorectal cancer | Colorectal |
| 42872396 | 1701000119104 | Primary adenocarcinoma of colon | Colorectal |
| 4181343 | 363402007 | Malignant tumor of esophagus | Oesophagus |
| 4089656 | 187727005 | Malignant tumor of lower third of esophagus | Oesophagus |
| 4089658 | 187734007 | Malignant neoplasm of cardio-esophageal junction of stomach | Oesophagus |
| 4092060 | 187726001 | Malignant tumor of middle third of esophagus | Oesophagus |

| | | | |
|---|---|---|---|
| 4092059 | 187723009 | Malignant tumor of thoracic part of esophagus | Oesophagus |
| 4095316 | 187724003 | Malignant tumor of abdominal part of esophagus | Oesophagus |
| 4094855 | 187725002 | Malignant tumor of upper third of esophagus | Oesophagus |
| 4094854 | 187722004 | Malignant tumor of cervical part of esophagus | Oesophagus |
| 25748 | 109835005 | Overlapping malignant neoplasm of esophagus | Oesophagus |
| 26638 | 371984007 | Primary malignant neoplasm of esophagus | Oesophagus |
| 135476 | 372017008 | Primary malignant neoplasm of thoracic esophagus | Oesophagus |
| 193138 | 371998004 | Primary malignant neoplasm of lower third of esophagus | Oesophagus |
| 193971 | 371962007 | Primary malignant neoplasm of abdominal esophagus | Oesophagus |
| 432260 | 372023003 | Primary malignant neoplasm of upper third of esophagus | Oesophagus |
| 437805 | 371999007 | Primary malignant neoplasm of middle third of esophagus | Oesophagus |
| 441224 | 371978009 | Primary malignant neoplasm of cervical esophagus | Oesophagus |
| 4081043 | 276803003 | Adenocarcinoma of esophagus | Oesophagus |
| 4081044 | 276804009 | Squamous cell carcinoma of esophagus | Oesophagus |
| 4110568 | 254539001 | Carcinoma of thoracic part of esophagus | Oesophagus |
| 4110569 | 254543002 | Carcinoma of abdominal part of esophagus | Oesophagus |
| 4111799 | 254535007 | Carcinoma of cervical part of esophagus | Oesophagus |
| 4112605 | 254547001 | Carcinoma of upper third of esophagus | Oesophagus |
| 4112606 | 254551004 | Carcinoma of lower third of esophagus | Oesophagus |
| 4115265 | 254549003 | Carcinoma of middle third of esophagus | Oesophagus |
| 4145865 | 307216009 | Perforated carcinoma of esophagus | Oesophagus |
| 4162997 | 372138000 | Carcinoma of esophagus | Oesophagus |
| 4207131 | 440501006 | Siewert type II adenocarcinoma of esophagogastric junction | Oesophagus |
| 4208017 | 438946002 | Siewert type I adenocarcinoma of esophagogastric junction | Oesophagus |
| 4258546 | 439478008 | Siewert type III adenocarcinoma of esophagogastric junction | Oesophagus |
| 36674609 | 458581000124106 | Metastatic HER2 positive gastroesophageal junction cancer | Oesophagus |
| 36715844 | 721617001 | Primary adenocarcinoma of lower third of esophagus due to Barrett esophagus | Oesophagus |
| 36715845 | 721618006 | Primary squamous cell carcinoma of upper third of esophagus | Oesophagus |
| 36715846 | 721620009 | Primary squamous cell carcinoma of lower third of esophagus | Oesophagus |
| 36715847 | 721621008 | Primary squamous cell carcinoma of overlapping lesion of esophagus | Oesophagus |
| 36715848 | 721622001 | Primary adenocarcinoma of upper third of esophagus | Oesophagus |
| 36715849 | 721623006 | Primary adenocarcinoma of middle third of esophagus | Oesophagus |
| 36715850 | 721624000 | Primary adenocarcinoma of overlapping lesion of esophagus | Oesophagus |
| 36716505 | 722533007 | Primary malignant neoplasm of esophagogastric junction | Oesophagus |
| 36717176 | 721628002 | Primary adenocarcinoma of esophagogastric junction | Oesophagus |
| 36717489 | 721619003 | Primary squamous cell carcinoma of middle third of esophagus | Oesophagus |
| 37016162 | 128041000119107 | Primary adenocarcinoma of distal third of esophagus | Oesophagus |

| | | | |
|---|---|---|---|
| 37204513 | 783183009 | Salivary gland type carcinoma of esophagus | Oesophagus |
| 37204806 | 783704005 | Undifferentiated carcinoma of esophagus | Oesophagus |
| 40493492 | 449153001 | Adenocarcinoma of lower esophagus | Oesophagus |
| 4178962 | 363375006 | Malignant tumor of tongue | Head and Neck |
| 4178968 | 363429002 | Malignant tumor of larynx | Head and Neck |
| 4181339 | 363393007 | Malignant tumor of tonsil | Head and Neck |
| 436045 | 271323007 | Malignant neoplasm of lip, oral cavity and pharynx | Head and Neck |
| 4180784 | 363379000 | Malignant tumor of parotid gland | Head and Neck |
| 4181338 | 363392002 | Malignant tumor of oropharynx | Head and Neck |
| 4177101 | 363385007 | Malignant tumor of floor of mouth | Head and Neck |
| 4092212 | 187842004 | Malignant tumor of supraglottis | Head and Neck |
| 4113107 | 255069008 | Carcinoma of lip, oral cavity and pharynx | Head and Neck |
| 4181332 | 363376007 | Malignant tumor of base of tongue | Head and Neck |
| 35610411 | 1092831000000100 | Overlapping malignant neoplasm of mouth | Head and Neck |
| 4181342 | 363399006 | Malignant tumor of hypopharynx | Head and Neck |
| 4095312 | 187692001 | Malignant tumor of nasopharynx | Head and Neck |
| 4092211 | 187841006 | Malignant tumor of glottis | Head and Neck |
| 4180779 | 363348004 | Malignant tumor of lip | Head and Neck |
| 4177243 | 363507003 | Malignant tumor of pharynx | Head and Neck |
| 25189 | 363505006 | Malignant tumor of oral cavity | Head and Neck |
| 4181336 | 363388009 | Malignant tumor of soft palate | Head and Neck |
| 4177107 | 363422006 | Malignant tumor of nasal cavity | Head and Neck |
| 4181333 | 363378008 | Malignant tumor of major salivary gland | Head and Neck |
| 4180789 | 363401000 | Malignant tumor of pyriform fossa | Head and Neck |
| 4180783 | 363377003 | Malignant tumor of lingual tonsil | Head and Neck |
| 4180786 | 363386008 | Malignant tumor of buccal mucosa | Head and Neck |
| 4180788 | 363394001 | Malignant tumor of tonsillar fossa | Head and Neck |
| 4181347 | 363425008 | Malignant tumor of maxillary sinus | Head and Neck |
| 4178965 | 363391009 | Malignant tumor of retromolar area | Head and Neck |
| 4153887 | 269515006 | Carcinoma of lip | Head and Neck |
| 4095447 | 187846001 | Malignant neoplasm of thyroid cartilage | Head and Neck |
| 22557 | 363380002 | Malignant tumor of submandibular gland | Head and Neck |
| 4180787 | 363387004 | Malignant tumor of hard palate | Head and Neck |
| 4178963 | 363382005 | Malignant tumor of gum | Head and Neck |
| 4181349 | 363431006 | Malignant tumor of laryngeal cartilage | Head and Neck |
| 4092057 | 187697007 | Malignant tumor of pharyngeal recess | Head and Neck |
| 4089647 | 187694000 | Malignant tumor of adenoid | Head and Neck |
| 4091931 | 187681002 | Malignant neoplasm of anterior epiglottis | Head and Neck |
| 4181337 | 363389001 | Malignant tumor of uvula | Head and Neck |
| 440655 | 187637005 | Malignant neoplasm of tongue, tip and lateral border | Head and Neck |
| 140955 | 109830000 | Overlapping malignant neoplasm of floor of mouth | Head and Neck |
| 4177111 | 363430007 | Malignant tumor of subglottis | Head and Neck |
| 4178964 | 363390005 | Malignant tumor of palate | Head and Neck |
| 4090224 | 187652003 | Malignant tumor of anterior floor of mouth | Head and Neck |
| 4177104 | 363400004 | Malignant tumor of postcricoid region | Head and Neck |
| 4181335 | 363384006 | Malignant tumor of lower gingiva | Head and Neck |
| 436043 | 109823006 | Overlapping malignant neoplasm of tongue | Head and Neck |
| 437220 | 94134006 | Primary malignant neoplasm of ventral surface of tongue | Head and Neck |
| 4177102 | 363395000 | Malignant tumor of vallecula | Head and Neck |
| 4089516 | 187604008 | Malignant neoplasm of lower lip, external | Head and Neck |
| 4177109 | 363426009 | Malignant tumor of ethmoid sinus | Head and Neck |

| | | | |
|---|---|---|---|
| 4094850 | 187709007 | Malignant neoplasm of posterior pharynx | Head and Neck |
| 4095442 | 187828007 | Malignant neoplasm of nasal cavities, middle ear and accessory sinuses | Head and Neck |
| 4095446 | 187844003 | Malignant neoplasm of cricoid cartilage | Head and Neck |
| 4181334 | 363383000 | Malignant tumor of upper gingiva | Head and Neck |
| 4094720 | 187675005 | Malignant tumor of tonsillar pillar | Head and Neck |
| 4091932 | 187682009 | Malignant neoplasm of epiglottis, free border | Head and Neck |
| 4181348 | 363427000 | Malignant tumor of frontal sinus | Head and Neck |
| 40492021 | 448868009 | Malignant neoplasm of lateral wall of oropharynx | Head and Neck |
| 4089652 | 187708004 | Malignant tumor aryepiglottic fold - hypopharyngeal aspect | Head and Neck |
| 4092210 | 187831008 | Malignant tumor of nasal vestibule | Head and Neck |
| 4177110 | 363428005 | Malignant tumor of sphenoid sinus | Head and Neck |
| 4094721 | 187688008 | Malignant tumor of posterior wall of oropharynx | Head and Neck |
| 4092213 | 187843009 | Malignant neoplasm of arytenoid cartilage | Head and Neck |
| 4094722 | 187693006 | Malignant tumor of posterior wall of nasopharynx | Head and Neck |
| 40381321 | 187633009 | Malignant neoplasm of dorsal surface of tongue | Head and Neck |
| 4089646 | 187685006 | Malignant neoplasm of junctional region of epiglottis | Head and Neck |
| 4094716 | 187659007 | Malignant tumor of upper buccal sulcus | Head and Neck |
| 4180785 | 363381003 | Malignant tumor of sublingual gland | Head and Neck |
| 4089520 | 187622006 | Malignant tumor of labial mucosa | Head and Neck |
| 4089644 | 187666008 | Malignant neoplasm of junction of hard and soft palate | Head and Neck |
| 4090227 | 187660002 | Malignant tumor of lower buccal sulcus | Head and Neck |
| 4095309 | 187683004 | Malignant neoplasm of glossoepiglottic fold | Head and Neck |
| 4177098 | 363360003 | Malignant tumor of anterior two-thirds of tongue | Head and Neck |
| 4178961 | 363373004 | Malignant tumor of vermilion border of lower lip | Head and Neck |
| 35610172 | 1090271000000100 | Malignant neoplasm of ventral surface of tongue | Head and Neck |
| 4089530 | 187653008 | Malignant tumor of lateral floor of mouth | Head and Neck |
| 4225833 | 421249001 | Malignant tumor of vermilion border of lip | Head and Neck |
| 22839 | 109369002 | Overlapping malignant neoplasm of larynx | Head and Neck |
| 28356 | 109824000 | Overlapping malignant neoplasm of major salivary gland | Head and Neck |
| 140958 | 109388009 | Kaposi's sarcoma of palate | Head and Neck |
| 432558 | 109367000 | Overlapping malignant neoplasm of nasopharynx | Head and Neck |
| 4002498 | 110013004 | Overlapping malignant neoplasm of tonsil | Head and Neck |
| 4089524 | 187631006 | Malignant neoplasm of base of tongue dorsal surface | Head and Neck |
| 4089526 | 187644001 | Malignant tumor of junctional zone of tongue | Head and Neck |
| 4089649 | 187701005 | Malignant neoplasm of floor of nasopharynx | Head and Neck |
| 4090216 | 187606005 | Malignant tumor of upper labial mucosa | Head and Neck |
| 4090217 | 187614004 | Malignant tumor of frenum of lower lip | Head and Neck |
| 4090226 | 187658004 | Malignant tumor of vestibule of mouth | Head and Neck |
| 4092209 | 187830009 | Malignant neoplasm of nasal conchae | Head and Neck |
| 4092214 | 187845002 | Malignant neoplasm of cuneiform cartilage | Head and Neck |
| 4093012 | 187601000 | Malignant neoplasm of upper lip, lipstick area | Head and Neck |
| 4093141 | 187640005 | Malignant tumor of anterior two-thirds of tongue - ventral surface | Head and Neck |
| 4093646 | 187608006 | Malignant tumor of frenum of upper lip | Head and Neck |
| 4093647 | 187613005 | Malignant neoplasm of lower lip, buccal aspect | Head and Neck |
| 4093650 | 187635002 | Malignant neoplasm of midline of tongue | Head and Neck |
| 4093651 | 187641009 | Malignant tumor of frenum linguae | Head and Neck |
| 4095313 | 187700006 | Malignant tumor of anterior wall of nasopharynx | Head and Neck |

| | | | |
|---|---|---|---|
| 4110435 | 254484001 | Malignant tumor of posterior margin of nasal septum and choanae | Head and Neck |
| 4115137 | 254459004 | Malignant tumor of anterior pillar of fauces | Head and Neck |
| 4118988 | 302815008 | Malignant tumor of frenum of lip | Head and Neck |
| 4150793 | 271568003 | Malignant tumor of lower labial mucosa | Head and Neck |
| 4170451 | 275399006 | Malignant tumor of lipstick area of lip | Head and Neck |
| 4177100 | 363374005 | Malignant tumor of commissure of lip | Head and Neck |
| 4177103 | 363398003 | Malignant tumor of lateral wall of nasopharynx | Head and Neck |
| 4180782 | 363372009 | Malignant tumor of vermilion border of upper lip | Head and Neck |
| 4181486 | 363506007 | Malignant tumor of nasal sinuses | Head and Neck |
| 46271011 | 709031009 | Malignant neoplasm of superior wall of nasopharynx | Head and Neck |
| 26052 | 371995001 | Primary malignant neoplasm of larynx | Head and Neck |
| 28083 | 93961004 | Primary malignant neoplasm of pharynx | Head and Neck |
| 31509 | 372020000 | Primary malignant neoplasm of tonsil | Head and Neck |
| 132258 | 93808006 | Primary malignant neoplasm of frontal sinus | Head and Neck |
| 132565 | 372026006 | Primary malignant neoplasm of vermilion border of lower lip | Head and Neck |
| 132832 | 93835002 | Primary malignant neoplasm of inner aspect of lip | Head and Neck |
| 133710 | 109822001 | Overlapping malignant neoplasm of lip | Head and Neck |
| 133969 | 94135007 | Primary malignant neoplasm of vermilion border of lip | Head and Neck |
| 134290 | 372002009 | Primary malignant neoplasm of palate | Head and Neck |
| 134579 | 371976008 | Primary malignant neoplasm of buccal mucosa | Head and Neck |
| 135750 | 93802007 | Primary malignant neoplasm of floor of mouth | Head and Neck |
| 136639 | 94067008 | Primary malignant neoplasm of sphenoidal sinus | Head and Neck |
| 137219 | 93836001 | Primary malignant neoplasm of inner aspect of lower lip | Head and Neck |
| 137800 | 93889000 | Primary malignant neoplasm of maxillary sinus | Head and Neck |
| 138074 | 372027002 | Primary malignant neoplasm of vermilion border of upper lip | Head and Neck |
| 138351 | 93837005 | Primary malignant neoplasm of inner aspect of upper lip | Head and Neck |
| 140046 | 93787005 | Primary malignant neoplasm of ethmoidal sinus | Head and Neck |
| 140950 | 371990006 | Primary malignant neoplasm of gum | Head and Neck |
| 253977 | 109366009 | Overlapping malignant neoplasm of accessory sinuses | Head and Neck |
| 254282 | 94049001 | Primary malignant neoplasm of soft palate | Head and Neck |
| 255192 | 371981004 | Primary malignant neoplasm of commissure of lip | Head and Neck |
| 256633 | 93687001 | Primary malignant neoplasm of base of tongue | Head and Neck |
| 259748 | 93659005 | Primary malignant neoplasm of accessory sinus | Head and Neck |
| 259755 | 94075002 | Primary malignant neoplasm of subglottis | Head and Neck |
| 260336 | 93816002 | Primary malignant neoplasm of glottis | Head and Neck |
| 261514 | 94080006 | Primary malignant neoplasm of supraglottis | Head and Neck |
| 261808 | 94138009 | Primary malignant neoplasm of vestibule of mouth | Head and Neck |
| 432833 | 93933005 | Primary malignant neoplasm of oropharynx | Head and Neck |
| 433704 | 93989001 | Primary malignant neoplasm of retromolar area | Head and Neck |
| 433709 | 94102002 | Primary malignant neoplasm of tonsillar fossa | Head and Neck |
| 434285 | 94129007 | Primary malignant neoplasm of uvula | Head and Neck |
| 434289 | 93868009 | Primary malignant neoplasm of lingual tonsil | Head and Neck |
| 434577 | 93829002 | Primary malignant neoplasm of hypopharyngeal aspect of aryepiglottic fold | Head and Neck |
| 434587 | 93773005 | Primary malignant neoplasm of dorsal surface of tongue | Head and Neck |

| | | | |
|---|---|---|---|
| 434588 | 372004005 | Primary malignant neoplasm of parotid gland | Head and Neck |
| 435190 | 93978008 | Primary malignant neoplasm of pyriform sinus | Head and Neck |
| 435474 | 93968005 | Primary malignant neoplasm of posterior hypopharyngeal wall | Head and Neck |
| 435478 | 372022008 | Primary malignant neoplasm of upper gum | Head and Neck |
| 436042 | 371968006 | Primary malignant neoplasm of anterior two-thirds of tongue | Head and Neck |
| 436344 | 93970001 | Primary malignant neoplasm of posterior wall of nasopharynx | Head and Neck |
| 436352 | 109370001 | Primary malignant neoplasm of laryngeal cartilage | Head and Neck |
| 436643 | 93670003 | Primary malignant neoplasm of anterior aspect of epiglottis | Head and Neck |
| 436922 | 93967000 | Primary malignant neoplasm of postcricoid region | Head and Neck |
| 437226 | 240163000 | Malignant neoplasm of nasopharyngeal wall | Head and Neck |
| 437498 | 94101009 | Primary malignant neoplasm of tongue | Head and Neck |
| 438080 | 93674007 | Primary malignant neoplasm of anterior wall of nasopharynx | Head and Neck |
| 438360 | 94132005 | Primary malignant neoplasm of vallecula | Head and Neck |
| 438367 | 93917007 | Primary malignant neoplasm of nasal cavity | Head and Neck |
| 438691 | 109832008 | Overlapping malignant neoplasm of oropharynx | Head and Neck |
| 438692 | 93861003 | Primary malignant neoplasm of lateral wall of nasopharynx | Head and Neck |
| 438694 | 371991005 | Primary malignant neoplasm of hard palate | Head and Neck |
| 438982 | 93672006 | Primary malignant neoplasm of anterior portion of floor of mouth | Head and Neck |
| 439404 | 372001002 | Primary malignant neoplasm of oral cavity | Head and Neck |
| 439738 | 94076001 | Primary malignant neoplasm of sublingual gland | Head and Neck |
| 439739 | 93971002 | Primary malignant neoplasm of posterior wall of oropharynx | Head and Neck |
| 439745 | 109828002 | Primary malignant neoplasm of salivary gland duct | Head and Neck |
| 439746 | 93831006 | Primary malignant neoplasm of hypopharynx | Head and Neck |
| 440036 | 93848003 | Primary malignant neoplasm of junctional zone of tongue | Head and Neck |
| 440044 | 109368005 | Overlapping malignant neoplasm of hypopharynx | Head and Neck |
| 440047 | 94103007 | Primary malignant neoplasm of tonsillar pillar | Head and Neck |
| 440335 | 371997009 | Primary malignant neoplasm of lower gum | Head and Neck |
| 440344 | 93860002 | Primary malignant neoplasm of lateral portion of floor of mouth | Head and Neck |
| 440345 | 93862005 | Primary malignant neoplasm of lateral wall of oropharynx | Head and Neck |
| 441223 | 94078000 | Primary malignant neoplasm of superior wall of nasopharynx | Head and Neck |
| 444224 | 109833003 | Overlapping malignant neoplasm of lip, oral cavity and pharynx | Head and Neck |
| 761907 | 184861000119102 | Primary adenocarcinoma of nasopharynx | Head and Neck |
| 4001170 | 109831001 | Overlapping malignant neoplasm of palate | Head and Neck |
| 4081190 | 276954004 | Squamous cell carcinoma of floor of mouth | Head and Neck |
| 4084147 | 241861008 | Metastatic malignant neoplasm to nasopharynx | Head and Neck |
| 4090228 | 187661003 | Malignant tumor of upper labial sulcus | Head and Neck |
| 4090229 | 187662005 | Malignant tumor of lower labial sulcus | Head and Neck |
| 4093649 | 187634003 | Malignant tumor of anterior two-thirds of tongue - dorsal surface | Head and Neck |
| 4110417 | 254390001 | Carcinoma of vermilion border of lower lip | Head and Neck |
| 4110420 | 254408000 | Malignant tumor of anterior two-thirds of tongue - lateral margin | Head and Neck |
| 4110422 | 254425003 | Carcinoma of lower gum | Head and Neck |

| | | | |
|---|---|---|---|
| 4110424 | 254431000 | Carcinoma of lateral part of floor of mouth | Head and Neck |
| 4110425 | 254436005 | Carcinoma of uvula | Head and Neck |
| 4110428 | 254445006 | Carcinoma of lower buccal sulcus | Head and Neck |
| 4110430 | 254450000 | Carcinoma of upper labial sulcus | Head and Neck |
| 4110432 | 254465004 | Carcinoma of submandibular gland | Head and Neck |
| 4110434 | 254478004 | Malignant tumor of inferior turbinate | Head and Neck |
| 4110559 | 254509006 | Malignant tumor of anterior commissure | Head and Neck |
| 4110562 | 254513004 | Malignant tumor of posterior commissure | Head and Neck |
| 4110565 | 254517003 | Malignant tumor of suprahyoid epiglottis | Head and Neck |
| 4110566 | 254520006 | Malignant tumor of infrahyoid epiglottis | Head and Neck |
| 4110567 | 254526000 | Malignant tumor of laryngeal ventricle | Head and Neck |
| 4111021 | 255070009 | Overlapping malignant neoplasm of oral cavity and lips and salivary glands | Head and Neck |
| 4111022 | 255075004 | Malignant tumor of lateral nasal wall | Head and Neck |
| 4111645 | 254393004 | Carcinoma of frenum of lip | Head and Neck |
| 4111649 | 254412006 | Malignant tumor of tip of tongue | Head and Neck |
| 4111653 | 254424004 | Carcinoma of upper gum | Head and Neck |
| 4111654 | 254427006 | Carcinoma of anterior part of floor of mouth | Head and Neck |
| 4111775 | 254434008 | Carcinoma of hard palate | Head and Neck |
| 4111776 | 254437001 | Squamous cell carcinoma of buccal mucosa | Head and Neck |
| 4111777 | 254441002 | Carcinoma of upper buccal sulcus | Head and Neck |
| 4111783 | 254457002 | Carcinoma of retromolar area | Head and Neck |
| 4112451 | 254389005 | Carcinoma of vermilion border of upper lip | Head and Neck |
| 4112453 | 254398008 | Carcinoma of frenum of upper lip | Head and Neck |
| 4112455 | 254402004 | Carcinoma of frenum of lower lip | Head and Neck |
| 4112456 | 254404003 | Carcinoma of commissure of lip | Head and Neck |
| 4112580 | 254417000 | Carcinoma of frenum linguae | Head and Neck |
| 4112581 | 254423005 | Carcinoma of lingual tonsil | Head and Neck |
| 4112585 | 254466003 | Carcinoma of sublingual gland | Head and Neck |
| 4112593 | 254481009 | Malignant tumor of middle turbinate | Head and Neck |
| 4112598 | 254503007 | Malignant tumor of inferior surface of soft palate | Head and Neck |
| 4113108 | 255071008 | Squamous cell carcinoma of lip | Head and Neck |
| 4115130 | 254435009 | Carcinoma of soft palate | Head and Neck |
| 4115135 | 254454009 | Carcinoma of lower labial sulcus | Head and Neck |
| 4115138 | 254462001 | Carcinoma of parotid gland | Head and Neck |
| 4116235 | 255072001 | Malignant tumor of salivary gland | Head and Neck |
| 4116237 | 255074000 | Malignant tumor of nasal cavity and nasopharynx | Head and Neck |
| 4116339 | 303012000 | Malignant tumor of posterior wall of hypopharynx | Head and Neck |
| 4145095 | 307502000 | Squamous cell carcinoma of mouth | Head and Neck |
| 4149844 | 269516007 | Tongue carcinoma | Head and Neck |
| 4155169 | 371988005 | Primary malignant neoplasm of false vocal cord | Head and Neck |
| 4155170 | 371994002 | Primary malignant neoplasm of laryngeal aspect of aryepiglottic fold | Head and Neck |
| 4155171 | 371996000 | Primary malignant neoplasm of lip | Head and Neck |
| 4156114 | 371975007 | Primary malignant neoplasm of border of tongue | Head and Neck |
| 4157451 | 372103002 | Carcinoma of glottis | Head and Neck |
| 4157452 | 372104008 | Carcinoma of subglottis | Head and Neck |
| 4158473 | 271943005 | Carcinoma of base of tongue | Head and Neck |
| 4158909 | 274085008 | Tonsil carcinoma | Head and Neck |
| 4162118 | 371974006 | Malignant neoplasm of border of tongue | Head and Neck |
| 4162122 | 372030009 | Primary malignant neoplasm of vocal cord | Head and Neck |
| 4162136 | 372105009 | Carcinoma of supraglottis | Head and Neck |

| | | | |
|---|---|---|---|
| 4162861 | 372000001 | Primary malignant neoplasm of minor salivary gland | Head and Neck |
| 4162999 | 372141009 | Carcinoma of vocal cord | Head and Neck |
| 4166769 | 274084007 | Palate carcinoma | Head and Neck |
| 4168069 | 275490009 | Carcinoma of tongue base - dorsal surface | Head and Neck |
| 4169288 | 418372008 | Squamous cell carcinoma of mucous membrane of lower lip | Head and Neck |
| 4169627 | 419842002 | Squamous cell carcinoma of oral mucous membrane | Head and Neck |
| 4170450 | 275396004 | Carcinoma of anterior two-thirds of tongue - dorsal surface | Head and Neck |
| 4172359 | 276975007 | Carcinoma of larynx | Head and Neck |
| 4173799 | 275394001 | Carcinoma ventral surface of tongue | Head and Neck |
| 4173800 | 275395000 | Carcinoma anterior 2/3 tongue ventrum | Head and Neck |
| 4173801 | 275397008 | Carcinoma of midline of tongue | Head and Neck |
| 4174593 | 276952000 | Squamous cell carcinoma of tongue | Head and Neck |
| 4174595 | 276962007 | Squamous cell carcinoma of palate | Head and Neck |
| 4174910 | 276953005 | Squamous cell carcinoma of gum | Head and Neck |
| 4177237 | 363488008 | Malignant tumor of false cord | Head and Neck |
| 4178978 | 363487003 | Malignant tumor of aryepiglottic fold - laryngeal aspect | Head and Neck |
| 4180909 | 363485006 | Malignant tumor of minor salivary gland | Head and Neck |
| 4180910 | 363486007 | Malignant tumor of vocal cord | Head and Neck |
| 4237016 | 405822008 | Squamous cell carcinoma of larynx | Head and Neck |
| 4246015 | 93667002 | Primary malignant neoplasm of alveolar ridge mucosa | Head and Neck |
| 4246037 | 93788000 | Primary malignant neoplasm of eustachian tube | Head and Neck |
| 4246138 | 93936002 | Primary malignant neoplasm of palatine bone | Head and Neck |
| 4246793 | 93662008 | Primary malignant neoplasm of adenoid | Head and Neck |
| 4246922 | 93784003 | Primary malignant neoplasm of epiglottis | Head and Neck |
| 4247830 | 93857009 | Primary malignant neoplasm of laryngeal commissure | Head and Neck |
| 4247831 | 93858004 | Primary malignant neoplasm of laryngeal surface of epiglottis | Head and Neck |
| 4247836 | 93883004 | Primary malignant neoplasm of major salivary gland | Head and Neck |
| 4247843 | 93918002 | Primary malignant neoplasm of nasal concha | Head and Neck |
| 4252536 | 408648004 | Squamous cell carcinoma of epiglottis | Head and Neck |
| 4256777 | 408649007 | Squamous cell carcinoma of pharynx | Head and Neck |
| 4300554 | 403889000 | Verrucous carcinoma of oral cavity | Head and Neck |
| 4302482 | 419240004 | Squamous cell carcinoma of mucous membrane of upper lip | Head and Neck |
| 4306501 | 422541001 | Undifferentiated carcinoma of nasopharynx | Head and Neck |
| 4307152 | 423106003 | Adenocarcinoma of nasopharynx | Head and Neck |
| 4307266 | 423189008 | Adenoid cystic carcinoma of submandibular gland | Head and Neck |
| 4308149 | 423793008 | Mucoepidermoid carcinoma of parotid gland | Head and Neck |
| 4308627 | 423318000 | Adenoid cystic carcinoma of oropharynx | Head and Neck |
| 4309248 | 422691006 | Squamous cell carcinoma of nasopharynx | Head and Neck |
| 4309385 | 422758009 | Carcinoma of nasal meatus | Head and Neck |
| 4309400 | 422833009 | Adenoid cystic carcinoma of salivary gland | Head and Neck |
| 4309545 | 423464009 | Squamous cell carcinoma of oropharynx | Head and Neck |
| 4310135 | 424849005 | Primary sarcoma of tongue | Head and Neck |
| 4310565 | 423424005 | Mucoepidermoid carcinoma of submandibular gland | Head and Neck |
| 4311283 | 423691004 | Malignant neoplasm of gum and contiguous sites | Head and Neck |

| | | | |
|---|---|---|---|
| 4311287 | 423708008 | Mucoepidermoid carcinoma of salivary gland | Head and Neck |
| 4311476 | 93812000 | Primary malignant neoplasm of gingival mucosa | Head and Neck |
| 4311479 | 93830007 | Primary malignant neoplasm of hypopharyngeal aspect of interarytenoid fold | Head and Neck |
| 4311719 | 424779008 | Primary Kaposi's sarcoma of oral cavity | Head and Neck |
| 4312032 | 94139001 | Primary malignant neoplasm of vestibule of nose | Head and Neck |
| 4312691 | 94077005 | Primary malignant neoplasm of submaxillary gland | Head and Neck |
| 4312798 | 425127006 | Carcinoma ex pleomorphic adenoma of parotid gland | Head and Neck |
| 4312929 | 423038006 | Polymorphous low grade adenocarcinoma of salivary gland | Head and Neck |
| 4313754 | 423615009 | Adenoid cystic carcinoma of parotid gland | Head and Neck |
| 4314314 | 425225007 | Malignant mixed tumor of salivary gland | Head and Neck |
| 36684473 | 226521000119108 | Primary malignant neoplasm of nasopharynx | Head and Neck |
| 36715796 | 721556000 | Primary adenocarcinoma of palate | Head and Neck |
| 36715797 | 721557009 | Primary adenocarcinoma of parotid gland | Head and Neck |
| 36715799 | 721560002 | Primary adenocarcinoma of nasal cavity | Head and Neck |
| 36715835 | 721605003 | Primary squamous cell carcinoma of overlapping lesion of accessory sinuses | Head and Neck |
| 36715836 | 721606002 | Primary adenocarcinoma of overlapping lesion of accessory sinuses | Head and Neck |
| 36715837 | 721607006 | Primary undifferentiated carcinoma of oropharynx | Head and Neck |
| 36716485 | 722509001 | Primary rhabdomyosarcoma of oral cavity | Head and Neck |
| 36716486 | 722510006 | Primary rhabdomyosarcoma of pharynx | Head and Neck |
| 36716501 | 722529000 | Primary malignant epithelial neoplasm of nasopharynx | Head and Neck |
| 36716502 | 722530005 | Primary squamous cell carcinoma of pharyngeal tonsil | Head and Neck |
| 36716610 | 722672002 | Primary squamous cell carcinoma of base of tongue | Head and Neck |
| 36716941 | 723182009 | Primary squamous cell carcinoma of nasal cavity | Head and Neck |
| 36717226 | 722674001 | Primary squamous cell carcinoma of parotid gland | Head and Neck |
| 36717352 | 722673007 | Primary squamous cell carcinoma of lingual tonsil | Head and Neck |
| 37018571 | 7391000119103 | Primary adenoid cystic carcinoma of nasopharynx | Head and Neck |
| 37018880 | 242951000119108 | Primary squamous cell carcinoma of vermilion border of lip | Head and Neck |
| 37018947 | 18121000119104 | Primary squamous cell carcinoma of palatine tonsil | Head and Neck |
| 37116583 | 733343005 | Primary squamous cell carcinoma of oral cavity | Head and Neck |
| 37116584 | 733344004 | Primary squamous cell carcinoma of lip | Head and Neck |
| 37117762 | 733345003 | Primary squamous cell carcinoma of pharynx | Head and Neck |
| 37204489 | 783155007 | Malignant epithelial neoplasm of salivary gland | Head and Neck |
| 40486174 | 449223003 | Malignant neoplasm of alveolus of maxilla | Head and Neck |
| 40486201 | 449248000 | Nasopharyngeal carcinoma | Head and Neck |
| 40486208 | 449254004 | Carcinoma of pharynx | Head and Neck |
| 40486214 | 449260004 | Malignant neoplasm of alveolus dentalis | Head and Neck |
| 40487580 | 449472007 | Malignant epithelial neoplasm of alveolus dentalis | Head and Neck |
| 40488048 | 449578008 | Malignant neoplasm of alveolus of mandible | Head and Neck |
| 40488898 | 448214005 | Malignant epithelial neoplasm of oropharynx | Head and Neck |
| 40490009 | 448509007 | Transglottic malignant neoplasm of larynx | Head and Neck |
| 40490990 | 448665005 | Malignant epithelial neoplasm of hypopharynx | Head and Neck |
| 40492934 | 448990005 | Carcinoma of nasal cavity | Head and Neck |
| 40492985 | 449034009 | Malignant neoplasm of anterior and lateral floor of mouth | Head and Neck |
| 40493495 | 449156009 | Carcinoma of floor of mouth | Head and Neck |

| | | | |
|---|---|---|---|
| 42536528 | 735450006 | Primary malignant neuroepitheliomatous neoplasm of nasal cavity | Head and Neck |
| 42537751 | 737308008 | Primary adenocarcinoma of sublingual gland | Head and Neck |
| 42537752 | 737309000 | Primary adenocarcinoma of submandibular gland | Head and Neck |
| 42537753 | 737310005 | Primary squamous cell carcinoma of submandibular gland | Head and Neck |
| 42539700 | 737311009 | Primary squamous cell carcinoma of sublingual gland | Head and Neck |
| 44782582 | 698011002 | Keratinizing squamous cell carcinoma of nasopharynx | Head and Neck |
| 44783808 | 697993003 | Undifferentiated carcinoma of nasal sinus | Head and Neck |
| 44783851 | 698048006 | Undifferentiated nonkeratinizing carcinoma of nasopharynx | Head and Neck |
| 45768824 | 707337006 | Primary adenocarcinoma of accessory sinus | Head and Neck |
| 45768828 | 707342003 | Primary adenocarcinoma of ethmoidal sinus | Head and Neck |
| 45768829 | 707343008 | Primary adenocarcinoma of frontal sinus | Head and Neck |
| 45768830 | 707344002 | Primary adenocarcinoma of sphenoidal sinus | Head and Neck |
| 45768831 | 707346000 | Primary carcinoma of ethmoidal sinus | Head and Neck |
| 45768832 | 707347009 | Primary carcinoma of maxillary sinus | Head and Neck |
| 45768833 | 707349007 | Primary carcinoma of frontal sinus | Head and Neck |
| 45768836 | 707353009 | Primary squamous cell carcinoma of accessory sinus | Head and Neck |
| 45768837 | 707355002 | Primary squamous cell carcinoma of sphenoidal sinus | Head and Neck |
| 45768838 | 707356001 | Primary squamous cell carcinoma of frontal sinus | Head and Neck |
| 45768839 | 707357005 | Primary squamous cell carcinoma of laryngeal cartilage | Head and Neck |
| 45768840 | 707358000 | Primary squamous cell carcinoma of larynx | Head and Neck |
| 45768871 | 707395000 | Primary adenocarcinoma of hypopharynx | Head and Neck |
| 45768872 | 707396004 | Primary oxyphilic adenocarcinoma of oropharynx | Head and Neck |
| 45768873 | 707397008 | Primary basal cell adenocarcinoma of oropharynx | Head and Neck |
| 45768874 | 707398003 | Primary polymorphous low grade adenocarcinoma of oropharynx | Head and Neck |
| 45768875 | 707399006 | Primary papillary adenocarcinoma of oropharynx | Head and Neck |
| 45768876 | 707400004 | Primary mucinous adenocarcinoma of oropharynx | Head and Neck |
| 45768877 | 707401000 | Primary clear cell adenocarcinoma of oropharynx | Head and Neck |
| 45768878 | 707402007 | Primary adenocarcinoma of oropharynx | Head and Neck |
| 45768882 | 707406005 | Primary mucoepidermoid carcinoma of hypopharynx | Head and Neck |
| 45768893 | 707421004 | Primary undifferentiated carcinoma of larynx | Head and Neck |
| 45768894 | 707422006 | Primary spindle cell squamous cell carcinoma of larynx | Head and Neck |
| 45768895 | 707423001 | Primary basaloid carcinoma of larynx | Head and Neck |
| 45768896 | 707425008 | Primary adenoid squamous cell carcinoma of larynx | Head and Neck |
| 45768897 | 707426009 | Primary papillary squamous cell carcinoma of larynx | Head and Neck |
| 45768898 | 707427000 | Primary verrucous carcinoma of larynx | Head and Neck |
| 45768899 | 707429002 | Overlapping squamous cell carcinoma of larynx | Head and Neck |
| 45768900 | 707430007 | Overlapping squamous cell carcinoma of laryngeal cartilage | Head and Neck |
| 45768940 | 707479004 | Primary adenocarcinoma of subglottis | Head and Neck |
| 45768942 | 707482009 | Primary papillary squamous cell carcinoma of hypopharynx | Head and Neck |
| 45768943 | 707483004 | Primary undifferentiated carcinoma of hypopharynx | Head and Neck |
| 45768944 | 707484005 | Primary adenoid squamous cell carcinoma of hypopharynx | Head and Neck |

| | | | |
|---|---|---|---|
| 45768945 | 707485006 | Primary adenosquamous carcinoma of hypopharynx | Head and Neck |
| 45768946 | 707486007 | Primary basaloid carcinoma of hypopharynx | Head and Neck |
| 45768947 | 707487003 | Primary giant cell carcinoma of hypopharynx | Head and Neck |
| 45768948 | 707489000 | Primary spindle cell squamous cell carcinoma of hypopharynx | Head and Neck |
| 45768949 | 707490009 | Primary verrucous carcinoma of hypopharynx | Head and Neck |
| 45768951 | 707492001 | Primary squamous cell carcinoma of hypopharynx | Head and Neck |
| 45768977 | 707528007 | Primary squamous cell carcinoma of nasopharynx | Head and Neck |
| 45768978 | 707529004 | Overlapping squamous cell carcinoma of oropharynx | Head and Neck |
| 45768980 | 707532001 | Primary squamous cell carcinoma of posterior wall of oropharynx | Head and Neck |
| 45768982 | 707535004 | Primary squamous cell carcinoma of lateral wall of oropharynx | Head and Neck |
| 45768984 | 707537007 | Primary squamous cell carcinoma of anterior surface of epiglottis | Head and Neck |
| 45768985 | 707539005 | Primary adenoid cystic carcinoma of hypopharynx | Head and Neck |
| 45769017 | 707575007 | Primary squamous cell carcinoma of supraglottis | Head and Neck |
| 45769018 | 707576008 | Primary squamous cell carcinoma of subglottis | Head and Neck |
| 45769021 | 707580003 | Primary basaloid carcinoma of oropharynx | Head and Neck |
| 45769022 | 707581004 | Primary papillary squamous cell carcinoma of oropharynx | Head and Neck |
| 45769023 | 707582006 | Primary spindle cell squamous cell carcinoma of oropharynx | Head and Neck |
| 45769024 | 707583001 | Primary adenosquamous carcinoma of oropharynx | Head and Neck |
| 45769026 | 707585008 | Primary squamous cell carcinoma of oropharynx | Head and Neck |
| 45769027 | 707586009 | Primary myoepithelial carcinoma of oropharynx | Head and Neck |
| 45769028 | 707587000 | Primary carcinoma ex pleomorphic adenoma of oropharynx | Head and Neck |
| 45769029 | 707588005 | Primary epithelial-myoepithelial carcinoma of oropharynx | Head and Neck |
| 45769030 | 707590006 | Primary acinar cell carcinoma of oropharynx | Head and Neck |
| 45769031 | 707591005 | Primary mucoepidermoid carcinoma of oropharynx | Head and Neck |
| 45769032 | 707592003 | Primary infiltrating duct carcinoma of oropharynx | Head and Neck |
| 45769058 | 707627009 | Primary salivary gland type carcinoma of hypopharynx | Head and Neck |
| 45769059 | 707628004 | Overlapping squamous cell carcinoma of hypopharynx | Head and Neck |
| 45769087 | 707660007 | Primary giant cell carcinoma of larynx | Head and Neck |
| 45769089 | 707662004 | Primary basaloid squamous cell carcinoma of larynx | Head and Neck |
| 45769091 | 707664003 | Primary squamous cell carcinoma of glottis | Head and Neck |
| 45769108 | 707686002 | Primary squamous cell carcinoma of posterior wall of hypopharynx | Head and Neck |
| 45769121 | 707703001 | Primary squamous cell carcinoma of postcricoid region | Head and Neck |
| 45769122 | 707704007 | Primary squamous cell carcinoma of pyriform sinus | Head and Neck |
| 45769123 | 707705008 | Nonkeratinizing carcinoma of the nasopharynx | Head and Neck |
| 45771012 | 707339009 | Primary adenocarcinoma of maxillary sinus | Head and Neck |
| 45771013 | 707345001 | Primary carcinoma of accessory sinus | Head and Neck |
| 45771015 | 707354003 | Primary squamous cell carcinoma of maxillary sinus | Head and Neck |
| 45771018 | 707424007 | Primary adenosquamous cell carcinoma of larynx | Head and Neck |
| 45771025 | 707589002 | Primary cystadenocarcinoma of oropharynx | Head and Neck |
| 45771026 | 707593008 | Primary salivary gland-type tumor of oropharynx | Head and Neck |

| | | | |
|---|---|---|---|
| 45771033 | 707697002 | Primary squamous cell carcinoma of hypopharyngeal aspect of aryepiglottic fold | Head and Neck |
| 45772927 | 707348004 | Primary carcinoma of sphenoidal sinus | Head and Neck |
| 45772928 | 707360003 | Primary lymphoepithelial carcinoma of larynx | Head and Neck |
| 45772941 | 707481002 | Primary basaloid squamous cell carcinoma of hypopharynx | Head and Neck |
| 45772951 | 707579001 | Primary basaloid squamous cell carcinoma of oropharynx | Head and Neck |
| 45773563 | 707359008 | Primary squamous cell carcinoma of ethmoidal sinus | Head and Neck |
| 45773564 | 707538002 | Primary squamous cell carcinoma of vallecula | Head and Neck |
| 4001171 | 109841003 | Liver cell carcinoma | Liver |
| 4246127 | 93870000 | Malignant neoplasm of liver | Liver |
| 201519 | 95214007 | Primary malignant neoplasm of liver | Liver |
| 4094864 | 187769009 | Primary carcinoma of liver | Liver |
| 4095432 | 187767006 | Malignant neoplasm of liver and intrahepatic bile ducts | Liver |
| 4001172 | 109843000 | Hepatoblastoma | Liver |
| 4003021 | 109844006 | Angiosarcoma of liver | Liver |
| 4099699 | 253018005 | Fibrolamellar hepatocellular carcinoma | Liver |
| 4166154 | 274902006 | Combined hepatocellular carcinoma and cholangiocarcinoma | Liver |
| 4252535 | 408646000 | Adenocarcinoma of liver | Liver |
| 36674832 | 770685009 | Undifferentiated carcinoma of liver and intrahepatic biliary tract | Liver |
| 37204022 | 787091002 | Adenocarcinoma of liver and intrahepatic biliary tract | Liver |
| 37311916 | 788982002 | Mesothelial carcinoma of liver | Liver |
| 37396736 | 716648006 | Embryonal sarcoma of liver | Liver |
| 443388 | 363358000 | Malignant tumor of lung | Lung |
| 258369 | 93880001 | Primary malignant neoplasm of lung | Lung |
| 4092216 | 187862007 | Malignant neoplasm of upper lobe of lung | Lung |
| 4089756 | 187870002 | Malignant neoplasm of lower lobe of lung | Lung |
| 4151250 | 269464000 | Malignant neoplasm of upper lobe, bronchus or lung | Lung |
| 4246121 | 93827000 | Primary malignant neoplasm of hilus of lung | Lung |
| 4089754 | 187866005 | Malignant neoplasm of middle lobe of lung | Lung |
| 4092217 | 187864008 | Malignant neoplasm of middle lobe, bronchus or lung | Lung |
| 258375 | 109371002 | Overlapping malignant neoplasm of bronchus and lung | Lung |
| 4092218 | 187865009 | Malignant neoplasm of middle lobe bronchus | Lung |
| 256646 | 372112000 | Primary malignant neoplasm of middle lobe, bronchus or lung | Lung |
| 261236 | 372135002 | Primary malignant neoplasm of upper lobe, bronchus or lung | Lung |
| 433973 | 93729006 | Primary malignant neoplasm of bronchus of left lower lobe | Lung |
| 762426 | 354701000119107 | Primary malignant neoplasm of left lung | Lung |
| 762427 | 354741000119109 | Primary malignant neoplasm of right lung | Lung |
| 4110587 | 254625005 | Malignant tumor of lung parenchyma | Lung |
| 4110589 | 254629004 | Large cell carcinoma of lung | Lung |
| 4110590 | 254631008 | Giant cell carcinoma of lung | Lung |
| 4110591 | 254632001 | Small cell carcinoma of lung | Lung |
| 4110705 | 254634000 | Squamous cell carcinoma of lung | Lung |

| | | | |
|---|---|---|---|
| 4110706 | 254638002 | Pancoast tumor | Lung |
| 4111807 | 254635004 | Epithelioid hemangioendothelioma of lung | Lung |
| 4112738 | 254626006 | Adenocarcinoma of lung | Lung |
| 4112739 | 254633006 | Oat cell carcinoma of lung | Lung |
| 4115276 | 254637007 | Non-small cell lung cancer | Lung |
| 4140471 | 427038005 | Epidermal growth factor receptor negative non-small cell lung cancer | Lung |
| 4143825 | 426964009 | Epidermal growth factor receptor positive non-small cell lung cancer | Lung |
| 4155293 | 372111007 | Carcinoma of lower lobe, bronchus or lung | Lung |
| 4157454 | 372110008 | Primary malignant neoplasm of lower lobe, bronchus or lung | Lung |
| 4162248 | 372113005 | Carcinoma of middle lobe, bronchus or lung | Lung |
| 4162252 | 372136001 | Carcinoma of upper lobe, bronchus or lung | Lung |
| 4196724 | 313353007 | Squamous cell carcinoma of bronchus in left lower lobe | Lung |
| 4196725 | 313355000 | Squamous cell carcinoma of bronchus in right lower lobe | Lung |
| 4197581 | 313354001 | Squamous cell carcinoma of bronchus in left upper lobe | Lung |
| 4197582 | 313356004 | Squamous cell carcinoma of bronchus in right middle lobe | Lung |
| 4197583 | 313357008 | Squamous cell carcinoma of bronchus in right upper lobe | Lung |
| 4208307 | 440173001 | Nonsquamous nonsmall cell neoplasm of lung | Lung |
| 4246027 | 93730001 | Primary malignant neoplasm of bronchus of left upper lobe | Lung |
| 4246126 | 93865007 | Primary malignant neoplasm of left upper lobe of lung | Lung |
| 4246148 | 93991009 | Primary malignant neoplasm of right lower lobe of lung | Lung |
| 4246804 | 93732009 | Primary malignant neoplasm of bronchus of right middle lobe | Lung |
| 4246805 | 93733004 | Primary malignant neoplasm of bronchus of right upper lobe | Lung |
| 4247727 | 93731002 | Primary malignant neoplasm of bronchus of right lower lobe | Lung |
| 4247832 | 93864006 | Primary malignant neoplasm of lower lobe of left lung | Lung |
| 4307118 | 423050000 | Large cell carcinoma of lung, TNM stage 2 | Lung |
| 4308479 | 423121009 | Non-small cell carcinoma of lung, TNM stage 4 | Lung |
| 4310448 | 423295000 | Squamous cell carcinoma of lung, TNM stage 1 | Lung |
| 4310703 | 424132000 | Non-small cell carcinoma of lung, TNM stage 1 | Lung |
| 4311501 | 93992002 | Primary malignant neoplasm of right middle lobe of lung | Lung |
| 4311997 | 422968005 | Non-small cell carcinoma of lung, TNM stage 3 | Lung |
| 4312274 | 425376008 | Squamous cell carcinoma of lung, TNM stage 4 | Lung |
| 4312567 | 93993007 | Primary malignant neoplasm of upper lobe of right lung | Lung |
| 4312768 | 424970000 | Large cell carcinoma of lung, TNM stage 3 | Lung |
| 4313200 | 423468007 | Squamous cell carcinoma of lung, TNM stage 2 | Lung |
| 4313751 | 423600008 | Large cell carcinoma of lung, TNM stage 4 | Lung |
| 4314040 | 424938000 | Large cell carcinoma of lung, TNM stage 1 | Lung |
| 4314172 | 425048006 | Non-small cell carcinoma of lung, TNM stage 2 | Lung |
| 4322387 | 425230006 | Squamous cell carcinoma of lung, TNM stage 3 | Lung |
| 36686537 | 12235561000119100 | Large cell carcinoma of left lung | Lung |

| | | | |
|---|---|---|---|
| 36686538 | 12235601000119100 | Large cell carcinoma of right lung | Lung |
| 36712707 | 1078881000119100 | Primary adenocarcinoma of lower lobe of left lung | Lung |
| 36712708 | 1078901000119100 | Primary adenocarcinoma of upper lobe of left lung | Lung |
| 36712709 | 1078931000119100 | Primary adenocarcinoma of lower lobe of right lung | Lung |
| 36712815 | 12240951000119100 | Squamous cell carcinoma of left lung | Lung |
| 36712816 | 12240991000119100 | Squamous cell carcinoma of right lung | Lung |
| 36712981 | 15956381000119100 | Adenocarcinoma of right lung | Lung |
| 36713366 | 683991000119103 | Extensive stage primary small cell carcinoma of lung | Lung |
| 36716426 | 722425009 | Reactive oxygen species 1 positive non-small cell lung cancer | Lung |
| 36716500 | 722528008 | Primary malignant neuroendocrine neoplasm of lung | Lung |
| 36717017 | 1078961000119100 | Primary adenocarcinoma of upper lobe of right lung | Lung |
| 37109576 | 723301009 | Squamous non-small cell lung cancer | Lung |
| 37110031 | 724056005 | Malignant neoplasm of lower lobe of right lung | Lung |
| 37110032 | 724058006 | Malignant neoplasm of upper lobe of left lung | Lung |
| 37110033 | 724059003 | Malignant neoplasm of lower lobe of left lung | Lung |
| 37110034 | 724060008 | Malignant neoplasm of right upper lobe of lung | Lung |
| 37311684 | 822969007 | Acinar cell cystadenocarcinoma of lung | Lung |
| 37395648 | 67811000119102 | Primary small cell malignant neoplasm of lung, TNM stage 1 | Lung |
| 37395649 | 67821000119109 | Primary small cell malignant neoplasm of lung, TNM stage 2 | Lung |
| 37395650 | 67831000119107 | Primary small cell malignant neoplasm of lung, TNM stage 3 | Lung |
| 37395651 | 67841000119103 | Primary small cell malignant neoplasm of lung, TNM stage 4 | Lung |
| 40391740 | 189815007 | Pulmonary blastoma | Lung |
| 40492938 | 448993007 | Carcinoma of lung | Lung |
| 42539251 | 15956341000119100 | Adenocarcinoma of left lung | Lung |
| 45766129 | 703228009 | Non-small cell lung cancer with mutation in epidermal growth factor receptor | Lung |
| 45766131 | 703230006 | Non-small cell lung cancer without mutation in epidermal growth factor receptor | Lung |
| 45768879 | 707403002 | Primary fetal adenocarcinoma of lung | Lung |
| 45768880 | 707404008 | Primary mixed subtype adenocarcinoma of lung | Lung |
| 45768881 | 707405009 | Primary adenosquamous carcinoma of lung | Lung |
| 45768883 | 707408006 | Primary small cell non-keratinizing squamous cell carcinoma of lung | Lung |
| 45768884 | 707409003 | Primary acinar cell carcinoma of lung | Lung |
| 45768885 | 707410008 | Primary solid carcinoma of lung | Lung |
| 45768886 | 707411007 | Primary papillary adenocarcinoma of lung | Lung |
| 45768916 | 707451005 | Primary adenocarcinoma of lung | Lung |
| 45768917 | 707452003 | Primary mucinous adenocarcinoma of lung | Lung |
| 45768918 | 707453008 | Primary clear cell squamous cell carcinoma of lung | Lung |
| 45768919 | 707454002 | Primary basaloid squamous cell carcinoma of lung | Lung |
| 45768920 | 707456000 | Primary undifferentiated carcinoma of lung | Lung |
| 45768921 | 707457009 | Primary spindle cell carcinoma of lung | Lung |
| 45768922 | 707458004 | Primary pleomorphic carcinoma of lung | Lung |
| 45768923 | 707460002 | Primary pseudosarcomatous carcinoma of lung | Lung |
| 45768927 | 707464006 | Primary myoepithelial carcinoma of lung | Lung |
| 45768928 | 707466008 | Primary adenoid cystic carcinoma of lung | Lung |
| 45768929 | 707467004 | Primary salivary gland type carcinoma of lung | Lung |

| | | | |
|---|---|---|---|
| 45768930 | 707468009 | Primary mixed mucinous and non-mucinous bronchiolo-alveolar carcinoma of lung | Lung |
| 45768931 | 707469001 | Primary non-mucinous bronchiolo-alveolar carcinoma of lung | Lung |
| 45768932 | 707470000 | Primary mucinous bronchiolo-alveolar carcinoma of lung | Lung |
| 45769034 | 707595001 | Primary mucinous cystadenocarcinoma of lung | Lung |
| 45769035 | 707596000 | Primary carcinosarcoma of lung | Lung |
| 45772933 | 707407001 | Primary signet ring cell carcinoma of lung | Lung |
| 45772938 | 707455001 | Primary papillary squamous cell carcinoma of lung | Lung |
| 45772939 | 707465007 | Primary mucoepidermoid carcinoma of lung | Lung |
| 46272955 | 711414003 | Primary clear cell adenocarcinoma of lung | Lung |
| 4180793 | 363418001 | Malignant tumor of pancreas | Pancreas |
| 4178967 | 363419009 | Malignant tumor of head of pancreas | Pancreas |
| 4094866 | 187793004 | Malignant tumor of pancreatic duct | Pancreas |
| 4095436 | 187792009 | Malignant tumor of tail of pancreas | Pancreas |
| 4092072 | 187791002 | Malignant tumor of body of pancreas | Pancreas |
| 4095437 | 187794005 | Malignant tumor of Islets of Langerhans | Pancreas |
| 25486 | 93843007 | Primary malignant neoplasm of islets of Langerhans | Pancreas |
| 199754 | 372003004 | Primary malignant neoplasm of pancreas | Pancreas |
| 432843 | 94082003 | Primary malignant neoplasm of tail of pancreas | Pancreas |
| 433423 | 93939009 | Primary malignant neoplasm of pancreatic duct | Pancreas |
| 434293 | 93715005 | Primary malignant neoplasm of body of pancreas | Pancreas |
| 440649 | 372119009 | Primary malignant neoplasm of head of pancreas | Pancreas |
| 4110585 | 254612002 | Carcinoma of endocrine pancreas | Pancreas |
| 4111024 | 255088001 | Malignant tumor of exocrine pancreas | Pancreas |
| 4112734 | 254611009 | Malignant tumor of endocrine pancreas | Pancreas |
| 4157459 | 372142002 | Carcinoma of pancreas | Pancreas |
| 4178960 | 363369002 | Carcinoma of tail of pancreas | Pancreas |
| 4181331 | 363368005 | Carcinoma of body of pancreas | Pancreas |
| 4209933 | 326072005 | Carcinoma of head of pancreas | Pancreas |
| 4340498 | 235966007 | Cystadenocarcinoma of pancreas | Pancreas |
| 36683250 | 780821007 | Invasive intraductal papillary-mucinous carcinoma of pancreas | Pancreas |
| 36713362 | 681621000119105 | Primary adenocarcinoma of body of pancreas | Pancreas |
| 36713363 | 681721000119103 | Primary adenocarcinoma of head of pancreas | Pancreas |
| 37204187 | 782697005 | Solid pseudopapillary carcinoma of pancreas | Pancreas |
| 37204852 | 783771003 | Acinar cell carcinoma of pancreas | Pancreas |
| 37206235 | 785879009 | Mucinous cystadenocarcinoma of pancreas | Pancreas |
| 37311469 | 792907004 | Pancreatic ductal adenocarcinoma | Pancreas |
| 37395837 | 715414009 | Familial malignant neoplasm of pancreas | Pancreas |
| 42872399 | 1651000119109 | Primary adenocarcinoma of pancreas | Pancreas |
| 45763891 | 700423003 | Adenocarcinoma of pancreas | Pancreas |
| 4163261 | 399068003 | Malignant tumor of prostate | Prostate |
| 200962 | 93974005 | Primary malignant neoplasm of prostate | Prostate |
| 4082919 | 278060005 | Endometrioid carcinoma of prostate | Prostate |
| 4116087 | 254900004 | Carcinoma of prostate | Prostate |
| 4141960 | 427492003 | Hormone refractory prostate cancer | Prostate |
| 4161028 | 399490008 | Adenocarcinoma of prostate | Prostate |
| 4164017 | 399590005 | Squamous cell carcinoma of prostate | Prostate |
| 4288534 | 396198006 | Small cell carcinoma of prostate | Prostate |
| 36684947 | 459381000124106 | Metastatic castration-resistant prostate cancer | Prostate |
| 36716186 | 722103009 | Hormone sensitive prostate cancer | Prostate |

| | | | |
|---|---|---|---|
| 37311236 | 823017009 | Infiltrating duct carcinoma of prostate | Prostate |
| 37311683 | 822970008 | Acinar cell cystadenocarcinoma of prostate | Prostate |
| 37395835 | 715412008 | Familial prostate cancer | Prostate |
| 443387 | 363349007 | Malignant tumor of stomach | Stomach |
| 4094856 | 187732006 | Malignant tumor of cardia | Stomach |
| 4089658 | 187734007 | Malignant neoplasm of cardio-esophageal junction of stomach | Stomach |
| 4092061 | 187740000 | Malignant tumor of pyloric antrum | Stomach |
| 4095320 | 187742008 | Malignant tumor of body of stomach | Stomach |
| 4094859 | 187738005 | Malignant neoplasm of pyloric canal of stomach | Stomach |
| 4149837 | 269459004 | Malignant tumor of lesser curve of stomach | Stomach |
| 4095317 | 187736009 | Malignant tumor of pylorus | Stomach |
| 4095319 | 187741001 | Malignant tumor of fundus of stomach | Stomach |
| 4149838 | 269460009 | Malignant tumor of greater curve of stomach | Stomach |
| 35610176 | 1090291000000100 | Malignant neoplasm of prepylorus of stomach | Stomach |
| 35610248 | 1090971000000100 | Malignant neoplasm of posterior wall of stomach | Stomach |
| 35610249 | 1090981000000100 | Malignant neoplasm of anterior wall of stomach | Stomach |
| 197803 | 109836006 | Overlapping malignant neoplasm of stomach | Stomach |
| 4094857 | 187733001 | Malignant neoplasm of cardiac orifice of stomach | Stomach |
| 192255 | 93977003 | Primary malignant neoplasm of pylorus | Stomach |
| 193422 | 93717002 | Primary malignant neoplasm of body of stomach | Stomach |
| 196044 | 372014001 | Primary malignant neoplasm of stomach | Stomach |
| 432838 | 93738008 | Primary malignant neoplasm of cardia of stomach | Stomach |
| 434292 | 93867004 | Primary malignant neoplasm of lesser curvature of stomach | Stomach |
| 435751 | 93809003 | Primary malignant neoplasm of fundus of stomach | Stomach |
| 437224 | 93818001 | Primary malignant neoplasm of greater curvature of stomach | Stomach |
| 438089 | 93976007 | Primary malignant neoplasm of pyloric antrum | Stomach |
| 4110570 | 254553001 | Carcinoma of cardia | Stomach |
| 4110571 | 254555008 | Carcinoma of fundus of stomach | Stomach |
| 4110572 | 254559002 | Carcinoma of pyloric antrum | Stomach |
| 4112607 | 254561006 | Carcinoma of pylorus | Stomach |
| 4112609 | 254567005 | Carcinoma of greater curve of stomach | Stomach |
| 4115266 | 254557000 | Carcinoma of body of stomach | Stomach |
| 4115267 | 254563009 | Carcinoma of lesser curve of stomach | Stomach |
| 4155299 | 372143007 | Carcinoma of stomach | Stomach |
| 4174763 | 276809004 | Early gastric cancer | Stomach |
| 4174764 | 276810009 | Late gastric cancer | Stomach |
| 4248802 | 408647009 | Adenocarcinoma of stomach | Stomach |
| 35624157 | 766757006 | Undifferentiated carcinoma of stomach | Stomach |
| 36715854 | 721629005 | Linitis plastica of stomach | Stomach |
| 36715856 | 721633003 | Primary adenocarcinoma of overlapping lesion of stomach | Stomach |
| 36715865 | 721643000 | Primary malignant mesenchymal neoplasm of stomach | Stomach |
| 36717177 | 721632008 | Primary adenocarcinoma of pyloric antrum of stomach | Stomach |
| 36717490 | 721630000 | Primary adenocarcinoma of cardia of stomach | Stomach |
| 36717622 | 681631000119108 | Primary adenocarcinoma of body of stomach | Stomach |
| 37396688 | 716586009 | Epstein-Barr virus associated gastric carcinoma | Stomach |
| 37396884 | 716859000 | Hereditary diffuse carcinoma of stomach | Stomach |

**S2: Attrition for study populations for incidence and survival**

| Reason | Breast | Colorectal | Head and Neck | Liver | Lung | Oesophageal | Pancreatic | Prostate | Stomach |
|---|---|---|---|---|---|---|---|---|---|
| **Starting population** | 17054819 | 17054819 | 17054819 | 17054819 | 17054819 | 17054819 | 17054819 | 17054819 | 17054819 |
| **Not missing year of birth** | 17054819 | 17054819 | 17054819 | 17054819 | 17054819 | 17054819 | 17054819 | 17054819 | 17054819 |
| **Not missing sex** | 17054819 | 17054819 | 17054819 | 17054819 | 17054819 | 17054819 | 17054819 | 17054819 | 17054819 |
| **Satisfies age criteria during the study period based on year of birth** | 15210165 | 15210165 | 15210165 | 15210165 | 15210165 | 15210165 | 15210165 | 15210165 | 15210165 |
| **Observation time available during study period** | 13978229 | 13978229 | 13978229 | 13978229 | 13978229 | 13978229 | 13978229 | 13978229 | 13978229 |
| **Satisfies age criteria during the study period** | 13978229 | 13978229 | 13978229 | 13978229 | 13978229 | 13978229 | 13978229 | 13978229 | 13978229 |
| **Prior history requirement fulfilled during study period** | 12254874 | 12254874 | 12254874 | 12254874 | 12254874 | 12254874 | 12254874 | 12254874 | 12254874 |
| **Female only (Breast cancer only)** | 6275193 | - | - | - | - | - | - | - | - |
| **Males only (Prostate cancer only)** | - | - | - | - | - | - | - | 5979681 | - |
| **Observation time available after applying age and prior history criteria** | 5848436 | 11388117 | 11388117 | 11388117 | 11388117 | 11388117 | 11388117 | 5539681 | 11388117 |
| **No prior cancer events** | 5832192 | 11381775 | 11386416 | 11387938 | 11385621 | 11387173 | 11387716 | 5531493 | 11387519 |
| **With a cancer diagnosis** | 85400 | 53797 | 12455 | 3999 | 45563 | 15170 | 10116 | 64925 | 7333 |
| **Cancer diagnosis not on same day as date of death** | 84984 | 53098 | 12381 | 3892 | 43903 | 14944 | 9770 | 64614 | 7156 |

**S3: Patient characteristics for each cancer stratified by calendar year of diagnosis**

**S3.1: Patient characteristics for breast cancer stratified by calendar year of diagnosis**

| Calendar Year of Diagnosis | 2000 to 2004 | 2005 to 2009 | 2010 to 2014 | 2015 to 2019 | 2020 to 2021 |
|---|---|---|---|---|---|
| **N** | 15530 | 25622 | 23980 | 15946 | 4322 |
| **Sex: Male (N[%])** | 0 (0%) | 0 (0%) | 0 (0%) | 0 (0%) | 0 (0%) |
| **Age (Median [IQR])** | 61 (52 to 73) | 62 (52 to 72) | 63 (52 to 73) | 63 (53 to 73) | 63 (53 to 74) |
| **Age Groups N (%)** | | | | | |
| **18-29** | 45 (0.3%) | 80 (0.3%) | 85 (0.4%) | 52 (0.3%) | 16 (0.4%) |
| **30-39** | 646 (4.2%) | 849 (3.3%) | 689 (2.9%) | 507 (3.2%) | 169 (3.9%) |
| **40-49** | 2001 (12.9%) | 3328 (13.0%) | 3446 (14.4%) | 1994 (12.5%) | 529 (12.2%) |
| **50-59** | 4309 (27.7%) | 6484 (25.3%) | 5521 (23.0%) | 4093 (25.7%) | 1089 (25.2%) |
| **60-69** | 3547 (22.8%) | 7032 (27.4%) | 6554 (27.3%) | 4077 (25.6%) | 1063 (24.6%) |
| **70-79** | 2701 (17.4%) | 4353 (17.0%) | 4187 (17.5%) | 3006 (18.9%) | 843 (19.5%) |
| **80-89** | 1813 (11.7%) | 2817 (11.0%) | 2761 (11.5%) | 1779 (11.2%) | 495 (11.5%) |
| **90+** | 468 (3.0%) | 679 (2.7%) | 737 (3.1%) | 438 (2.7%) | 118 (2.7%) |
| **Median days of prior history (IQR)** | 1541 (888 to 3,001) | 2291 (1,370 to 3,502) | 3815 (2,687 to 5,002) | 5102 (3,571 to 6,131) | 6236.5 (3,576 to 6,987) |
| **Atrial fibrillation** | 369 (2.4%) | 787 (3.1%) | 863 (3.6%) | 608 (3.8%) | 170 (3.9%) |
| **Heart failure** | 327 (2.1%) | 415 (1.6%) | 355 (1.5%) | 229 (1.4%) | 54 (1.2%) |
| **Ischemic heart disease** | 778 (5.0%) | 1192 (4.7%) | 1100 (4.6%) | 648 (4.1%) | 160 (3.7%) |
| **Cerebrovascular disease** | 497 (3.2%) | 714 (2.8%) | 774 (3.2%) | 527 (3.3%) | 164 (3.8%) |
| **Hyperlipidemia** | 606 (3.9%) | 1697 (6.6%) | 1998 (8.3%) | 1296 (8.1%) | 268 (6.2%) |
| **Hypertensive disorder** | 2489 (16.0%) | 5276 (20.6%) | 5575 (23.2%) | 3508 (22.0%) | 924 (21.4%) |
| **Pulmonary embolism** | 68 (0.4%) | 151 (0.6%) | 177 (0.7%) | 137 (0.9%) | 49 (1.1%) |
| **Venous Thrombosis** | 536 (3.5%) | 876 (3.4%) | 1130 (4.7%) | 747 (4.7%) | 196 (4.5%) |
| **Type 2 diabetes** | 467 (3.0%) | 1389 (5.4%) | 1653 (6.9%) | 1219 (7.6%) | 355 (8.2%) |
| **Chronic liver disease** | 21 (0.1%) | 45 (0.2%) | 69 (0.3%) | 65 (0.4%) | 16 (0.4%) |
| **Renal impairment** | 126 (0.8%) | 1732 (6.8%) | 2728 (11.4%) | 1638 (10.3%) | 352 (8.1%) |
| **COPD** | 320 (2.1%) | 768 (3.0%) | 833 (3.5%) | 640 (4.0%) | 190 (4.4%) |
| **Gastrointestinal hemorrhage** | 571 (3.7%) | 1150 (4.5%) | 1410 (5.9%) | 1033 (6.5%) | 239 (5.5%) |
| **Osteoarthritis** | 2241 (14.4%) | 4032 (15.7%) | 4638 (19.3%) | 3101 (19.4%) | 805 (18.6%) |
| **Depressive disorder** | 1948 (12.5%) | 3230 (12.6%) | 4013 (16.7%) | 2915 (18.3%) | 756 (17.5%) |
| **Dementia** | 117 (0.8%) | 303 (1.2%) | 357 (1.5%) | 239 (1.5%) | 66 (1.5%) |

**S3.2: Patient characteristics for colorectal cancer stratified by calendar year of diagnosis**

| Calendar Year of Diagnosis | 2000 to 2004 | 2005 to 2009 | 2010 to 2014 | 2015 to 2019 | 2020 to 2021 |
|---|---|---|---|---|---|
| N | 8610 | 15705 | 16015 | 10264 | 3203 |
| Sex: Male (N[%]) | 4689 (54.5%) | 8655 (55.1%) | 8960 (55.9%) | 5708 (55.6%) | 1788 (55.8%) |
| Age (Median [IQR]) | 72 (63 to 79) | 72 (64 to 80) | 72 (64 to 80) | 72 (63 to 80) | 72 (63 to 80) |
| Age Groups N (%) | | | | | |
| 18-29 | 14 (0.2%) | 47 (0.3%) | 38 (0.2%) | 20 (0.2%) | <5 |
| 30-39 | 78 (0.9%) | 119 (0.8%) | 160 (1.0%) | 117 (1.1%) | 50 (1.6%) |
| 40-49 | 308 (3.6%) | 584 (3.7%) | 599 (3.7%) | 380 (3.7%) | 130 (4.1%) |
| 50-59 | 1111 (12.9%) | 1775 (11.3%) | 1807 (11.3%) | 1284 (12.5%) | 390 (12.2%) |
| 60-69 | 2044 (23.7%) | 3957 (25.2%) | 4190 (26.2%) | 2479 (24.2%) | 796 (24.9%) |
| 70-79 | 2943 (34.2%) | 5094 (32.4%) | 5046 (31.5%) | 3195 (31.1%) | 1008 (31.5%) |
| 80-89 | 1822 (21.2%) | 3643 (23.2%) | 3586 (22.4%) | 2357 (23.0%) | 710 (22.2%) |
| 90+ | 290 (3.4%) | 486 (3.1%) | 589 (3.7%) | 432 (4.2%) | 115 (3.6%) |
| Median days of prior history (IQR) | 1679 (918 to 3,338) | 2458 (1,575 to 3,783) | 3894 (2,848 to 5,102) | 5227 (3,992 to 6,183) | 6371 (4,506 to 7,186) |
| Atrial fibrillation | 393 (4.6%) | 964 (6.1%) | 1179 (7.4%) | 901 (8.8%) | 310 (9.7%) |
| Heart failure | 363 (4.2%) | 534 (3.4%) | 482 (3.0%) | 316 (3.1%) | 101 (3.2%) |
| Ischemic heart disease | 954 (11.1%) | 1724 (11.0%) | 1592 (9.9%) | 969 (9.4%) | 256 (8.0%) |
| Cerebrovascular disease | 474 (5.5%) | 908 (5.8%) | 1038 (6.5%) | 654 (6.4%) | 214 (6.7%) |
| Hyperlipidemia | 439 (5.1%) | 1386 (8.8%) | 1787 (11.2%) | 1074 (10.5%) | 323 (10.1%) |
| Hypertensive disorder | 1679 (19.5%) | 4247 (27.0%) | 4904 (30.6%) | 3124 (30.4%) | 978 (30.5%) |
| Pulmonary embolism | 65 (0.8%) | 170 (1.1%) | 243 (1.5%) | 173 (1.7%) | 73 (2.3%) |
| Venous Thrombosis | 412 (4.8%) | 776 (4.9%) | 888 (5.5%) | 585 (5.7%) | 187 (5.8%) |
| Type 2 diabetes | 456 (5.3%) | 1551 (9.9%) | 2007 (12.5%) | 1419 (13.8%) | 482 (15.0%) |
| Chronic liver disease | 13 (0.2%) | 41 (0.3%) | 67 (0.4%) | 55 (0.5%) | 25 (0.8%) |
| Renal impairment | 146 (1.7%) | 1813 (11.5%) | 2821 (17.6%) | 1652 (16.1%) | 459 (14.3%) |
| COPD | 413 (4.8%) | 905 (5.8%) | 1049 (6.6%) | 759 (7.4%) | 239 (7.5%) |
| Gastrointestinal hemorrhage | 2118 (24.6%) | 3054 (19.4%) | 2954 (18.4%) | 1678 (16.3%) | 441 (13.8%) |
| Osteoarthritis | 1385 (16.1%) | 2833 (18.0%) | 3487 (21.8%) | 2230 (21.7%) | 648 (20.2%) |
| Depressive disorder | 751 (8.7%) | 1338 (8.5%) | 1666 (10.4%) | 1213 (11.8%) | 384 (12.0%) |
| Dementia | 76 (0.9%) | 190 (1.2%) | 240 (1.5%) | 211 (2.1%) | 57 (1.8%) |

**S3.3: Patient characteristics for head and neck cancer stratified by calendar year of diagnosis**

| Calendar Year of Diagnosis | 2000 to 2004 | 2005 to 2009 | 2010 to 2014 | 2015 to 2019 | 2020 to 2021 |
|---|---|---|---|---|---|
| **N** | 1737 | 3422 | 3777 | 2749 | 770 |
| **Sex: Male (N[%])** | 1143 (65.8%) | 2361 (69.0%) | 2632 (69.7%) | 1952 (71.0%) | 526 (68.3%) |
| **Age (Median [IQR])** | 64 (55 to 74) | 64 (56 to 73) | 64 (56 to 73) | 65 (56 to 73) | 66 (57 to 74) |
| **Age Groups N (%)** | | | | | |
| **18-29** | 15 (0.9%) | 28 (0.8%) | 24 (0.6%) | 23 (0.8%) | 5 (0.6%) |
| **30-39** | 51 (2.9%) | 63 (1.8%) | 69 (1.8%) | 54 (2.0%) | 21 (2.7%) |
| **40-49** | 166 (9.6%) | 310 (9.1%) | 333 (8.8%) | 214 (7.8%) | 48 (6.2%) |
| **50-59** | 435 (25.0%) | 837 (24.5%) | 929 (24.6%) | 638 (23.2%) | 162 (21.0%) |
| **60-69** | 464 (26.7%) | 1008 (29.5%) | 1178 (31.2%) | 840 (30.6%) | 240 (31.2%) |
| **70-79** | 367 (21.1%) | 775 (22.6%) | 784 (20.8%) | 659 (24.0%) | 205 (26.6%) |
| **80-89** | 208 (12.0%) | 344 (10.1%) | 391 (10.4%) | 276 (10.0%) | 74 (9.6%) |
| **90+** | 31 (1.8%) | 57 (1.7%) | 69 (1.8%) | 45 (1.6%) | 15 (1.9%) |
| **Median days of prior history (IQR)** | 1629 (891 to 3,029) | 2326.5 (1,459 to 3,548) | 3695 (2,620 to 4,825) | 5118 (3,562 to 6,112) | 6144 (3,570 to 7,023) |
| **Atrial fibrillation** | 68 (3.9%) | 127 (3.7%) | 167 (4.4%) | 166 (6.0%) | 45 (5.8%) |
| **Heart failure** | 48 (2.8%) | 58 (1.7%) | 64 (1.7%) | 64 (2.3%) | 21 (2.7%) |
| **Ischemic heart disease** | 167 (9.6%) | 300 (8.8%) | 289 (7.7%) | 212 (7.7%) | 61 (7.9%) |
| **Cerebrovascular disease** | 105 (6.0%) | 167 (4.9%) | 227 (6.0%) | 175 (6.4%) | 53 (6.9%) |
| **Hyperlipidemia** | 84 (4.8%) | 253 (7.4%) | 337 (8.9%) | 266 (9.7%) | 56 (7.3%) |
| **Hypertensive disorder** | 317 (18.2%) | 812 (23.7%) | 951 (25.2%) | 701 (25.5%) | 185 (24.0%) |
| **Pulmonary embolism** | 5 (0.3%) | 24 (0.7%) | 25 (0.7%) | 27 (1.0%) | 5 (0.6%) |
| **Venous Thrombosis** | 41 (2.4%) | 99 (2.9%) | 164 (4.3%) | 118 (4.3%) | 29 (3.8%) |
| **Type 2 diabetes** | 72 (4.1%) | 230 (6.7%) | 325 (8.6%) | 288 (10.5%) | 78 (10.1%) |
| **Chronic liver disease** | 13 (0.7%) | 39 (1.1%) | 57 (1.5%) | 55 (2.0%) | 14 (1.8%) |
| **Renal impairment** | 28 (1.6%) | 242 (7.1%) | 425 (11.3%) | 294 (10.7%) | 78 (10.1%) |
| **COPD** | 137 (7.9%) | 290 (8.5%) | 380 (10.1%) | 316 (11.5%) | 94 (12.2%) |
| **Gastrointestinal hemorrhage** | 85 (4.9%) | 185 (5.4%) | 249 (6.6%) | 221 (8.0%) | 57 (7.4%) |
| **Osteoarthritis** | 240 (13.8%) | 478 (14.0%) | 623 (16.5%) | 462 (16.8%) | 135 (17.5%) |
| **Depressive disorder** | 194 (11.2%) | 413 (12.1%) | 516 (13.7%) | 424 (15.4%) | 108 (14.0%) |
| **Dementia** | 16 (0.9%) | 37 (1.1%) | 49 (1.3%) | 49 (1.8%) | 12 (1.6%) |

**S3.4: Patient characteristics for liver cancer stratified by calendar year of diagnosis**

| Calendar Year of Diagnosis | 2000 to 2004 | 2005 to 2009 | 2010 to 2014 | 2015 to 2019 | 2020 to 2021 |
|---|---|---|---|---|---|
| **N** | 334 | 954 | 1262 | 1093 | 356 |
| **Sex: Male (N[%])** | 226 (67.7%) | 640 (67.1%) | 927 (73.5%) | 791 (72.4%) | 264 (74.2%) |
| **Age (Median [IQR])** | 70 (61 to 79) | 70 (61 to 77) | 71 (63 to 79) | 71 (63 to 78) | 72 (65 to 78) |
| **Age Groups N (%)** | | | | | |
| **18-29** | <5 | <5 | <5 | <5 | <5 |
| **30-39** | 7 (2.1%) | 7 (0.7%) | 8 (0.6%) | <5 | <5 |
| **40-49** | 21 (6.3%) | 46 (4.8%) | 40 (3.2%) | 21 (1.9%) | 10 (2.8%) |
| **50-59** | 43 (12.9%) | 143 (15.0%) | 173 (13.7%) | 165 (15.1%) | 36 (10.1%) |
| **60-69** | 89 (26.6%) | 262 (27.5%) | 328 (26.0%) | 302 (27.6%) | 90 (25.3%) |
| **70-79** | 100 (29.9%) | 305 (32.0%) | 423 (33.5%) | 368 (33.7%) | 141 (39.6%) |
| **80-89** | 65 (19.5%) | 173 (18.1%) | 268 (21.2%) | 209 (19.1%) | 68 (19.1%) |
| **90+** | 7 (2.1%) | 14 (1.5%) | 19 (1.5%) | 21 (1.9%) | 8 (2.2%) |
| **Median days of prior history (IQR)** | 1710 (832 to 3,080) | 2359 (1,430 to 3,754) | 3912 (2,890 to 5,153) | 5242 (4,019 to 6,217) | 6239 (4,591 to 7,170) |
| **Atrial fibrillation** | 11 (3.3%) | 59 (6.2%) | 106 (8.4%) | 87 (8.0%) | 35 (9.8%) |
| **Heart failure** | 16 (4.8%) | 47 (4.9%) | 53 (4.2%) | 39 (3.6%) | 11 (3.1%) |
| **Ischemic heart disease** | 39 (11.7%) | 135 (14.2%) | 158 (12.5%) | 146 (13.4%) | 47 (13.2%) |
| **Cerebrovascular disease** | 12 (3.6%) | 55 (5.8%) | 91 (7.2%) | 80 (7.3%) | 27 (7.6%) |
| **Hyperlipidemia** | 14 (4.2%) | 66 (6.9%) | 106 (8.4%) | 94 (8.6%) | 28 (7.9%) |
| **Hypertensive disorder** | 66 (19.8%) | 268 (28.1%) | 398 (31.5%) | 346 (31.7%) | 121 (34.0%) |
| **Pulmonary embolism** | 5 (1.5%) | 7 (0.7%) | 18 (1.4%) | 19 (1.7%) | <5 |
| **Venous Thrombosis** | 10 (3.0%) | 35 (3.7%) | 73 (5.8%) | 76 (7.0%) | 21 (5.9%) |
| **Type 2 diabetes** | 29 (8.7%) | 201 (21.1%) | 347 (27.5%) | 360 (32.9%) | 129 (36.2%) |
| **Chronic liver disease** | 40 (12.0%) | 145 (15.2%) | 260 (20.6%) | 279 (25.5%) | 115 (32.3%) |
| **Renal impairment** | 5 (1.5%) | 112 (11.7%) | 265 (21.0%) | 193 (17.7%) | 67 (18.8%) |
| **COPD** | 15 (4.5%) | 79 (8.3%) | 126 (10.0%) | 96 (8.8%) | 45 (12.6%) |
| **Gastrointestinal hemorrhage** | 41 (12.3%) | 104 (10.9%) | 132 (10.5%) | 132 (12.1%) | 45 (12.6%) |
| **Osteoarthritis** | 49 (14.7%) | 175 (18.3%) | 310 (24.6%) | 274 (25.1%) | 93 (26.1%) |
| **Depressive disorder** | 28 (8.4%) | 113 (11.8%) | 163 (12.9%) | 163 (14.9%) | 60 (16.9%) |
| **Dementia** | <5 | 7 (0.7%) | 19 (1.5%) | 20 (1.8%) | 6 (1.7%) |

**S3.5: Patient characteristics for lung cancer stratified by calendar year of diagnosis**

| Calendar Year of Diagnosis | 2000 to 2004 | 2005 to 2009 | 2010 to 2014 | 2015 to 2019 | 2020 to 2021 |
|---|---|---|---|---|---|
| **N** | 6916 | 12655 | 13187 | 9747 | 3058 |
| **Sex: Male (N[%])** | 4118 (59.5%) | 7105 (56.1%) | 6951 (52.7%) | 4865 (49.9%) | 1530 (50.0%) |
| **Age (Median [IQR])** | 72 (64 to 78) | 72 (64 to 79) | 72 (65 to 79) | 73 (66 to 79) | 73 (66 to 79) |
| **Age Groups N (%)** | | | | | |
| **18-29** | <5 | 8 (0.1%) | 9 (0.1%) | <5 | <5 |
| **30-39** | 25 (0.4%) | 36 (0.3%) | 34 (0.3%) | 23 (0.2%) | 11 (0.4%) |
| **40-49** | 181 (2.6%) | 318 (2.5%) | 300 (2.3%) | 189 (1.9%) | 42 (1.4%) |
| **50-59** | 822 (11.9%) | 1317 (10.4%) | 1351 (10.2%) | 894 (9.2%) | 276 (9.0%) |
| **60-69** | 1770 (25.6%) | 3542 (28.0%) | 3649 (27.7%) | 2513 (25.8%) | 775 (25.3%) |
| **70-79** | 2690 (38.9%) | 4560 (36.0%) | 4580 (34.7%) | 3719 (38.2%) | 1196 (39.1%) |
| **80-89** | 1284 (18.6%) | 2638 (20.8%) | 2857 (21.7%) | 2096 (21.5%) | 671 (21.9%) |
| **90+** | 142 (2.1%) | 236 (1.9%) | 407 (3.1%) | 309 (3.2%) | 86 (2.8%) |
| **Median days of prior history (IQR)** | 1827.5 (973 to 3,636) | 2468 (1,579 to 3,820) | 3867 (2,881 to 5,068) | 5252 (4,161 to 6,238) | 6367 (4,796 to 7,105) |
| **Atrial fibrillation** | 380 (5.5%) | 750 (5.9%) | 910 (6.9%) | 858 (8.8%) | 309 (10.1%) |
| **Heart failure** | 468 (6.8%) | 489 (3.9%) | 481 (3.6%) | 409 (4.2%) | 159 (5.2%) |
| **Ischemic heart disease** | 1055 (15.3%) | 1704 (13.5%) | 1788 (13.6%) | 1292 (13.3%) | 398 (13.0%) |
| **Cerebrovascular disease** | 540 (7.8%) | 922 (7.3%) | 1121 (8.5%) | 927 (9.5%) | 330 (10.8%) |
| **Hyperlipidemia** | 389 (5.6%) | 1208 (9.5%) | 1527 (11.6%) | 1135 (11.6%) | 327 (10.7%) |
| **Hypertensive disorder** | 1270 (18.4%) | 3333 (26.3%) | 3905 (29.6%) | 2961 (30.4%) | 935 (30.6%) |
| **Pulmonary embolism** | 69 (1.0%) | 170 (1.3%) | 243 (1.8%) | 234 (2.4%) | 88 (2.9%) |
| **Venous Thrombosis** | 318 (4.6%) | 585 (4.6%) | 695 (5.3%) | 542 (5.6%) | 191 (6.2%) |
| **Type 2 diabetes** | 306 (4.4%) | 1142 (9.0%) | 1516 (11.5%) | 1353 (13.9%) | 502 (16.4%) |
| **Chronic liver disease** | 18 (0.3%) | 53 (0.4%) | 69 (0.5%) | 69 (0.7%) | 38 (1.2%) |
| **Renal impairment** | 80 (1.2%) | 1465 (11.6%) | 2374 (18.0%) | 1716 (17.6%) | 521 (17.0%) |
| **COPD** | 1277 (18.5%) | 2660 (21.0%) | 3379 (25.6%) | 2910 (29.9%) | 937 (30.6%) |
| **Gastrointestinal hemorrhage** | 366 (5.3%) | 785 (6.2%) | 945 (7.2%) | 780 (8.0%) | 237 (7.8%) |
| **Osteoarthritis** | 1226 (17.7%) | 2448 (19.3%) | 3031 (23.0%) | 2370 (24.3%) | 766 (25.0%) |
| **Depressive disorder** | 815 (11.8%) | 1427 (11.3%) | 1951 (14.8%) | 1669 (17.1%) | 535 (17.5%) |
| **Dementia** | 76 (1.1%) | 154 (1.2%) | 224 (1.7%) | 252 (2.6%) | 66 (2.2%) |

**S3.6: Patient characteristics for oesophageal cancer stratified by calendar year of diagnosis**

| Calendar Year of Diagnosis | 2000 to 2004 | 2005 to 2009 | 2010 to 2014 | 2015 to 2019 | 2020 to 2021 |
|---|---|---|---|---|---|
| N | 2500 | 4323 | 4371 | 3061 | 915 |
| Sex: Male (N[%]) | 1655 (66.2%) | 2942 (68.1%) | 2963 (67.8%) | 2158 (70.5%) | 630 (68.9%) |
| Age (Median [IQR]) | 72 (63 to 80) | 71 (62 to 79) | 72 (63 to 80) | 72 (64 to 79) | 72 (64 to 80) |
| Age Groups N (%) | | | | | |
| 18-29 | <5 | <5 | <5 | <5 | 0 |
| 30-39 | 9 (0.4%) | 22 (0.5%) | 13 (0.3%) | 11 (0.4%) | 7 (0.8%) |
| 40-49 | 91 (3.6%) | 145 (3.4%) | 143 (3.3%) | 72 (2.4%) | 20 (2.2%) |
| 50-59 | 343 (13.7%) | 620 (14.3%) | 519 (11.9%) | 373 (12.2%) | 106 (11.6%) |
| 60-69 | 600 (24.0%) | 1131 (26.2%) | 1226 (28.0%) | 835 (27.3%) | 227 (24.8%) |
| 70-79 | 798 (31.9%) | 1389 (32.1%) | 1318 (30.2%) | 1031 (33.7%) | 320 (35.0%) |
| 80-89 | 562 (22.5%) | 892 (20.6%) | 940 (21.5%) | 614 (20.1%) | 198 (21.6%) |
| 90+ | 95 (3.8%) | 121 (2.8%) | 208 (4.8%) | 122 (4.0%) | 37 (4.0%) |
| Median days of prior history (IQR) | 1751 (996 to 3,556) | 2462 (1,602 to 3,732) | 3895 (2,884 to 5,204) | 5250 (4,081 to 6,186) | 6364 (4,816 to 7,214) |
| Atrial fibrillation | 113 (4.5%) | 233 (5.4%) | 321 (7.3%) | 259 (8.5%) | 83 (9.1%) |
| Heart failure | 111 (4.4%) | 167 (3.9%) | 147 (3.4%) | 107 (3.5%) | 36 (3.9%) |
| Ischemic heart disease | 305 (12.2%) | 514 (11.9%) | 481 (11.0%) | 318 (10.4%) | 88 (9.6%) |
| Cerebrovascular disease | 145 (5.8%) | 271 (6.3%) | 290 (6.6%) | 210 (6.9%) | 52 (5.7%) |
| Hyperlipidemia | 117 (4.7%) | 377 (8.7%) | 467 (10.7%) | 325 (10.6%) | 89 (9.7%) |
| Hypertensive disorder | 497 (19.9%) | 1140 (26.4%) | 1361 (31.1%) | 936 (30.6%) | 267 (29.2%) |
| Pulmonary embolism | 16 (0.6%) | 49 (1.1%) | 75 (1.7%) | 64 (2.1%) | 27 (3.0%) |
| Venous Thrombosis | 107 (4.3%) | 200 (4.6%) | 231 (5.3%) | 196 (6.4%) | 55 (6.0%) |
| Type 2 diabetes | 125 (5.0%) | 398 (9.2%) | 509 (11.6%) | 460 (15.0%) | 140 (15.3%) |
| Chronic liver disease | 7 (0.3%) | 25 (0.6%) | 21 (0.5%) | 30 (1.0%) | 9 (1.0%) |
| Renal impairment | 27 (1.1%) | 467 (10.8%) | 790 (18.1%) | 526 (17.2%) | 122 (13.3%) |
| COPD | 156 (6.2%) | 336 (7.8%) | 391 (8.9%) | 350 (11.4%) | 101 (11.0%) |
| Gastrointestinal hemorrhage | 168 (6.7%) | 309 (7.1%) | 352 (8.1%) | 251 (8.2%) | 76 (8.3%) |
| Osteoarthritis | 401 (16.0%) | 780 (18.0%) | 932 (21.3%) | 714 (23.3%) | 192 (21.0%) |
| Depressive disorder | 227 (9.1%) | 383 (8.9%) | 492 (11.3%) | 355 (11.6%) | 110 (12.0%) |
| Dementia | 22 (0.9%) | 54 (1.2%) | 82 (1.9%) | 47 (1.5%) | 15 (1.6%) |

**S3.7: Patient characteristics for pancreatic cancer stratified by calendar year of diagnosis**

| Calendar Year of Diagnosis | 2000 to 2004 | 2005 to 2009 | 2010 to 2014 | 2015 to 2019 | 2020 to 2021 |
|---|---|---|---|---|---|
| N | 1330 | 2805 | 3047 | 2211 | 723 |
| Sex: Male (N[%]) | 678 (51.0%) | 1365 (48.7%) | 1517 (49.8%) | 1094 (49.5%) | 381 (52.7%) |
| Age (Median [IQR]) | 73 (63 to 80) | 72 (64 to 80) | 73 (65 to 80) | 73 (65 to 80) | 73 (65 to 80) |
| Age Groups N (%) | | | | | |
| 18-29 | 0 | <5 | <5 | <5 | 0 |
| 30-39 | 6 (0.5%) | 17 (0.6%) | 7 (0.2%) | <5 | <5 |
| 40-49 | 46 (3.5%) | 90 (3.2%) | 96 (3.2%) | 59 (2.7%) | 23 (3.2%) |
| 50-59 | 173 (13.0%) | 327 (11.7%) | 298 (9.8%) | 260 (11.8%) | 78 (10.8%) |
| 60-69 | 304 (22.9%) | 724 (25.8%) | 834 (27.4%) | 521 (23.6%) | 163 (22.5%) |
| 70-79 | 443 (33.3%) | 885 (31.6%) | 960 (31.5%) | 764 (34.6%) | 264 (36.5%) |
| 80-89 | 309 (23.2%) | 644 (23.0%) | 707 (23.2%) | 511 (23.1%) | 166 (23.0%) |
| 90+ | 49 (3.7%) | 117 (4.2%) | 142 (4.7%) | 91 (4.1%) | 27 (3.7%) |
| Median days of prior history (IQR) | 1903 (1,066 to 3,666) | 2584 (1,690 to 4,031) | 3978 (2,834 to 5,218) | 5225 (4,084 to 6,270) | 6315 (4,340 to 7,058) |
| Atrial fibrillation | 72 (5.4%) | 128 (4.6%) | 216 (7.1%) | 171 (7.7%) | 58 (8.0%) |
| Heart failure | 60 (4.5%) | 73 (2.6%) | 83 (2.7%) | 74 (3.3%) | 31 (4.3%) |
| Ischemic heart disease | 163 (12.3%) | 295 (10.5%) | 343 (11.3%) | 231 (10.4%) | 74 (10.2%) |
| Cerebrovascular disease | 91 (6.8%) | 158 (5.6%) | 200 (6.6%) | 154 (7.0%) | 46 (6.4%) |
| Hyperlipidemia | 69 (5.2%) | 253 (9.0%) | 354 (11.6%) | 258 (11.7%) | 71 (9.8%) |
| Hypertensive disorder | 282 (21.2%) | 758 (27.0%) | 917 (30.1%) | 699 (31.6%) | 211 (29.2%) |
| Pulmonary embolism | 6 (0.5%) | 33 (1.2%) | 51 (1.7%) | 60 (2.7%) | 24 (3.3%) |
| Venous Thrombosis | 73 (5.5%) | 169 (6.0%) | 225 (7.4%) | 163 (7.4%) | 40 (5.5%) |
| Type 2 diabetes | 147 (11.1%) | 510 (18.2%) | 670 (22.0%) | 547 (24.7%) | 191 (26.4%) |
| Chronic liver disease | <5 | 10 (0.4%) | 16 (0.5%) | 10 (0.5%) | <5 |
| Renal impairment | 14 (1.1%) | 347 (12.4%) | 550 (18.1%) | 392 (17.7%) | 96 (13.3%) |
| COPD | 54 (4.1%) | 176 (6.3%) | 252 (8.3%) | 202 (9.1%) | 59 (8.2%) |
| Gastrointestinal hemorrhage | 88 (6.6%) | 184 (6.6%) | 248 (8.1%) | 195 (8.8%) | 60 (8.3%) |
| Osteoarthritis | 249 (18.7%) | 613 (21.9%) | 772 (25.3%) | 529 (23.9%) | 183 (25.3%) |
| Depressive disorder | 148 (11.1%) | 294 (10.5%) | 406 (13.3%) | 298 (13.5%) | 83 (11.5%) |
| Dementia | 13 (1.0%) | 36 (1.3%) | 66 (2.2%) | 42 (1.9%) | 10 (1.4%) |

**S3.8: Patient characteristics for prostate cancer stratified by calendar year of diagnosis**

| Calendar Year of Diagnosis | 2000 to 2004 | 2005 to 2009 | 2010 to 2014 | 2015 to 2019 | 2020 to 2021 |
|---|---|---|---|---|---|
| N | 10364 | 18181 | 19077 | 13520 | 3783 |
| Sex: Male (N[%]) | 10364 (100.0%) | 18181 (100.0%) | 19077 (100.0%) | 13520 (100.0%) | 3783 (100.0%) |
| Age (Median [IQR]) | 73 (67 to 79) | 72 (65 to 79) | 71 (65 to 78) | 71 (65 to 77) | 72 (66 to 77) |
| Age Groups N (%) | | | | | |
| 18-29 | <5 | <5 | 0 | 0 | 0 |
| 30-39 | <5 | <5 | <5 | <5 | <5 |
| 40-49 | 48 (0.5%) | 102 (0.6%) | 176 (0.9%) | 99 (0.7%) | 23 (0.6%) |
| 50-59 | 796 (7.7%) | 1612 (8.9%) | 1739 (9.1%) | 1382 (10.2%) | 312 (8.2%) |
| 60-69 | 2776 (26.8%) | 5620 (30.9%) | 6322 (33.1%) | 4264 (31.5%) | 1186 (31.4%) |
| 70-79 | 4177 (40.3%) | 6863 (37.7%) | 7076 (37.1%) | 5308 (39.3%) | 1601 (42.3%) |
| 80-89 | 2286 (22.1%) | 3569 (19.6%) | 3328 (17.4%) | 2171 (16.1%) | 562 (14.9%) |
| 90+ | 277 (2.7%) | 409 (2.2%) | 434 (2.3%) | 293 (2.2%) | 97 (2.6%) |
| Median days of prior history (IQR) | 1714 (943 to 3,392) | 2534 (1,596 to 3,898) | 3988 (2,935 to 5,242) | 5279 (4,040 to 6,266) | 6316 (4,575 to 7,146) |
| Atrial fibrillation | 550 (5.3%) | 1128 (6.2%) | 1351 (7.1%) | 1084 (8.0%) | 335 (8.9%) |
| Heart failure | 459 (4.4%) | 560 (3.1%) | 455 (2.4%) | 391 (2.9%) | 99 (2.6%) |
| Ischemic heart disease | 1416 (13.7%) | 2191 (12.1%) | 2251 (11.8%) | 1469 (10.9%) | 383 (10.1%) |
| Cerebrovascular disease | 633 (6.1%) | 1014 (5.6%) | 1154 (6.0%) | 825 (6.1%) | 241 (6.4%) |
| Hyperlipidemia | 628 (6.1%) | 1710 (9.4%) | 2378 (12.5%) | 1605 (11.9%) | 399 (10.5%) |
| Hypertensive disorder | 2239 (21.6%) | 5128 (28.2%) | 6223 (32.6%) | 4347 (32.2%) | 1157 (30.6%) |
| Pulmonary embolism | 68 (0.7%) | 141 (0.8%) | 202 (1.1%) | 176 (1.3%) | 54 (1.4%) |
| Venous Thrombosis | 389 (3.8%) | 727 (4.0%) | 981 (5.1%) | 705 (5.2%) | 215 (5.7%) |
| Type 2 diabetes | 528 (5.1%) | 1436 (7.9%) | 2112 (11.1%) | 1617 (12.0%) | 461 (12.2%) |
| Chronic liver disease | 8 (0.1%) | 26 (0.1%) | 42 (0.2%) | 44 (0.3%) | 20 (0.5%) |
| Renal impairment | 268 (2.6%) | 2057 (11.3%) | 2992 (15.7%) | 1900 (14.1%) | 495 (13.1%) |
| COPD | 537 (5.2%) | 1098 (6.0%) | 1284 (6.7%) | 943 (7.0%) | 241 (6.4%) |
| Gastrointestinal hemorrhage | 655 (6.3%) | 1250 (6.9%) | 1590 (8.3%) | 1100 (8.1%) | 276 (7.3%) |
| Osteoarthritis | 1828 (17.6%) | 3488 (19.2%) | 4127 (21.6%) | 2873 (21.2%) | 800 (21.1%) |
| Depressive disorder | 591 (5.7%) | 1227 (6.7%) | 1542 (8.1%) | 1191 (8.8%) | 364 (9.6%) |
| Dementia | 67 (0.6%) | 138 (0.8%) | 236 (1.2%) | 163 (1.2%) | 43 (1.1%) |

**S3.9: Patient characteristics for stomach cancer stratified by calendar year of diagnosis**

| Calendar Year of Diagnosis | 2000 to 2004 | 2005 to 2009 | 2010 to 2014 | 2015 to 2019 | 2020 to 2021 |
|---|---|---|---|---|---|
| N | 1318 | 2467 | 2153 | 1090 | 305 |
| Sex: Male (N[%]) | 823 (62.4%) | 1538 (62.3%) | 1372 (63.7%) | 676 (62.0%) | 192 (63.0%) |
| Age (Median [IQR]) | 75 (66 to 81) | 75 (68 to 81) | 76 (67 to 82) | 75 (66 to 82) | 74 (65 to 82) |
| Age Groups N (%) | | | | | |
| 18-29 | <5 | 8 (0.3%) | 8 (0.4%) | <5 | 0 |
| 30-39 | 14 (1.1%) | 21 (0.9%) | 19 (0.9%) | 12 (1.1%) | <5 |
| 40-49 | 27 (2.0%) | 76 (3.1%) | 80 (3.7%) | 40 (3.7%) | 9 (3.0%) |
| 50-59 | 118 (9.0%) | 189 (7.7%) | 163 (7.6%) | 107 (9.8%) | 35 (11.5%) |
| 60-69 | 271 (20.6%) | 442 (17.9%) | 400 (18.6%) | 186 (17.1%) | 64 (21.0%) |
| 70-79 | 467 (35.4%) | 924 (37.5%) | 734 (34.1%) | 379 (34.8%) | 93 (30.5%) |
| 80-89 | 355 (26.9%) | 695 (28.2%) | 637 (29.6%) | 308 (28.3%) | 85 (27.9%) |
| 90+ | 63 (4.8%) | 112 (4.5%) | 112 (5.2%) | 57 (5.2%) | 16 (5.2%) |
| Median days of prior history (IQR) | 1812 (1,000 to 3,654) | 2488 (1,567 to 3,874) | 3873 (2,793 to 5,114) | 5232.5 (4,129 to 6,144) | 6288 (4,287 to 7,019) |
| Atrial fibrillation | 67 (5.1%) | 171 (6.9%) | 188 (8.7%) | 100 (9.2%) | 34 (11.1%) |
| Heart failure | 84 (6.4%) | 106 (4.3%) | 103 (4.8%) | 36 (3.3%) | 8 (2.6%) |
| Ischemic heart disease | 213 (16.2%) | 321 (13.0%) | 306 (14.2%) | 120 (11.0%) | 42 (13.8%) |
| Cerebrovascular disease | 105 (8.0%) | 160 (6.5%) | 183 (8.5%) | 84 (7.7%) | 30 (9.8%) |
| Hyperlipidemia | 67 (5.1%) | 198 (8.0%) | 223 (10.4%) | 124 (11.4%) | 23 (7.5%) |
| Hypertensive disorder | 253 (19.2%) | 648 (26.3%) | 703 (32.7%) | 332 (30.5%) | 80 (26.2%) |
| Pulmonary embolism | 16 (1.2%) | 26 (1.1%) | 45 (2.1%) | 19 (1.7%) | 6 (2.0%) |
| Venous thrombosis | 73 (5.5%) | 122 (4.9%) | 114 (5.3%) | 71 (6.5%) | 16 (5.2%) |
| Type 2 diabetes | 77 (5.8%) | 262 (10.6%) | 313 (14.5%) | 167 (15.3%) | 60 (19.7%) |
| Chronic liver disease | <5 | 5 (0.2%) | 7 (0.3%) | 7 (0.6%) | <5 |
| Renal impairment | 22 (1.7%) | 318 (12.9%) | 452 (21.0%) | 205 (18.8%) | 51 (16.7%) |
| COPD | 106 (8.0%) | 196 (7.9%) | 208 (9.7%) | 121 (11.1%) | 31 (10.2%) |
| Gastrointestinal hemorrhage | 126 (9.6%) | 256 (10.4%) | 209 (9.7%) | 111 (10.2%) | 30 (9.8%) |
| Osteoarthritis | 248 (18.8%) | 515 (20.9%) | 512 (23.8%) | 308 (28.3%) | 56 (18.4%) |
| Depressive disorder | 115 (8.7%) | 243 (9.9%) | 247 (11.5%) | 132 (12.1%) | 38 (12.5%) |
| Dementia | 15 (1.1%) | 41 (1.7%) | 34 (1.6%) | 39 (3.6%) | 8 (2.6%) |

**S4: Crude annual incidence rates (per 100000 person years) for nine cancers from 2000 to 2021 (red line indicating start of covid pandemic in 2020).**

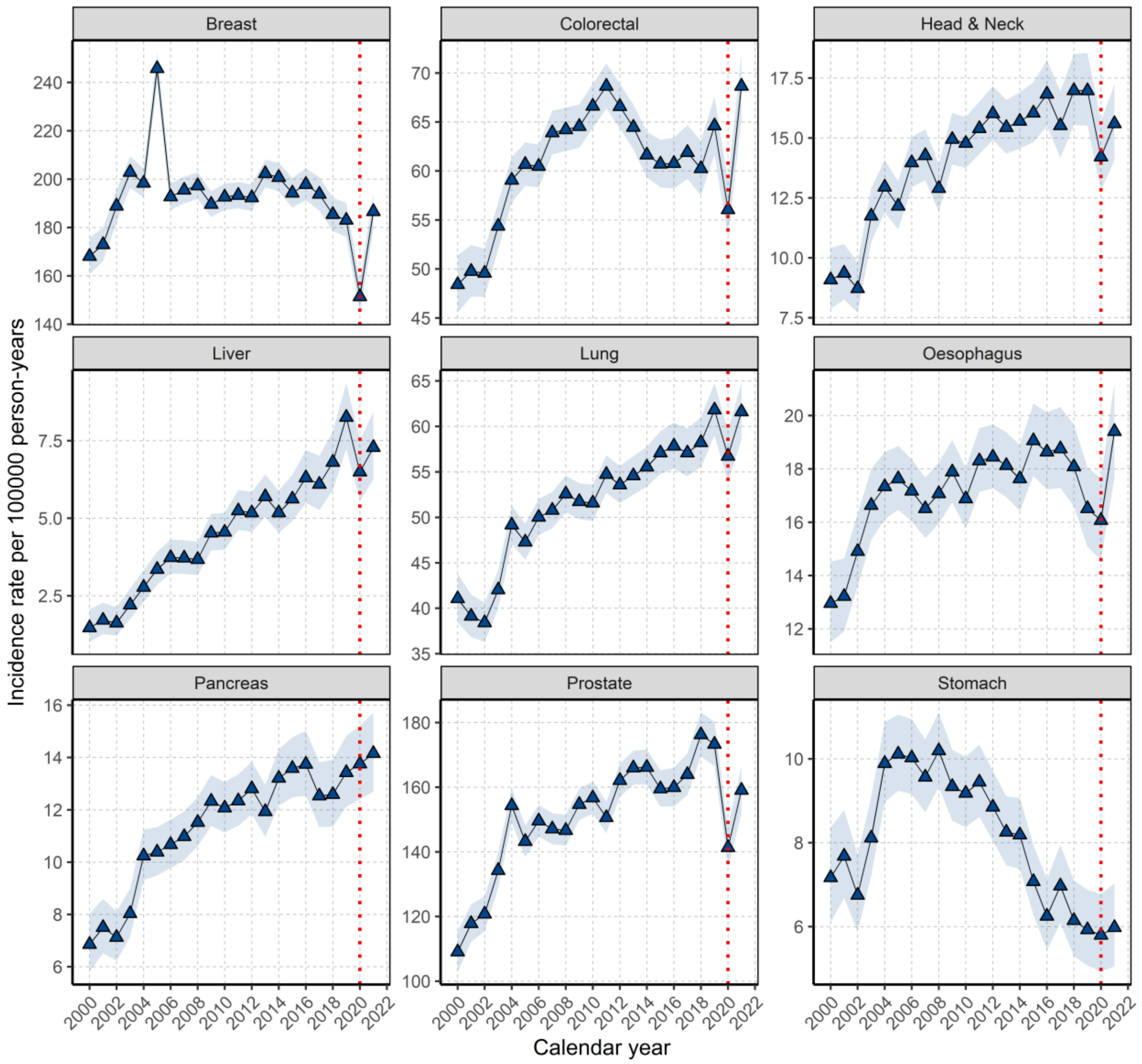

**S5: Individual KM survival curves for cancer outcomes stratified by calendar time of cancer diagnosis**

**S5.1: KM survival curve for breast cancer stratified by calendar time of cancer diagnosis**

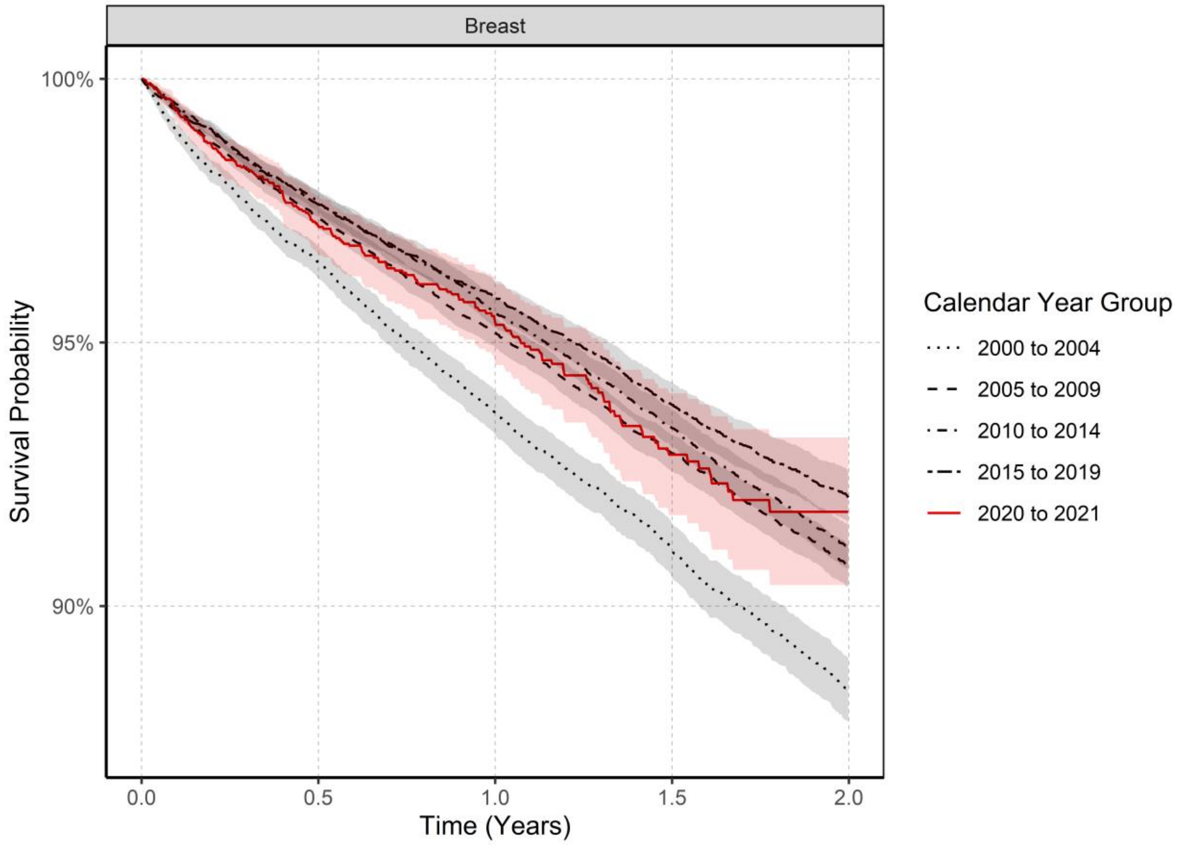

**S5.2: KM survival curve for colorectal cancer stratified by calendar time of cancer diagnosis**

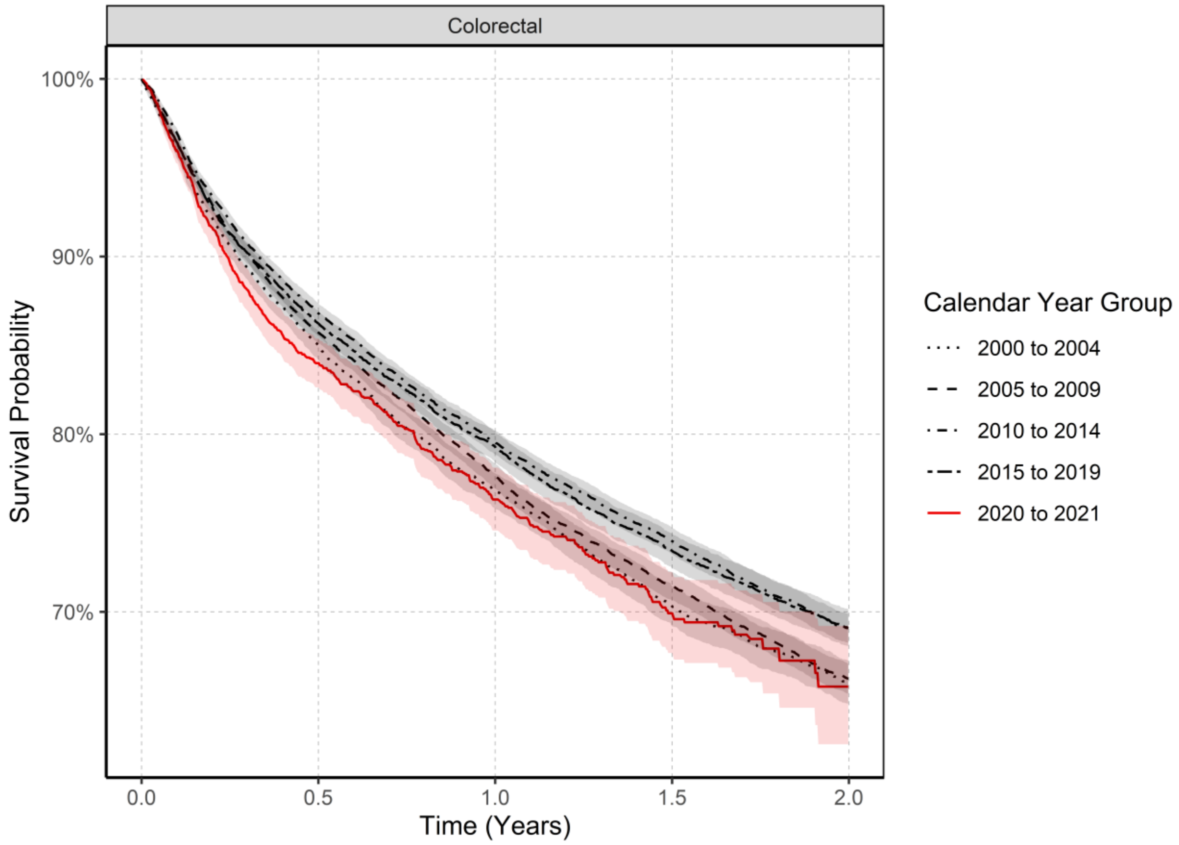

**S5.3: KM survival curve for head and neck cancers stratified by calendar time of cancer diagnosis**

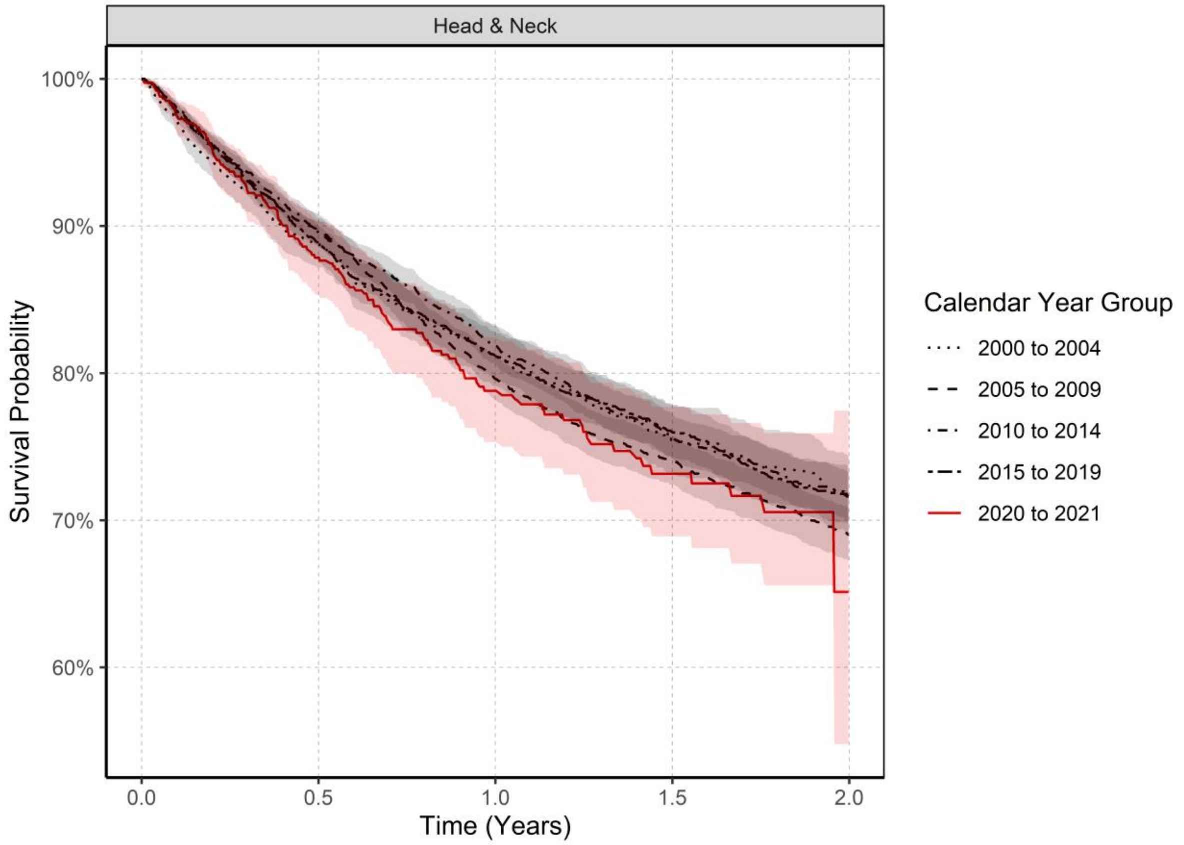

**S5.4: KM survival curve for liver cancer stratified by calendar time of cancer diagnosis**

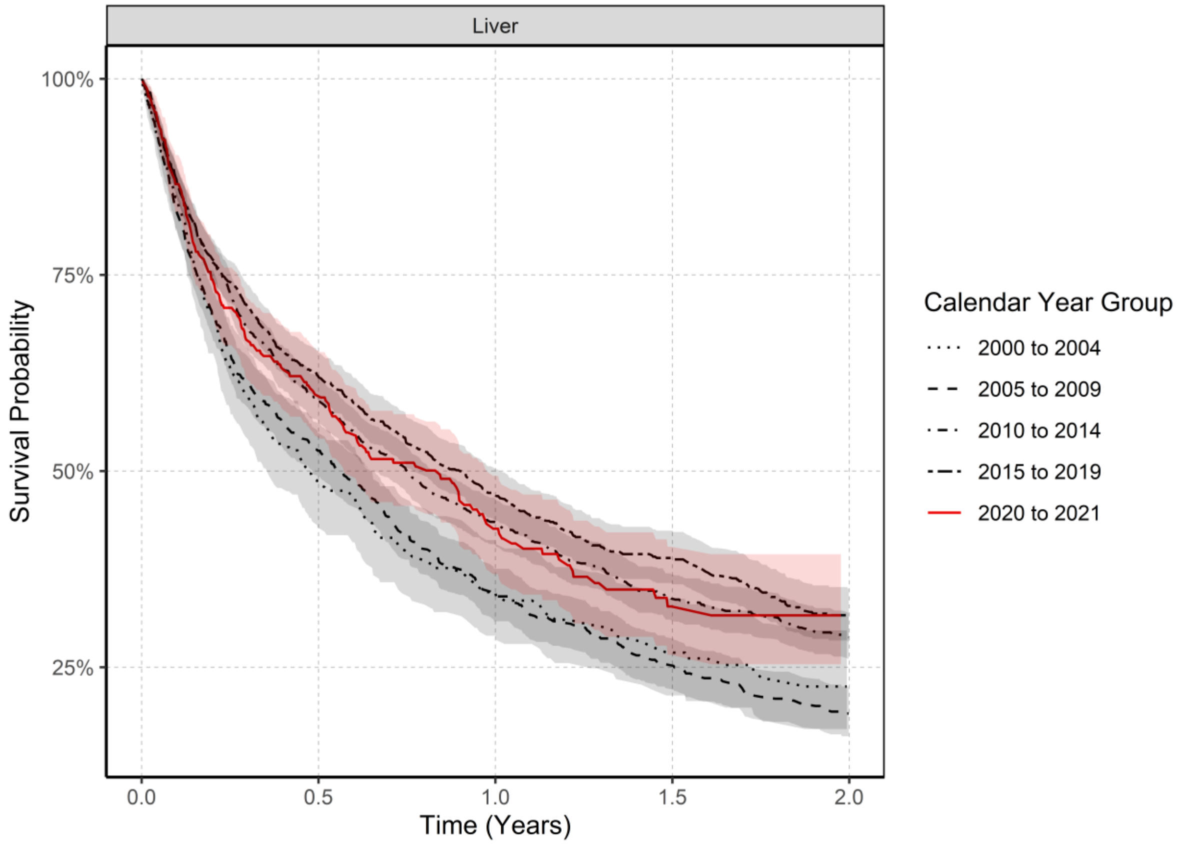

**S5.5: KM survival curve for lung cancer stratified by calendar time of cancer diagnosis**

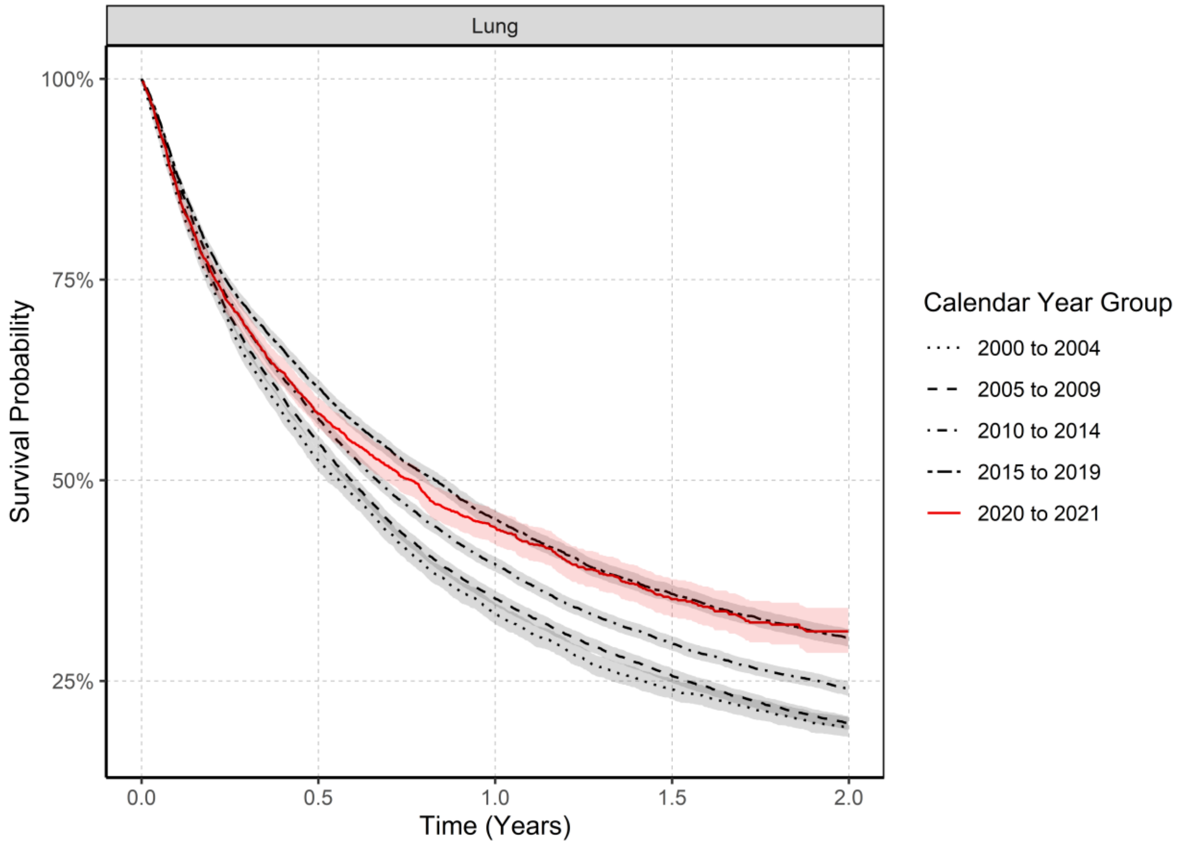

**S5.6: KM survival curve for oesophageal cancer stratified by calendar time of cancer diagnosis**

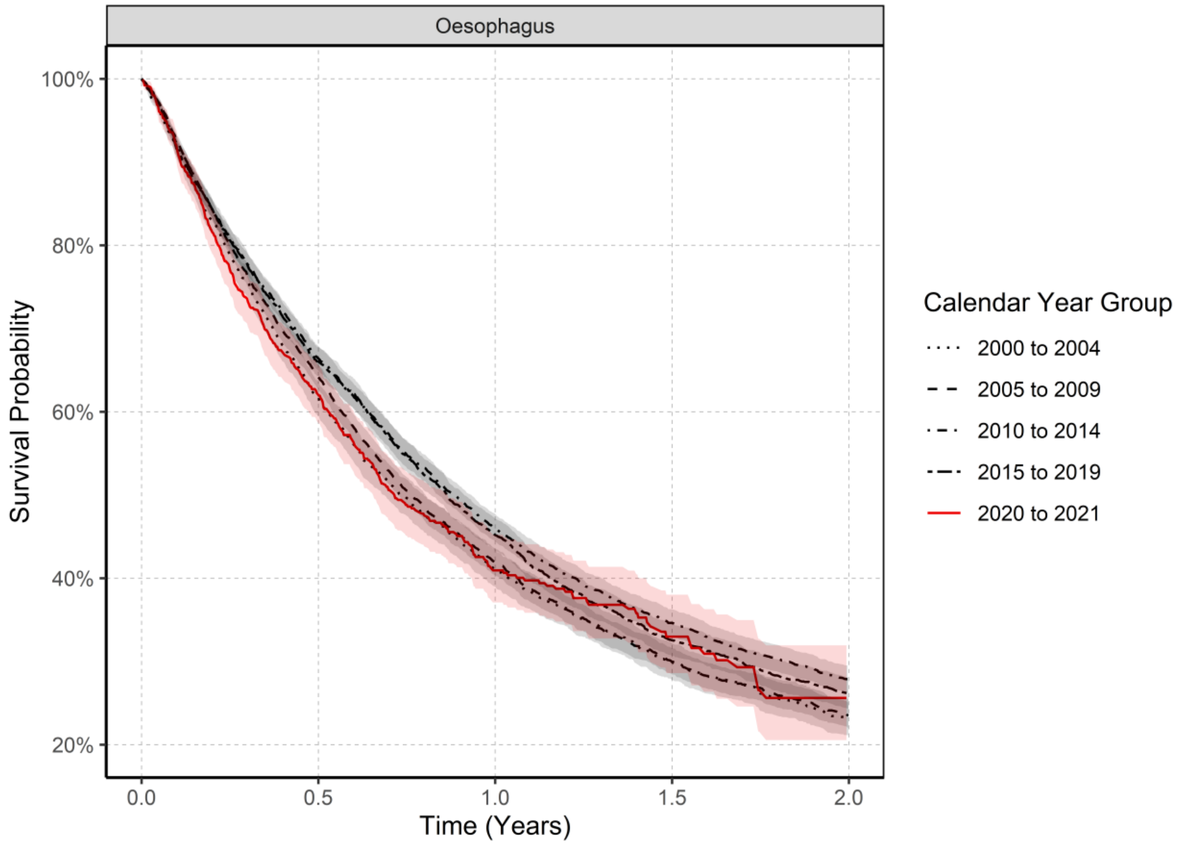

**S5.7: KM survival curve for pancreatic cancer stratified by calendar time of cancer diagnosis**

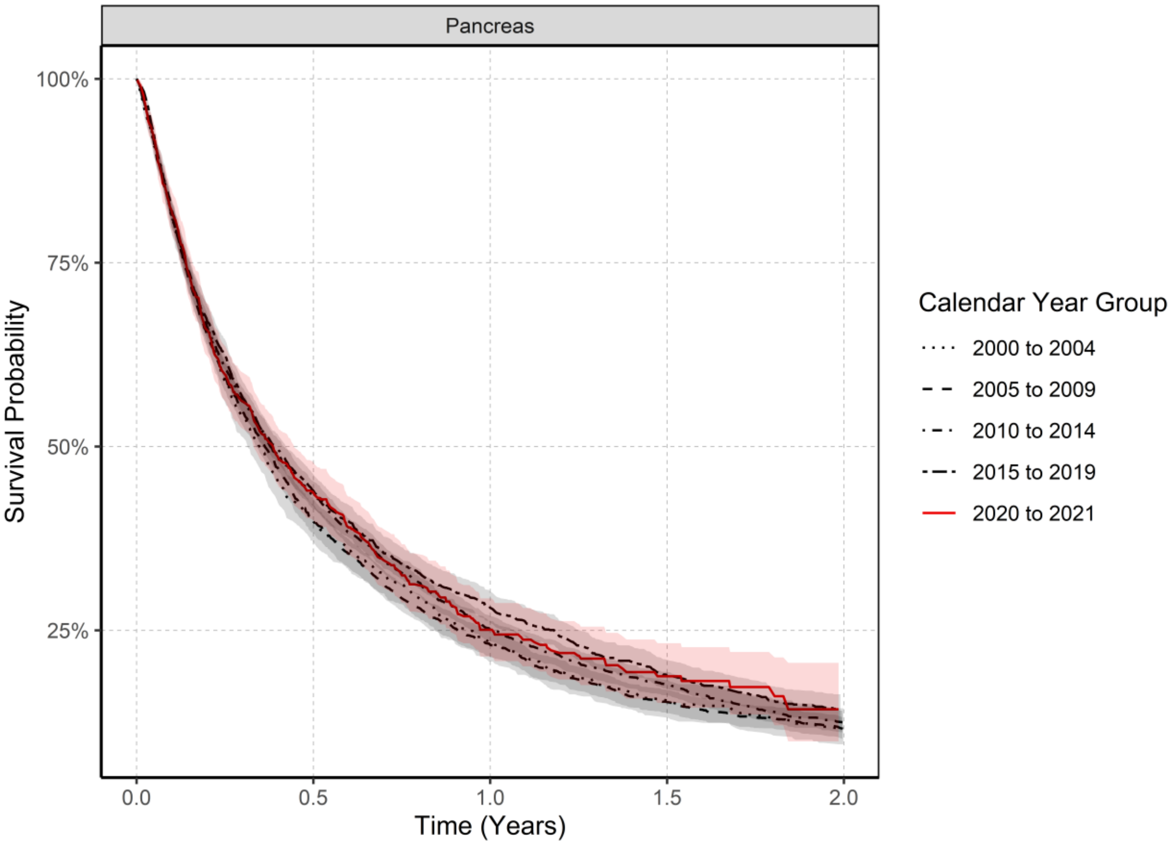

**S5.8: KM survival curve for prostate cancer stratified by calendar time of cancer diagnosis**

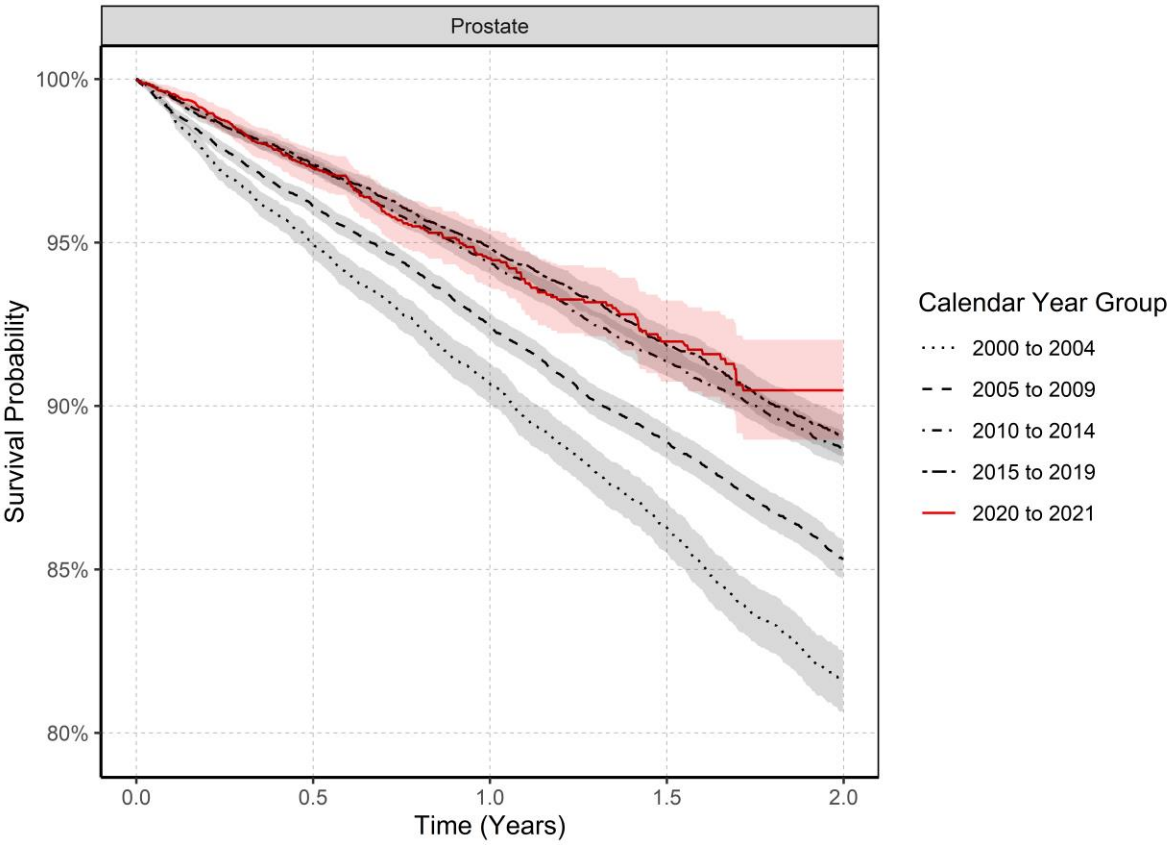

**S5.9: KM survival curve for stomach cancer stratified by calendar time of cancer diagnosis**

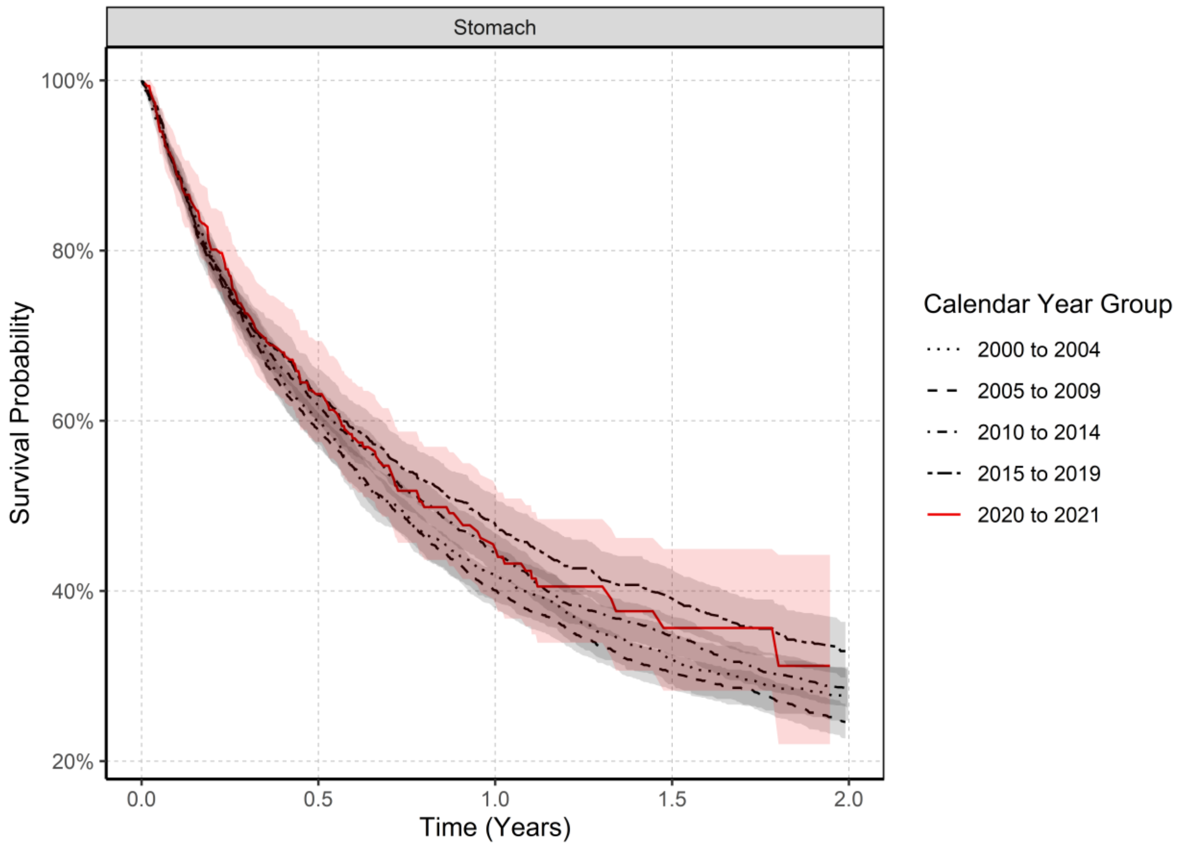

**S6: Numbers at risk, events, and censoring from KM survival data stratified by calendar time**

Below is a table showing the numbers of patients at risk, number of events and number censored and survival rates at each time point (0.5-2 years) from the KM survival curves. Missing values indicate results were censored due to values being <5.

| Cancer | Calendar Year | Time (Years) | Number at risk | Number of events | Number censored | Survival Rate % (95% CI) |
|---|---|---|---|---|---|---|
| Breast | 2000 to 2004 | 0.5 | 12684 | 500 | 2163 | 96.5 (96.2 to 96.8) |
| | | 1 | 10098 | 342 | 2244 | 93.7 (93.3 to 94.1) |
| | | 1.5 | 8130 | 253 | 1715 | 91.1 (90.6 to 91.6) |
| | | 2 | 6104 | 215 | 1811 | 88.4 (87.8 to 89.0) |
| | 2005 to 2009 | 0.5 | 21999 | 632 | 2871 | 97.4 (97.2 to 97.6) |
| | | 1 | 18953 | 469 | 2577 | 95.2 (94.9 to 95.5) |
| | | 1.5 | 15934 | 420 | 2599 | 92.9 (92.6 to 93.3) |
| | | 2 | 13173 | 338 | 2423 | 90.8 (90.4 to 91.2) |
| | 2010 to 2014 | 0.5 | 20463 | 523 | 2922 | 97.7 (97.5 to 97.9) |
| | | 1 | 17280 | 409 | 2774 | 95.6 (95.3 to 95.9) |
| | | 1.5 | 14374 | 365 | 2541 | 93.4 (93.0 to 93.7) |
| | | 2 | 11560 | 318 | 2496 | 91.1 (90.7 to 91.5) |
| | 2015 to 2019 | 0.5 | 13411 | 355 | 2152 | 97.6 (97.4 to 97.9) |
| | | 1 | 11252 | 227 | 1932 | 95.8 (95.5 to 96.2) |
| | | 1.5 | 9360 | 218 | 1674 | 93.8 (93.4 to 94.2) |
| | | 2 | 7586 | 161 | 1613 | 92.1 (91.6 to 92.6) |
| | 2020 to 2021 | 0.5 | 2865 | 101 | 1343 | 97.2 (96.7 to 97.8) |
| | | 1 | 1732 | 43 | 1090 | 95.4 (94.6 to 96.2) |
| | | 1.5 | 786 | 32 | 914 | 92.9 (91.7 to 94.0) |
| | | 2 | 0 | 7 | 779 | 91.8 (90.4 to 93.2) |
| Colorectal | 2000 to 2004 | 0.5 | 5900 | 1149 | 1242 | 85.0 (84.2 to 85.8) |
| | | 1 | 4349 | 523 | 1028 | 76.8 (75.8 to 77.8) |
| | | 1.5 | 3256 | 330 | 763 | 70.4 (69.3 to 71.5) |
| | | 2 | 2404 | 180 | 672 | 66.0 (64.8 to 67.3) |
| | 2005 to 2009 | 0.5 | 11639 | 2085 | 1759 | 85.7 (85.2 to 86.3) |
| | | 1 | 9128 | 1023 | 1488 | 77.7 (77.0 to 78.4) |
| | | 1.5 | 7194 | 680 | 1254 | 71.5 (70.7 to 72.3) |
| | | 2 | 5492 | 482 | 1220 | 66.2 (65.4 to 67.1) |
| | 2010 to 2014 | 0.5 | 12177 | 1983 | 1748 | 86.8 (86.3 to 87.4) |
| | | 1 | 9770 | 962 | 1445 | 79.5 (78.9 to 80.2) |
| | | 1.5 | 7781 | 631 | 1358 | 74.0 (73.3 to 74.8) |
| | | 2 | 6092 | 481 | 1208 | 69.1 (68.3 to 69.9) |
| | 2015 to 2019 | 0.5 | 7541 | 1323 | 1361 | 86.3 (85.6 to 86.9) |
| | | 1 | 5842 | 562 | 1137 | 79.3 (78.5 to 80.1) |
| | | 1.5 | 4559 | 398 | 885 | 73.5 (72.5 to 74.4) |
| | | 2 | 3531 | 248 | 780 | 69.1 (68.1 to 70.1) |
| | 2020 to 2021 | 0.5 | 1837 | 447 | 907 | 84.0 (82.6 to 85.4) |
| | | 1 | 1007 | 133 | 697 | 76.3 (74.6 to 78.1) |
| | | 1.5 | 425 | 60 | 522 | 69.9 (67.7 to 72.2) |
| | | 2 | 0 | 13 | 412 | 65.8 (62.5 to 69.2) |

| | | | | | | |
|---|---|---|---|---|---|---|
| Head & Neck | 2000 to 2004 | 0.5 | 1247 | 177 | 276 | 88.7 (87.2 to 90.3) |
| | | 1 | 911 | 97 | 239 | 81.1 (79.1 to 83.2) |
| | | 1.5 | 667 | 57 | 187 | 75.4 (73.1 to 77.9) |
| | | 2 | 494 | 29 | 144 | 71.7 (69.1 to 74.3) |
| | 2005 to 2009 | 0.5 | 2672 | 327 | 405 | 89.8 (88.8 to 90.9) |
| | | 1 | 2018 | 281 | 373 | 79.6 (78.1 to 81.1) |
| | | 1.5 | 1631 | 133 | 254 | 74.0 (72.4 to 75.7) |
| | | 2 | 1272 | 102 | 257 | 69.0 (67.3 to 70.8) |
| | 2010 to 2014 | 0.5 | 2953 | 368 | 448 | 89.6 (88.6 to 90.6) |
| | | 1 | 2327 | 235 | 391 | 81.9 (80.6 to 83.3) |
| | | 1.5 | 1842 | 164 | 321 | 75.7 (74.2 to 77.3) |
| | | 2 | 1467 | 93 | 282 | 71.6 (70.0 to 73.3) |
| | 2015 to 2019 | 0.5 | 2087 | 283 | 370 | 88.9 (87.7 to 90.1) |
| | | 1 | 1636 | 169 | 282 | 81.2 (79.6 to 82.8) |
| | | 1.5 | 1284 | 95 | 257 | 76.1 (74.3 to 77.9) |
| | | 2 | 1014 | 69 | 201 | 71.7 (69.7 to 73.6) |
| | 2020 to 2021 | 0.5 | 460 | 78 | 230 | 87.8 (85.3 to 90.4) |
| | | 1 | 269 | 39 | 152 | 78.8 (75.3 to 82.4) |
| | | 1.5 | 124 | 14 | 131 | 73.2 (68.9 to 77.7) |
| | | 2 | - | - | - | 65.1 (54.8 to 77.4) |
| Liver | 2000 to 2004 | 0.5 | 112 | 142 | 52 | 48.4 (42.7 to 54.8) |
| | | 1 | 64 | 29 | 19 | 34.6 (29.0 to 41.2) |
| | | 1.5 | 38 | 12 | 14 | 26.9 (21.4 to 33.7) |
| | | 2 | - | - | - | 22.5 (17.1 to 29.7) |
| | 2005 to 2009 | 0.5 | 406 | 409 | 99 | 52.6 (49.3 to 56.1) |
| | | 1 | 224 | 134 | 48 | 34.0 (30.8 to 37.5) |
| | | 1.5 | 131 | 51 | 42 | 25.4 (22.3 to 28.8) |
| | | 2 | 77 | 29 | 25 | 19.1 (16.2 to 22.5) |
| | 2010 to 2014 | 0.5 | 637 | 478 | 126 | 59.3 (56.5 to 62.2) |
| | | 1 | 406 | 161 | 70 | 43.4 (40.6 to 46.5) |
| | | 1.5 | 263 | 84 | 59 | 33.7 (30.9 to 36.8) |
| | | 2 | 185 | 35 | 43 | 28.8 (26.0 to 31.9) |
| | 2015 to 2019 | 0.5 | 550 | 381 | 148 | 62.1 (59.2 to 65.2) |
| | | 1 | 348 | 124 | 78 | 47.0 (43.8 to 50.3) |
| | | 1.5 | 230 | 55 | 63 | 38.9 (35.7 to 42.4) |
| | | 2 | 149 | 41 | 40 | 31.4 (28.2 to 35.0) |
| | 2020 to 2021 | 0.5 | 153 | 127 | 72 | 59.8 (54.5 to 65.5) |
| | | 1 | 69 | 36 | 48 | 42.7 (36.8 to 49.4) |
| | | 1.5 | 29 | 13 | 27 | 32.7 (26.6 to 40.3) |
| | | 2 | - | - | - | 31.6 (25.4 to 39.4) |
| Lung | 2000 to 2004 | 0.5 | 2660 | 2732 | 858 | 52.5 (51.3 to 53.9) |
| | | 1 | 1346 | 878 | 436 | 33.4 (32.1 to 34.8) |
| | | 1.5 | 786 | 351 | 209 | 24.0 (22.8 to 25.3) |
| | | 2 | 473 | 141 | 172 | 19.2 (18.0 to 20.5) |
| | 2005 to 2009 | 0.5 | 5718 | 5169 | 1224 | 54.7 (53.8 to 55.7) |
| | | 1 | 3112 | 1895 | 711 | 35.3 (34.4 to 36.2) |
| | | 1.5 | 1885 | 778 | 449 | 25.8 (24.9 to 26.7) |
| | | 2 | 1172 | 400 | 313 | 19.8 (18.9 to 20.6) |
| | 2010 to 2014 | 0.5 | 6355 | 5098 | 1394 | 57.8 (56.9 to 58.7) |

| | | | | | | |
|---|---|---|---|---|---|---|
| | | 1 | 3652 | 1861 | 842 | 39.6 (38.7 to 40.5) |
| | | 1.5 | 2289 | 842 | 521 | 29.7 (28.8 to 30.6) |
| | | 2 | 1505 | 399 | 385 | 24.0 (23.2 to 24.9) |
| | 2015 to 2019 | 0.5 | 4995 | 3467 | 1188 | 61.6 (60.6 to 62.6) |
| | | 1 | 3065 | 1235 | 695 | 45.2 (44.1 to 46.3) |
| | | 1.5 | 2028 | 589 | 448 | 35.8 (34.8 to 37.0) |
| | | 2 | 1403 | 284 | 341 | 30.4 (29.3 to 31.5) |
| | 2020 to 2021 | 0.5 | 1244 | 1124 | 677 | 58.3 (56.4 to 60.2) |
| | | 1 | 607 | 254 | 383 | 44.1 (42.1 to 46.3) |
| | | 1.5 | 245 | 93 | 269 | 35.2 (32.9 to 37.7) |
| | | 2 | 0 | 19 | 226 | 31.2 (28.5 to 34.1) |
| Oesophagus | 2000 to 2004 | 0.5 | 1219 | 841 | 339 | 61.5 (59.5 to 63.6) |
| | | 1 | 639 | 365 | 215 | 41.2 (39.0 to 43.5) |
| | | 1.5 | 364 | 158 | 117 | 30.0 (27.9 to 32.3) |
| | | 2 | 214 | 74 | 76 | 23.2 (21.2 to 25.5) |
| | 2005 to 2009 | 0.5 | 2379 | 1427 | 457 | 64.3 (62.8 to 65.8) |
| | | 1 | 1329 | 770 | 280 | 42.0 (40.4 to 43.6) |
| | | 1.5 | 807 | 354 | 168 | 30.0 (28.5 to 31.6) |
| | | 2 | 524 | 161 | 122 | 23.5 (22.0 to 25.1) |
| | 2010 to 2014 | 0.5 | 2475 | 1356 | 499 | 66.5 (65.0 to 68.0) |
| | | 1 | 1476 | 706 | 293 | 46.1 (44.5 to 47.7) |
| | | 1.5 | 933 | 341 | 202 | 34.6 (33.1 to 36.3) |
| | | 2 | 622 | 166 | 145 | 27.9 (26.3 to 29.5) |
| | 2015 to 2019 | 0.5 | 1734 | 968 | 342 | 66.1 (64.3 to 67.8) |
| | | 1 | 1007 | 508 | 219 | 45.2 (43.3 to 47.2) |
| | | 1.5 | 609 | 262 | 136 | 32.6 (30.7 to 34.5) |
| | | 2 | 414 | 110 | 85 | 26.2 (24.4 to 28.1) |
| | 2020 to 2021 | 0.5 | 394 | 296 | 218 | 62.1 (58.7 to 65.7) |
| | | 1 | 149 | 108 | 137 | 40.9 (37.1 to 45.2) |
| | | 1.5 | 55 | 19 | 75 | 33.0 (28.6 to 38.0) |
| | | 2 | 0 | 9 | 46 | 25.6 (20.6 to 31.9) |
| Pancreas | 2000 to 2004 | 0.5 | 378 | 662 | 164 | 40.0 (37.2 to 43.1) |
| | | 1 | 170 | 142 | 66 | 23.3 (20.7 to 26.2) |
| | | 1.5 | 88 | 50 | 32 | 15.6 (13.3 to 18.4) |
| | | 2 | 52 | 20 | 16 | 11.7 (9.5 to 14.4) |
| | 2005 to 2009 | 0.5 | 916 | 1497 | 273 | 40.2 (38.3 to 42.2) |
| | | 1 | 456 | 370 | 90 | 23.0 (21.4 to 24.9) |
| | | 1.5 | 249 | 142 | 65 | 15.3 (13.8 to 16.9) |
| | | 2 | 161 | 56 | 32 | 11.6 (10.2 to 13.2) |
| | 2010 to 2014 | 0.5 | 1086 | 1580 | 318 | 43.4 (41.6 to 45.3) |
| | | 1 | 526 | 424 | 136 | 25.2 (23.5 to 26.9) |
| | | 1.5 | 319 | 145 | 62 | 17.7 (16.2 to 19.4) |
| | | 2 | 180 | 87 | 52 | 12.5 (11.1 to 14.1) |
| | 2015 to 2019 | 0.5 | 767 | 1127 | 291 | 44.1 (41.9 to 46.3) |
| | | 1 | 407 | 258 | 102 | 28.1 (26.1 to 30.3) |
| | | 1.5 | 227 | 121 | 59 | 19.0 (17.1 to 21.1) |
| | | 2 | 142 | 52 | 33 | 14.3 (12.6 to 16.3) |
| | 2020 to 2021 | 0.5 | 218 | 360 | 133 | 43.8 (40.1 to 47.9) |
| | | 1 | 79 | 79 | 60 | 25.1 (21.4 to 29.3) |

| | | | | | | |
|---|---|---|---|---|---|---|
| | | 1.5 | 31 | 16 | 32 | 18.8 (15.1 to 23.2) |
| | | 2 | - | - | - | 14.3 (9.9 to 20.6) |
| Prostate | 2000 to 2004 | 0.5 | 8031 | 471 | 1734 | 95.0 (94.5 to 95.4) |
| | | 1 | 6186 | 328 | 1517 | 90.7 (90.1 to 91.3) |
| | | 1.5 | 4778 | 271 | 1137 | 86.3 (85.5 to 87.1) |
| | | 2 | 3531 | 237 | 1010 | 81.6 (80.6 to 82.5) |
| | 2005 to 2009 | 0.5 | 15248 | 661 | 2177 | 96.1 (95.8 to 96.4) |
| | | 1 | 12649 | 543 | 2056 | 92.4 (92.0 to 92.9) |
| | | 1.5 | 10405 | 450 | 1794 | 88.9 (88.4 to 89.4) |
| | | 2 | 8378 | 387 | 1640 | 85.3 (84.7 to 85.9) |
| | 2010 to 2014 | 0.5 | 16362 | 475 | 2178 | 97.4 (97.1 to 97.6) |
| | | 1 | 13569 | 460 | 2333 | 94.4 (94.1 to 94.8) |
| | | 1.5 | 11102 | 402 | 2065 | 91.4 (90.9 to 91.8) |
| | | 2 | 8942 | 303 | 1857 | 88.7 (88.1 to 89.2) |
| | 2015 to 2019 | 0.5 | 11354 | 327 | 1817 | 97.4 (97.1 to 97.7) |
| | | 1 | 9310 | 275 | 1769 | 94.9 (94.5 to 95.3) |
| | | 1.5 | 7441 | 267 | 1602 | 91.9 (91.3 to 92.4) |
| | | 2 | 5913 | 203 | 1325 | 89.1 (88.5 to 89.7) |
| | 2020 to 2021 | 0.5 | 2639 | 88 | 1052 | 97.3 (96.7 to 97.8) |
| | | 1 | 1588 | 60 | 991 | 94.5 (93.6 to 95.4) |
| | | 1.5 | 794 | 32 | 762 | 92.0 (90.7 to 93.2) |
| | | 2 | 0 | 10 | 784 | 90.5 (89.0 to 92.0) |
| Stomach | 2000 to 2004 | 0.5 | 591 | 448 | 181 | 60.1 (57.2 to 63.0) |
| | | 1 | 325 | 165 | 101 | 41.8 (38.8 to 45.0) |
| | | 1.5 | 208 | 70 | 47 | 32.0 (29.0 to 35.3) |
| | | 2 | 128 | 25 | 55 | 27.7 (24.8 to 31.0) |
| | 2005 to 2009 | 0.5 | 1232 | 932 | 255 | 59.0 (57.0 to 61.1) |
| | | 1 | 729 | 369 | 134 | 40.1 (38.0 to 42.2) |
| | | 1.5 | 459 | 162 | 108 | 30.4 (28.4 to 32.6) |
| | | 2 | 311 | 81 | 67 | 24.6 (22.7 to 26.7) |
| | 2010 to 2014 | 0.5 | 1134 | 767 | 231 | 61.8 (59.6 to 63.9) |
| | | 1 | 708 | 297 | 129 | 44.5 (42.2 to 46.8) |
| | | 1.5 | 480 | 150 | 78 | 34.6 (32.5 to 37.0) |
| | | 2 | 327 | 80 | 73 | 28.4 (26.3 to 30.7) |
| | 2015 to 2019 | 0.5 | 583 | 378 | 120 | 63.0 (60.1 to 66.1) |
| | | 1 | 379 | 131 | 73 | 47.9 (44.8 to 51.2) |
| | | 1.5 | 270 | 64 | 45 | 39.3 (36.1 to 42.7) |
| | | 2 | 190 | 41 | 39 | 32.9 (29.8 to 36.4) |
| | 2020 to 2021 | 0.5 | 137 | 97 | 70 | 63.1 (57.5 to 69.4) |
| | | 1 | 61 | 31 | 45 | 45.5 (39.1 to 52.9) |
| | | 1.5 | 17 | 9 | 35 | 35.6 (28.3 to 44.9) |
| | | 2 | - | - | - | 31.2 (22.0 to 44.2) |
